# Low-burden metrics for monitoring healthy diets across contexts: A multi-country validation analysis using quantitative 24-hour dietary intake data

**DOI:** 10.1101/2025.06.20.25329980

**Authors:** Giles T. Hanley-Cook, Edward A. Frongillo, Isabela Fleury Sattamini, Luc Ingenbleek, Jennifer C. Coates, Bridget A. Holmes, the Healthy Diets Monitoring Initiative (HDMI)

## Abstract

**Background:** Unhealthy diets are a major public health concern. However, a lack of available and up-to-date quantitative dietary intake data, accurate low-burden data collection methods, and valid and interpretable metrics across contexts has hindered frequent monitoring of diets globally.

**Methods:** In this multi-country validation study, we compared the relationships between low-burden metrics of a healthy diet, identified by the FAO, UNICEF, and WHO Healthy Diets Monitoring Initiative, and a suite of reference metrics of dietary intake. To this end, we used harmonized open access quantitative 24-hour recall and food record data collected from 77,118 adolescent and adult females across 27 countries.

**Findings:** On any given day, non-consumption of sweet foods or sweet beverages was associated with greater population-level adherence to <10% energy from free sugars in available upper-middle income countries [odds ratio (95% CI): 5.35 (5.05, 5.66)]. Food group diversity score (FGDS) was positively associated with mean adequacy ratio of micronutrients [*β* of 1-SD change (95% CI): ∼11 percentage points (9, 12)], while NCD-Protect score best predicted consumption of ≥400 g/day of fruits and vegetables [range odds ratio of 1-SD changes (95% CI): 2.56 to 3.01 (2.40 to 3.13) in lower-middle and high income countries, respectively]. FGDS and Global Diet Quality Score Positive were associated with achieving ≥25 g/day of fibre and ≥3,510 mg/day of potassium across contexts. No indicator accurately predicted adherence to WHO guidelines for sodium intake or energy intake ranges for carbohydrates, lipids, and proteins.

**Interpretation:** Accurate low-burden dietary assessment methods can provide valid metrics that enable timely monitoring of key characteristics of healthy diets globally. Specifically, avoiding sweet foods or sweet beverages serves as an indicator for population-level adherence to the WHO guideline for free sugar intake, while metrics reflecting nutritious food group diversity are strongly predictive of better micronutrient adequacy and adherence to WHO guidelines for fruits and vegetables, fibre, and potassium intakes across settings.

**Research in context:** *Evidence before this study:* Unhealthy diets are a common cause of all forms of malnutrition and are the major risk factor for non-communicable diseases. However, a lack of up-to-date nationally representative quantitative dietary intake data, and a lack of consensus on fit-for-purpose lower-burden data collection methods and metrics, based on comparative epidemiological evidence, has hindered monitoring of healthy diets globally.

*Added value of this study:* Using open access quantitative 24-hour recall and food record data collected in 27 countries, our validation analyses compared the relationships between measures of a healthy diet, deemed most promising for global surveillance by the FAO, UNICEF, and WHO Healthy Diets Monitoring Initiative, and a suite of reference metrics of dietary intake, both within and across countries.

*Implications of all the available evidence:* Nationally representative quantitative dietary intake assessments remain the reference for nutrition. Nevertheless, complementary low-burden methods can yield metrics that are suitable for high-frequency monitoring of healthy diets. Specifically, on any given day, avoiding sweet foods or sweet beverages is an accurate population-level indicator for achieving the WHO guideline of <10% kcal/day from free sugars, while metrics capturing nutritious food group diversity are associated with better micronutrient adequacy, as well as, higher probabilities of reaching WHO guidelines of ≥400 and ≥25 g/day for fruits and vegetables and fibre, respectively, and ≥3,510 mg/day of potassium across contexts.

## Introduction

In 2024, a joint statement by the Food and Agriculture Organization of the United Nations (FAO) and the World Health Organization (WHO) defined a healthy diet as having four characteristics or sub-constructs, namely, adequacy in essential nutrients without excess, balance in energy intake and macronutrients, moderation in foods, nutrients, or other compounds associated with detrimental health effects, and diversity in nutritious foods within and across food groups (1).

Unhealthy diets, characterized by insufficient consumption of whole grains, fruits and vegetables, and excessive intakes of sodium and free sugars, are a common cause of all forms of malnutrition (2) and are the major risk factor for non-communicable diseases (NCDs), such as cardiovascular diseases, cancers, and type 2 diabetes (3). Consequently, unhealthy diets give rise to an estimated 10 to 12 million deaths annually (4). Frequent population-level monitoring of dietary intake is therefore essential to inform evidence-based public health policies and nutrition-specific and -sensitive interventions aimed at improving diets (5). These actions are paramount to reverse the stagnating progress against undernutrition, and rising rates of micronutrient deficiencies, overweight and obesity, and diet-related NCDs worldwide (6).

While nationally representative quantitative individual-level dietary assessments, such as multiple day 24-hour recalls and food records, henceforth referred to as 24-HRs for readability, are indispensable for identifying usual nutrient gaps and evaluating the impact of interventions over time (7), the persistent lack of up-to-date dietary intake data poses a significant challenge to monitoring the healthiness of diets (8). Despite advances in quantitative data collection methods and technology, many countries, particularly low-and middle-income countries, still face limitations in collecting, analysing, and reporting quantitative dietary intake on a regular basis at the population-level (9).

Quantitative dietary intake data should be collected as frequently as possible at national level. Available methods, however, remain labour-intensive and time-consuming, and require significant technical expertise and financial resources, which makes them impractical for national and global surveillance, especially in contexts with rapidly changing diets (10). This underscores the need for the ongoing development of lower-burden methods, which may provide simplified yet informative food group consumption data for timely monitoring of healthy diets worldwide. An additional barrier to global monitoring, however, is the limited evidence for available metrics – for which data can be collected using low-burden methods – being valid markers for one or more of the four priority characteristics of a healthy diet across countries (11,12).

If healthy diets are to become central to post-Sustainable Development Goals (SDGs) and World Health Assembly commitment frameworks, and countries are to be supported in recurrently collecting nationally representative dietary intake data, the nutrition community must reach consensus on fit-for-purpose low-burden methods and metrics (13). The Healthy Diets Monitoring Initiative (HDMI) – a partnership among FAO, the United Nations Children’s Fund (UNICEF), and WHO – identified a set of measures for which routine data collection was deemed most promising globally, while acknowledging that comparative studies assessing their construct validity and cross-context equivalence are urgently needed (14).

The objective of our research was to establish the construct validity and cross-context equivalence of low-burden metrics. To this end, we used multi-country quantitative 24-HR data to compare the strength of associations between metrics of a healthy diet and a suite of reference metrics of dietary intake, such as the WHO guidelines of consuming <2,000 mg/day of sodium (15) and ≥400 g/day of fruits and vegetables (16), within and across countries.

## Methods

Our research is reported using the Strengthening the Reporting of Observational Studies in Epidemiology**–**Nutritional Epidemiology (STROBE-nut) checklist (17).

Previous studies have shown that composite metrics, weighting both nutritious and unhealthy food groups, have limited comparability and interpretability across contexts, because a wide range of contrasting diets can theoretically return analogous values (11,18). Hence, the subsequent analyses, which follow a research protocol and statistical analysis plan finalised in May 2024, focused on metrics reflecting only one of the four priority characteristics of a healthy diet, which include moderation or nutrient adequacy.

### Data sources

Individual-level data were analysed from 77,118 non-pregnant females aged 15-49 years from 44 quantitative 24-hour dietary intake surveys, which were available open access on FAO/WHO GIFT in May 2024 (19). This population group was purposively selected because, at the time of data analysis, this was the broadest population group for which all priority metrics of a healthy diet had been assessed for validity, although against distinct outcomes **–** not all of which were reference metrics of dietary intake **–** and across different countries (12).

Details on the 44 surveys, including their sampling framework, timing and modality of data collection, dietary assessment method, and any analyses conducted by data providers (e.g., exclusion criteria, recipe disaggregation, food composition tables and databases used), are available on the FAO/WHO GIFT website: https://www.fao.org/gift-individual-food-consumption/data/en. All demographic and dietary intake data were either self-reported by the female respondent (41 surveys) or recorded by a trained interviewer (3 surveys).

The current study included 2,408 females from five low income countries (LIC; six surveys), 32,766 from 11 lower-middle income countries (LMIC; 24 surveys), 38,847 from eight upper-middle income countries (UMIC; ten surveys), and 1,867 from three high income countries (HIC; four surveys) (**Figure S1; Table S1**). For 3,665 females, for whom data on physiological status were missing, we assumed non-pregnant non-lactating status. No other missing data were imputed.

The objective of this research was not to estimate usual food or nutrient intake distributions at the population level; hence, for the 32 surveys with repeated 24-HR data among a sub-sample of females **–** ten included <100 repeats or subsamples <10% **–** only data from the first day of dietary intake assessment were used to calculate the metrics of a healthy diet and quantitative reference metrics of dietary intake.

### Metrics of a healthy diet

#### FoodEx2 matching

Previously, food and drink items and (decomposed) mixed dishes reported in the available surveys on FAO/WHO GIFT were coded with 2,102 unique European Food Safety Authority (EFSA) FoodEx2 food classification and description system base term codes (20). For the current study, at least three FoodEx2 experts from FAO independently assigned each FoodEx2 term code to the distinct food (sub)groups conventionally enumerated and used to construct the food group diversity score (FGDS) (21), Global Diet Quality Score Positive (GDQS+) and Negative (GDQS–) (22), NCD-Protect and NCD-Risk scores (23), and Nova ultra-processed food (UPF) score (24,25). In addition, each FoodEx2 term code was also assigned to one of four groups of the Nova food classification system. Further methodological details are provided below.

##### Food group diversity score

For the FGDS, which aims to measure the characteristic dietary diversity or nutrient adequacy, 17 food subgroups were aggregated into the 10 predefined food groups (21): 1) starchy staple foods; 2) beans, peas and lentils; 3) nuts and seeds; 4) dairy products (milk, yogurt, and cheese); 5) flesh foods (meat, fish and seafood, poultry, and liver or organ meats); 6) eggs; 7) dark green leafy vegetables; 8) vitamin A-rich fruits and vegetables; 9) other vegetables; and 10) other fruits. The FGDS (0–10 points) was constructed by summing the number of food groups consumed in at least 15 g/day. Other food items, or in other words their respective FoodEx2 codes, were assigned to 13 additional food groups also enumerated through the conventional survey module, but not used in the construction of FGDS (21): i) packaged salty snacks; ii) deep fried snacks; iii) instant noodles; iv) fast food restaurant foods; v) sweet foods; vi) sugar-sweetened beverages; vii) sweetened infusions; viii) insects and small protein foods; ix) wild plants; x) red palm oil; xi) other oils and fats; xii) condiments and seasonings; xiii) other beverages and foods. As part of the FGDS module, collection of food groups i-vii is recommended, while viii-xiii are optional (21).

##### Global Diet Quality Score Positive and Negative

For the GDQS+ (0-32 points), which aims to measure the characteristic nutrient adequacy, the 16 food groups are (22): 1) citrus fruits; 2) deep-orange fruits, 3) other fruits; 4) dark-green leafy vegetables; 5) cruciferous vegetables; 6) deep-orange vegetables; 7) other vegetables; 8) legumes; 9) deep-orange tubers; 10) nuts and seeds; 11) whole grains; 12) liquid oils; 13) fish and shellfish; 14) poultry and game meat; 15) low-fat dairy; and 16) eggs. Furthermore, for the GDQS– (0-17 points), which aims to measure the characteristic moderation, the nine food groups are (22): i) high-fat dairy (in milk equivalents); ii) red meat; iii) processed meat; iv) refined grains and baked goods; v) sweets and ice cream; vi) sugar-sweetened beverages; vii) juice; viii) white roots and tubers; and ix) purchased deep fried foods (doubly classified, thus also including the food group of the primary ingredient). Following supplementary guidance from *Intake* (2021), amounts of hard cheeses consumed (g/day) were converted to milk equivalents using a conversion factor of 6.1 (26). Points were assigned based on three (e.g., 0, 1, 2) or four categories (e.g., 0, 1, 2, 0) of consumed quantities in g/day, specific to each food group. In brief, food groups 1-16 are defined as healthy (i.e., more points for higher g/day intake), food groups i and ii are classified as unhealthy when consumed in excessive amounts (i.e., increasing points are given until specific amounts have been consumed, after which no points are given), and food groups iii-ix are defined as unhealthy (i.e., more points for lower g/day intake) (22). GDQS+ and GDQS– were obtained by summing food groups 1-16 and i-ix (i.e., lower GDQS–when more and larger amounts of unhealthy food groups are consumed), respectively.

##### NCD-Protect and NCD-Risk scores

For the NCD-Protect score (0-9 points), which aims to measure the characteristic moderation or nutrient adequacy, the nine food groups are (23): 1) whole grains; 2) pulses; 3) nuts and seeds; 4) vitamin A-rich orange vegetables; 5) dark-green leafy vegetables; 6) other vegetables; 7) vitamin A-rich fruits; 8) citrus; and 9) other fruits. For the NCD-Risk score (0-9 points), which aims to measure moderation, 10 food subgroups were aggregated into the eight predefined food groups (23): i) sugar-sweetened beverages; ii) baked/grain-based sweets; iii) other sweets; iv) processed meat; v) unprocessed red meat; vi) deep fried foods; vii) fast foods and instant noodles; and viii) packaged ultra-processed salty snacks. The consumption of food groups 1-9 in at least 15 g/day quantities scores 1 point each towards NCD-Protect score, whereas for NCD-Risk score the consumption of food groups i-viii scores 1 point each – except food group iv, which scores 2 points (i.e., higher NCD-Risk score when a greater number of unhealthy food groups are consumed). Other food sub-groups, which are enumerated through the DQQ, are reflected in the FGDS or the optional and recommended food groups captured by the FGDS module (27).

##### Nova ultra-processed food score

For the Nova UPF score, which aims to measure the characteristic moderation, the 23 UPF subgroups used in the available Brazilian and Colombian screener are (24,25): 1) margarine; 2) loaf, hot dog, or hamburger bread; 3) regular or diet soda; 4) biscuits with or without filling; 5) packaged snacks, shoestring potatoes or crackers; 6) chocolate bar or bonbon; 7) ham, salami, or mortadella; 8) sausage; hamburger, or nuggets; 9) fruit-or chocolate-flavoured yogurt; 10) canned or bottled fruit juice; 11) powdered drink mix; 12) mayonnaise, ketchup, or mustard; 13) ice cream or popsicle; 14) chocolate drink; 15) French fries, either frozen or from fast food restaurant chains; 16) instant noodles and packaged soup; 17) tea-based beverage (ice tea-type); 18) pizza, either frozen or from fast food restaurant chains; 19) frozen lasagne or other frozen ready-made meals; 20) ready-made salad sauce; 21) packaged cake; 22) cereal bar; and 23) breakfast cereal. The consumption of food groups 1-23 in quantities >0 g/day scores 1 point each. The Nova UPF score (0–23 points) was constructed by summing the number of UPF subgroups consumed.

### Reference metrics of dietary intake

Details of the primary and secondary quantitative reference metrics of dietary intake for each characteristic of a healthy diet – which can be calculated using available data on FAO/WHO GIFT **–** are presented in **Table 1**. Reference metrics requiring additional FoodEx2 base term code matching or external data sources are described in further detail below.

**Table 1.**
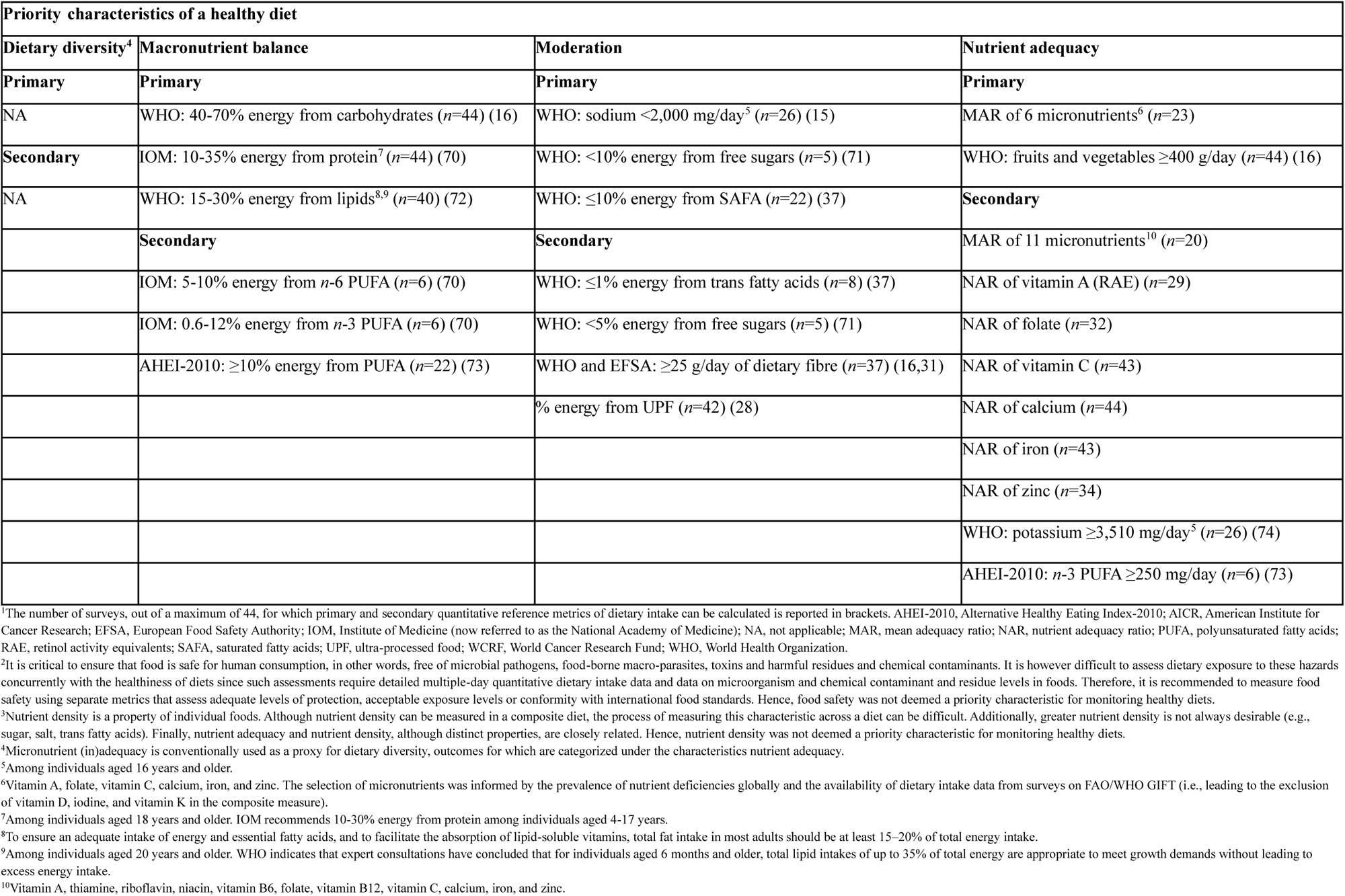
Primary and secondary quantitative reference metrics of dietary intake^1,2,3^.

For the characteristic moderation, food and drinks were defined as UPF when assigned to group four of the Nova food classification system. These are formulations of ingredients, mostly of exclusive industrial use, made by a series of industrial processes, many requiring sophisticated equipment and technology – hence ultra-processed. Examples include ready-to-consume products, such as carbonated soft drinks, sweet or savoury packaged snacks, chocolate, confectionery, ice-cream; mass-produced packaged breads and buns; margarines and other spreads; biscuits, pastries, cakes, and cake mixes, breakfast cereals, cereal and energy bars; energy drinks; milk drinks, fruit-flavoured yoghurts and drinks; cocoa drinks, instant sauces, any pre-prepared ready-to-heat products, including pies and pasta and pizza dishes, poultry and fish nuggets and sticks, sausages, burgers, hot dogs, and other reconstituted meat products, and powdered and packaged instant soups, noodles, and desserts (28).

For the characteristic nutrient adequacy, the nutrient adequacy ratio (NAR) value for a given nutrient is the ratio of an individual’s nutrient intake over the Estimated Average Requirement (EAR) for the corresponding sex (i.e., female), age (i.e., 15-17, 18-24, and 25-50 years), and physiological status category (i.e., non-pregnant non-lactating or non-pregnant lactating). We employed the harmonized EARs proposed by Allen and colleagues (2020) (29), which are a combination of values from the Institute of Medicine (30) and EFSA (31). The bioavailability of iron (i.e., low, medium, and high) and zinc (i.e., unrefined, semi-unrefined, semi-refined, and refined) were determined by a country’s Human Development Index in 2022 (i.e., low, medium, high, and very high) (32,33). NARs were truncated at one, indicating intake was equal to or above the EAR, so that high consumption of one micronutrient did not compensate for low consumption of another. For each non-pregnant female, the mean adequacy ratio (MAR) was calculated as the sum of NARs, divided by the number of micronutrients.

### Statistical analysis

Data management and statistical analysis were conducted in Stata (16.1, StataCorp LLC). All analyses were first conducted by World Bank country income classification (1 July 2023–30 June 2024) for didactic purposes (i.e., LIC, LMIC, UMIC, and HIC). Descriptive data are presented as means ± SD and median (*P*^25^ and *P*^75^) or percentages [95% confidence interval (CI)].

To assess the direction and magnitude of associations between mean-standardized measures of a healthy diet (e.g., FGDS and GDQS+ in *z*-scores) and reference measures (e.g., MAR of six micronutrients) or reference indicators of dietary intake (e.g., ≥400 g/day of fruits and vegetables), multilevel linear and logistic regression models were fitted, respectively, with a random intercept for survey, random slope for healthy diet measure, and an unstructured covariance matrix. For linear regression models, the normality of residuals was assessed by visual inspection of normal probability and quantile-normal plots. In cases where the assumption of normally distributed residuals was not met, log, cubic, square, inverse, inverse square, and inverse cubic transformations of the dependent variable were calculated to determine if they improved the model fit. If transformations did not improve the model fit, quantile regression models were used to compare the median, rather than the mean, of reference measures of dietary intake across values of measures of a healthy diet. For logistic regression models, the assumption of linearity in the logit (i.e., log odds) for measures of a healthy diet was assessed by also including a quadratic term in the model and examining the likelihood ratio test to determine the best model fit. Data visualisations were selected based on the strongest mean-standardized (i.e., comparative) associations, but crude values (i.e., original scale) rather than *z*-scores of the healthy diet measures were subsequently modelled for ease of interpretation.

If strong associations were observed **–** defined as a consistent odds ratio ≥1.5 for reference indicators of dietary intake, for each SD-increment in a healthy diet measure across World Bank income classifications **–** the analyses were repeated for individual surveys. Furthermore, the performance of various values of healthy diet measures, in other words indicator thresholds, for predicting a reference indicator of dietary intake, on a given day, were assessed using the area under the curve (AUC) from receiver operating characteristics (ROC) analyses and an informed assessment of the sensitivity, specificity, percentage correctly classified, and positive likelihood ratio test. Moreover, to further assess cross-context equivalence, the extent to which the mean (95% CI) of healthy diet measures differentiated the mean (95% CI) of reference measures of dietary intake across surveys, on a given day, was examined using range plots and Spearman’s rank correlations coefficients (*ρ*).

As sensitivity analyses, we repeated analyses of primary reference indicators of dietary intake using continuous (e.g., mg/day of sodium), rather than dichotomous dependent variables (e.g., < or ≥ 2,000 mg/day), to assess the direction of their relationships with healthy diet measures. Furthermore, as macronutrient balance also refers to energy intake without excess, we re-analysed relationships with % energy recommendations for carbohydrates, protein, and lipids among the sub-sample of females with dietary energy intakes between 500 and 3,500 kcal/day (34). Following a HDMI technical expert consultation held in February 2025, additional analyses were conducted to examine the associations between the consumption of three unhealthy foods groups, namely 1) sweet foods; 2) sweet beverages; and 3) fried and salty foods, and primary reference metrics for moderation, as well as the relationship between a 6-point Fruits and Vegetables-Global Dietary Recommendation (FV-GDR) score [i.e., food groups 4-9 of the NCD-Protect score (35)] and primary reference metrics for nutrient adequacy – noting that these metrics can be readily constructed from both the parsimonious FGDS module (21) and expanded DQQ (27).

To avoid misuse and misinterpretation of *P*-values, point estimates and 95% CIs are presented whenever possible (36).

## Results

### Sample characteristics

The median age of non-pregnant females was 24 years (16, 36) and 10.3% were lactating. FGDS was lowest in LIC and LMIC due to low consumption of animal-source food groups. GDQS+ and NCD-Protect score were highest in LIC and HIC, however, because of relatively high and diverse consumption of plant-based food groups, which was also reflected by the highest prevalence of females consuming ≥400 g/day of fruits and vegetables in these settings (**Tables S2-3**). Consumption of food groups, such as unprocessed red meat and white roots and tubers, are scored either positively (e.g., FGDS) or negatively (e.g., GDQS– and NCD-Risk score) depending on the measure used. Furthermore, consumption of food groups classified as unhealthy was greatest in HIC and UMIC, as reflected by higher NCD-Risk and Nova UPF scores and lower GDQS– (**Table S2**). Lastly, MAR of six or 11 micronutrients, % energy from UPF, and unprocessed red meat intakes were greatest among females in HIC, while the % achieving guidelines for sodium, saturated fatty acids, and fibre intakes were lowest in these settings (**Tables S3-4**).

### Associations between healthy diet measures and metrics of dietary intake

The assumptions for linear and logistic regression were met; therefore, untransformed (mean-standardized) dietary intake data were modelled.

#### Macronutrient balance

No healthy diet metric adequately predicted achieving % energy guidelines for carbohydrates or lipids, including for poly-unsaturated fatty acids, across the four World Bank income classifications (**Tables S5-6**).

While higher FGDS was moderately associated with a higher probability of consuming 10-35% of energy from protein across contexts [odds (OR) of 1-SD increment (95% CI): 1.62 (1.39, 1.89) in LIC to 2.40 (1.94, 2.97) in HIC] (**Figure 1**; **Table S5**), test characteristics for indicator thresholds were poor (**Figure S2**; **Table S7**), and the rank correlation between the 43 survey means of FGDS and the % achieving the energy guideline for protein was moderate (*ρ*=0.43) (**Figure S3**; **Table S8**).

Sensitivity analyses confirmed our main findings (data not shown).

**Figure 1.**
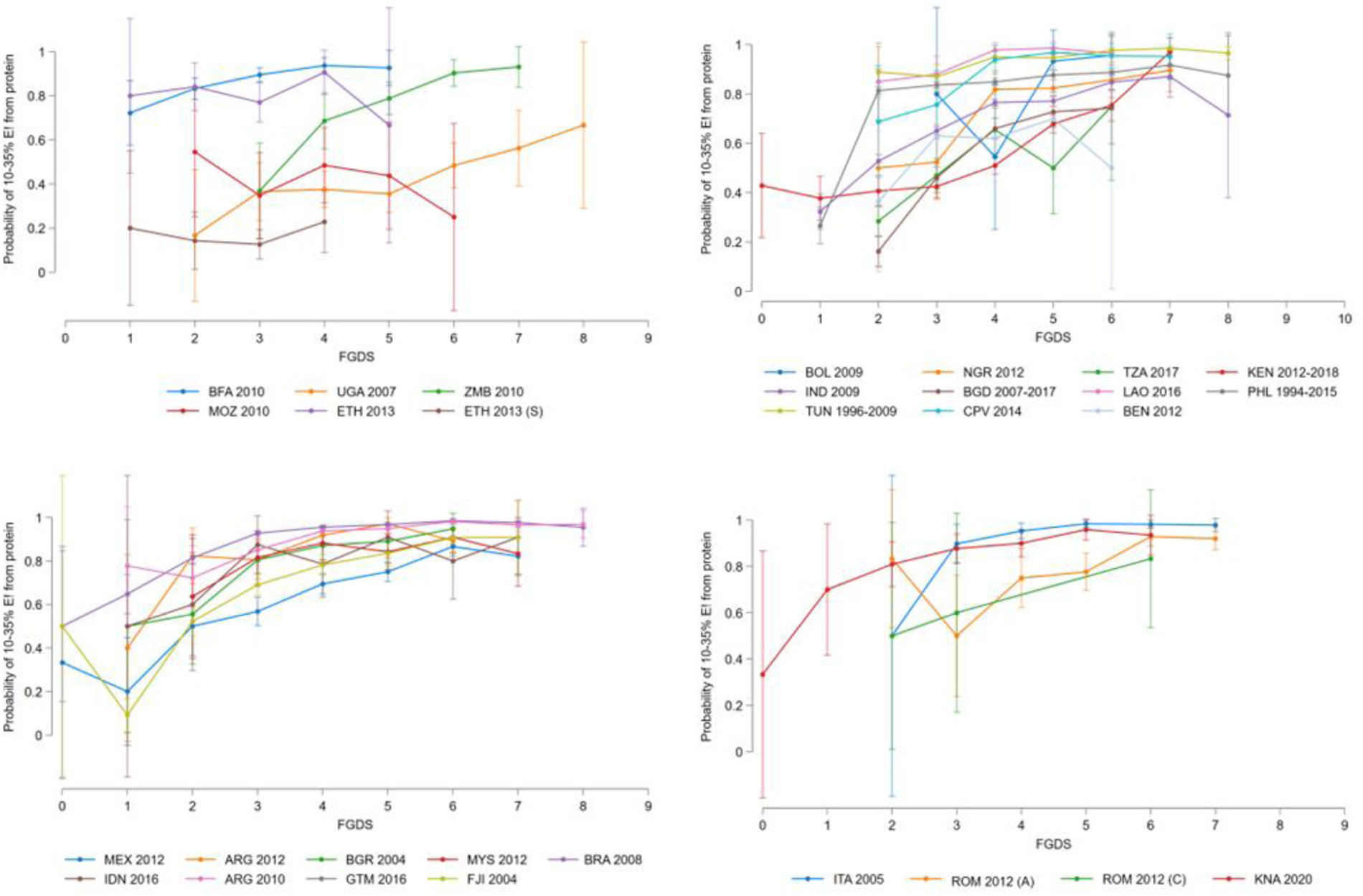
Relationship between the probability of 10-35% of dietary energy intake (E!) from protein and food group diversity score (FGDS) among non-pregnant females (18–49 years) in low (top left), lower-middle (top right), upper-middle (bottom left), and high income countries (bottom right). Error bars represent the 95% confidence intervals around the predicted mean probabilities from logistic regression models. ARG, Republic of Argentina; BEN, Republic of Benin; BFA, Burkina Faso; BGD, People’s Republic of Bangladesh; BGR, Republic of Bulgaria; BOL, Plurinational State of Bolivia; BRA, Federative Republic of Brazil; CPV, Republic of Cabo Verde; ETH, Federal Democratic Republic of Ethiopia; FJI, Republic of Fiji; GTM, Republic of Guatemala; HIC, IDN, Republic of Indonesia; IND, Republic of India; ITA, Republic of Italy; KEN, Republic of Kenya; KNA, Saint Kitts and Nevis; LAO, Lao People’s Democratic Republic; MEX, United Mexican States; MOZ, Republic of Mozambique; MYS, Malaysia; NGA, Federal Republic of Nigeria; PHL, Republic of the Philippines; ROM, Romania; TUN, Tunisian Republic; TZA, United Republic of Tanzania; UGA, Republic of Uganda; ZMB, Republic of Zambia.

#### Moderation

No healthy diet metric accurately predicted adherence to <2,000 mg/day of sodium or % energy guidelines for saturated or trans-fatty acids (**Tables S5-6**). Nevertheless, sensitivity analyses indicated that higher GDQS– was negatively associated with absolute sodium intakes across World Bank country income classifications, while similar findings were observed for lower NCD-Risk and Nova UPF scores in LIC, UMIC, and HIC (data not shown).

Furthermore, lower Nova UPF score was strongly associated with a higher likelihood of consuming <10% of energy from free sugars in UMIC [OR of 1-SD increment (95% CI): 3.52 (3.40, 3.64)], while no data were available for other World Bank country income classifications (**Figure 2**; **Table S5**). Moreover, test characteristics for a Nova UPF score ≤3 were borderline acceptable across the five surveys (**Figure S4**; **Table S9**), and the rank correlations between the survey means of Nova UPF score and % achieving the energy guidelines for free sugars were monotonic (*ρ*=1.00) (**Figure S5; Tables S8 and S10**). GDQS– and NCD-Risk score performed statistically equivalently to one another (**Table S5)**, while test characteristics were most acceptable for NCD-Risk scores ≤2 (**Figure S6**; **Table S9**), and NCD-Risk score also showed strong rank correlations with % energy guidelines for free sugars across the available surveys (**Figure S7**; **Tables S8 and S10**).

**Figure 2.**
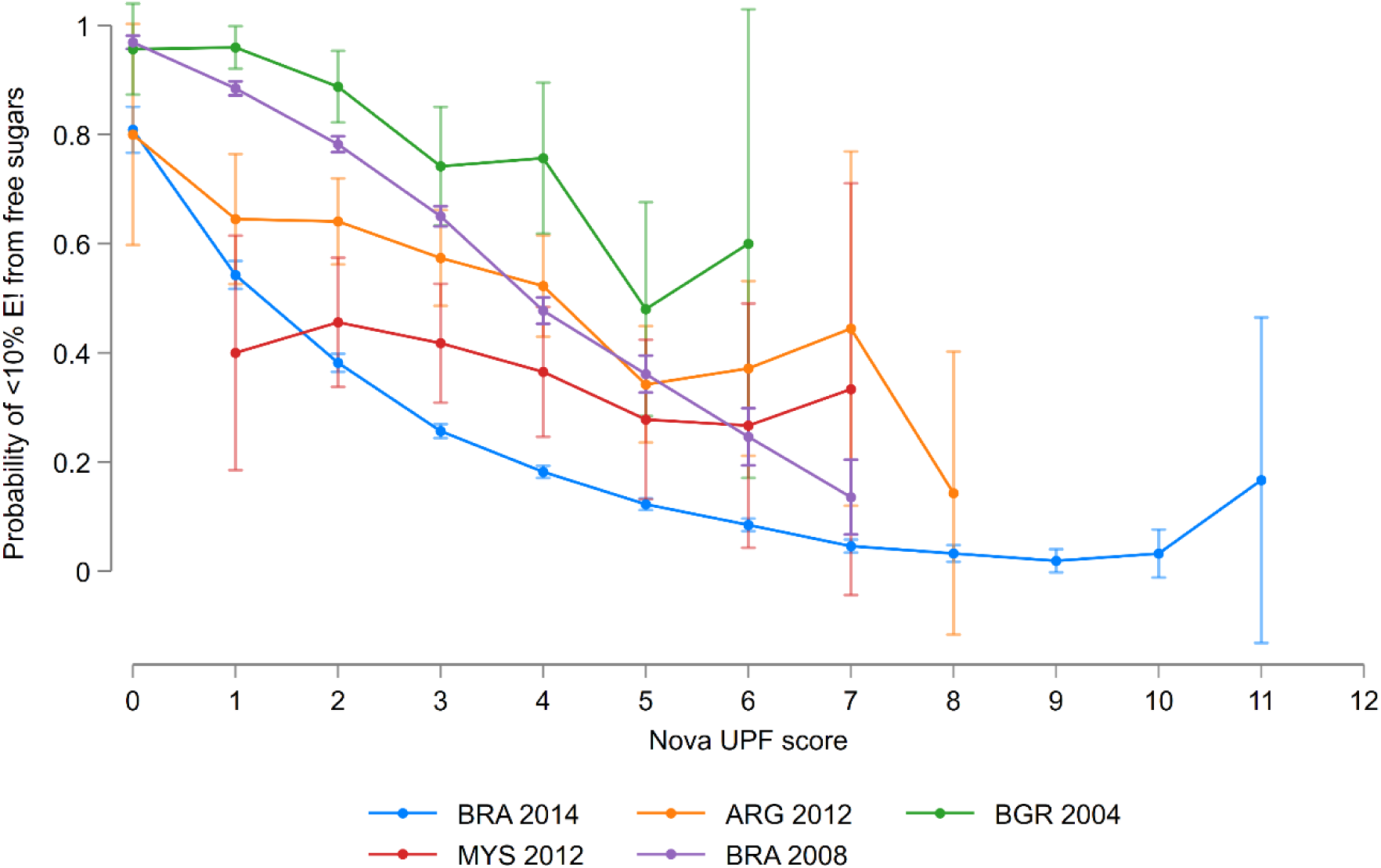
Relationship between the probability of <10% of dietary energy intake (E!) from free sugars and Nova ultra-processed food (UPF) score among non-pregnant females (15–49 years) from five survey in four upper-middle income countries. Error bars represent the 95% confidence intervals around the predicted mean probabilities from logistic regression models. ARG, Argentina; BGR, Bulgaria; BRA, Brazil; MYS, Malaysia.

However, additional analyses indicated that non-consumption of sweet foods or sweet beverages was most strongly associated with achieving <10% energy from free sugars [OR (95% CI): 5.35 (5.05, 5.66)] (**Figure 3**; **Table S11**), and this indicator also showed the most acceptable test characteristics in UMIC (**Figure S8**; **Table S9**).

**Figure 3.**
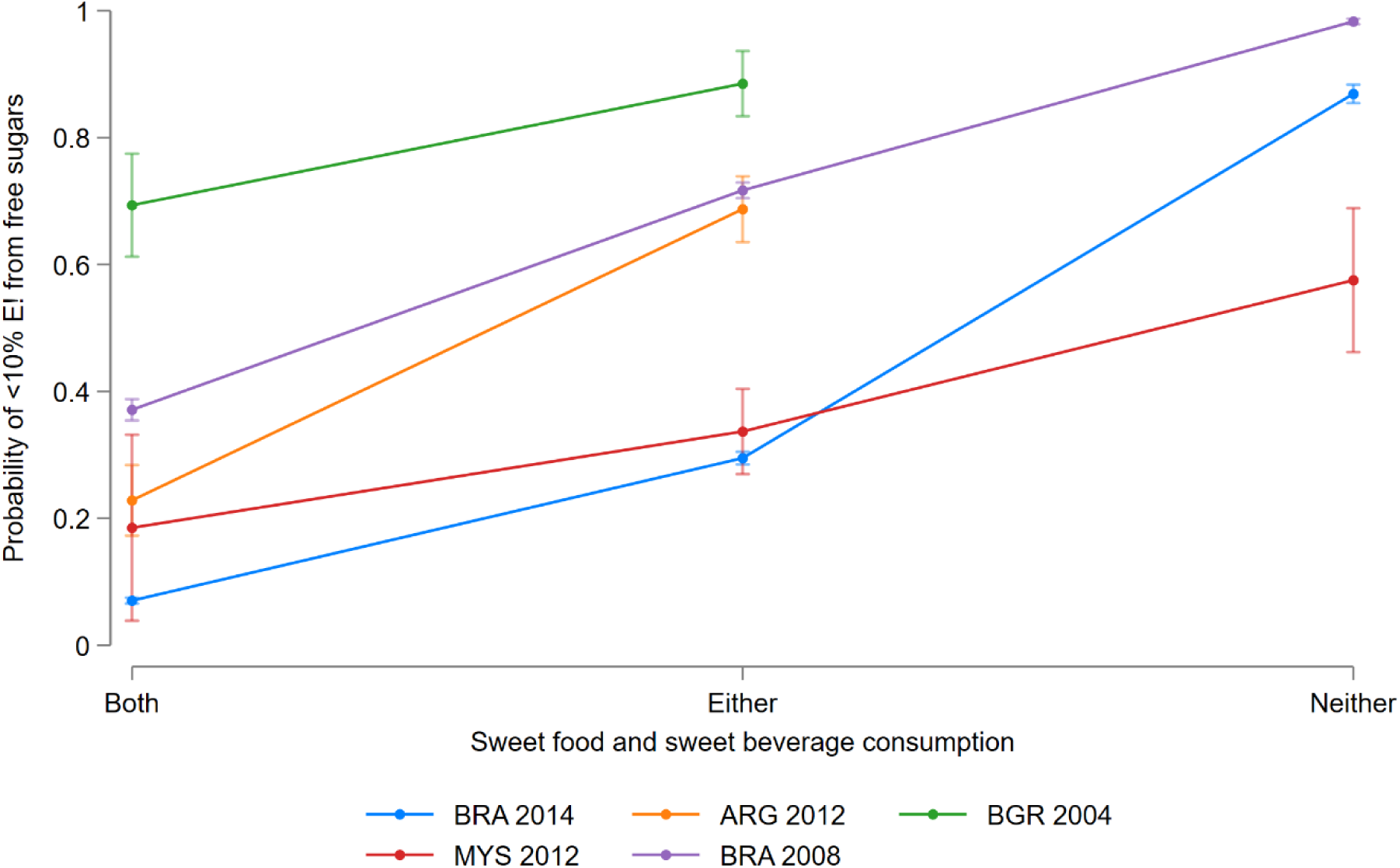
Relationship between the probability of <10% of dietary energy intake (E!) from free sugars and sweet food and sweet beverage consumption among non-pregnant females (15–49 years) from five survey in four upper-middle income countries. Error bars represent the 95% confidence intervals around the predicted mean probabilities from logistic regression models. ARG, Argentina; BGR, Bulgaria; BRA, Brazil; MYS, Malaysia.

Lower Nova UPF score was most strongly associated with lesser % energy from UPF, and the rank correlation between the 44 survey means of Nova UPF score and % energy from UPF was strong. In addition, FGDS, GDQS+, and NCD-Protect score were strongly associated with consuming ≥25 g/day of fibre; nevertheless, the strength of relationship was stronger for GDQS+ in LMIC and UMIC, and the 37 survey means of GDQS+ were most strongly rank correlated with the % reaching the guideline for fibre (**Tables S6 and S10**).

#### Nutrient adequacy

Higher FGDS was strongly associated with a higher MAR of six [*β* of 1-SD increment (95% CI): ∼11 percentage points (pp) (9, 12)] or 11 micronutrients, and each individual micronutrient across contexts (**Figure4; Tables S5-6 and S12**). Furthermore, greater FGDS was associated with a higher probability of an MAR ≥0.60 or ≥0.70 (**Figure S9**), test characteristics for FGDS ≥5 were acceptable across World Bank income classifications (**Figure S10**; **Table S13**), and the rank correlation between the 22 survey means of FGDS and MAR of six micronutrients were also strong (*ρ*=0.79) (**Figure S11**; **Tables S8 and S10**). For MAR of six but not 11 micronutrients, marginally weaker although statistically comparable findings were observed for GDQS+ across settings (**Tables S5-6**). Both the test characteristics for an indicator threshold and rank correlations across surveys were, however, weaker for GDQS+ as compared to FGDS (**Tables S8, S10, and S13**).

Likewise, greater NCD-Protect score was strongly associated with a higher probability of consuming ≥400 g/day of fruits and vegetables across World Bank country income classifications [OR of 1-SD increment (95% CI): 2.56 (2.40, 2.72) in LMIC to 3.01 (2.90, 3.13) in HIC] (**Figure 5; Table S5**). No uniform indicator threshold could be proposed across settings, however, as test characteristics were only acceptable for NCD-Protect scores ≥4 and ≥3 in LIC and UMIC, respectively (**Figure S12**; **Table S13**). Moreover, the rank correlation between the 44 survey means of NCD-Protect score and % reaching the guideline for fruits and vegetable intake was moderate (*ρ*=0.56) (**Figure S13**). FGDS performed statistically equivalently to NCD-Protect score across World Bank country income classifications (**Table S5)**, whereas ≥5 and ≥6 food group thresholds showed more acceptable test characteristics in LMIC and HIC, respectively, as compared to NCD-Protect score (**Figure S14**). In addition, mean FGDS showed the strongest rank correlation with % achieving the fruits and vegetables intake guideline across surveys (*ρ*=0.66) (**Figure S15**; **Table S8**).

**Figure 4.**
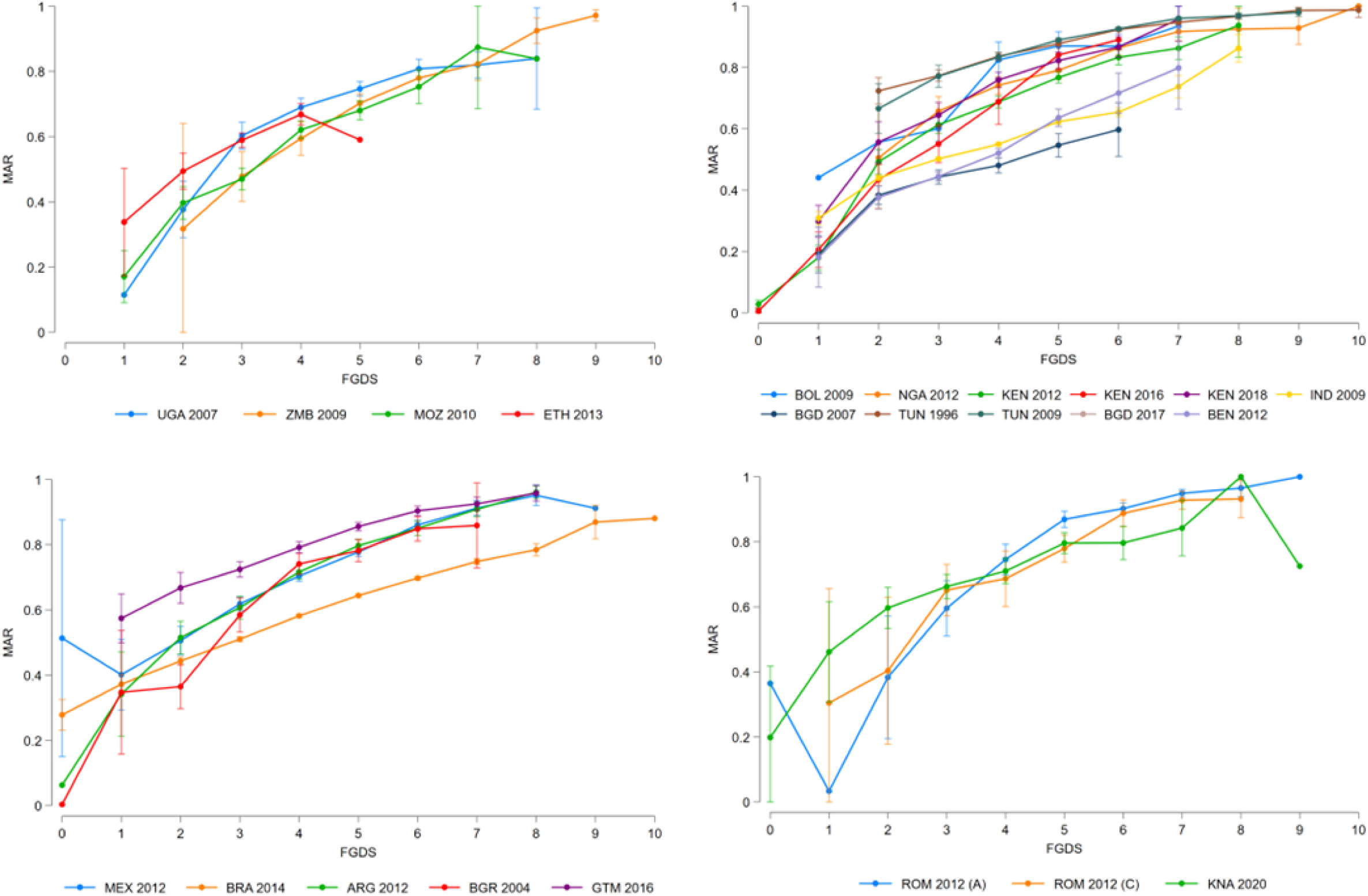
Relationship between the mean adequacy ratio (MAR) of six micronutrients (vitamin A, vitamin C, folate, calcium, iron, and zinc) and food group diversity score (FGDS) among non-pregnant females (15–49 years) in low (top left), lower-middle (top right), upper-middle (bottom left), and high income countries (bottom right). Error bars represent the 95% confidence intervals around the predicted means from linear regression models. ARG, Republic of Argentina; BEN, Republic of Benin; BFA, Burkina Faso; BGD, People’s Republic of Bangladesh; BGR, Republic of Bulgaria; BOL, Plurinational State of Bolivia; BRA, Federative Republic of Brazil; CPV, Republic of Cabo Verde; ETH, Federal Democratic Republic of Ethiopia; FJI, Republic of Fiji; GTM, Republic of Guatemala; HIC, IDN, Republic of Indonesia; IND, Republic of India; ITA, Republic of Italy; KEN, Republic of Kenya; KNA, Saint Kitts and Nevis; LAO, Lao People’s Democratic Republic; MEX, United Mexican States; MOZ, Republic of Mozambique; MYS, Malaysia; NGA, Federal Republic of Nigeria; PHL, Republic of the Philippines; ROM, Romania; TUN, Tunisian Republic; TZA, United Republic of Tanzania; UGA, Republic of Uganda; ZMB, Republic of Zambia.

**Figure 5.**
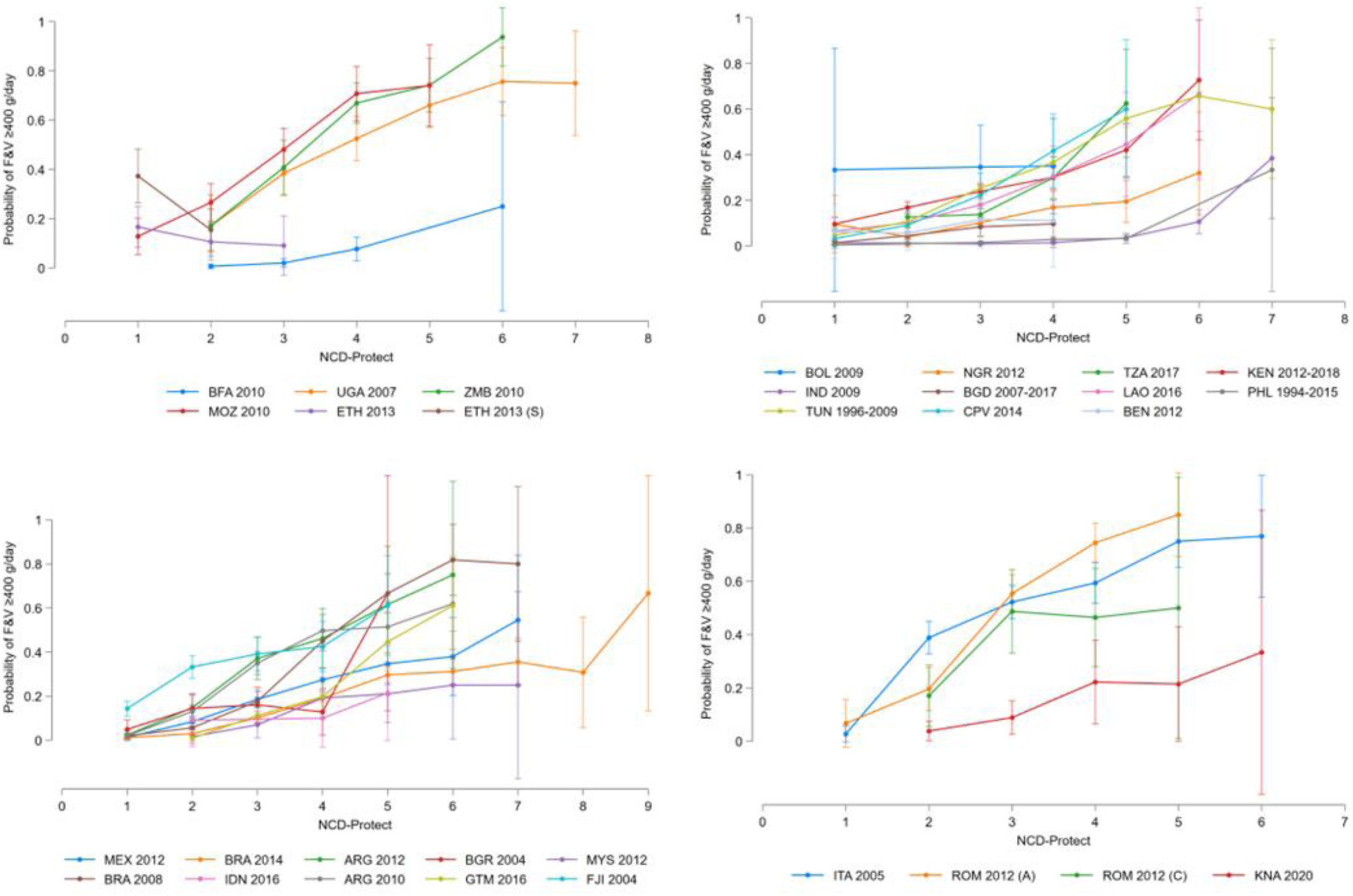
Relationship between the probability of consuming at least 400 grams per day of fruits and vegetables (F&V) and non-communicable disease (NCD)-Protect score among non-pregnant females (15–49 years) in low (top left), lower-middle (top right), upper-middle (bottom left), and high income countries (bottom right). Error bars represent the 95% confidence intervals around the predicted mean probabilities from logistic regression models. ARG, Republic of Argentina; BEN, Republic of Benin; BFA, Burkina Faso; BGD, People’s Republic of Bangladesh; BGR, Republic of Bulgaria; BOL, Plurinational State of Bolivia; BRA, Federative Republic of Brazil; CPV, Republic of Cabo Verde; ETH, Federal Democratic Republic of Ethiopia; FJI, Republic of Fiji; GTM, Republic of Guatemala; HIC, IDN, Republic of Indonesia; IND, Republic of India; ITA, Republic of Italy; KEN, Republic of Kenya; KNA, Saint Kitts and Nevis; LAO, Lao People’s Democratic Republic; MEX, United Mexican States; MOZ, Republic of Mozambique; MYS, Malaysia; NGA, Federal Republic of Nigeria; PHL, Republic of the Philippines; ROM, Romania; TUN, Tunisian Republic; TZA, United Republic of Tanzania; UGA, Republic of Uganda; ZMB, Republic of Zambia.

Sensitivity analyses showed that FGDS and NCD-Protect score were more strongly associated with absolute fruits and vegetables intake than GDQS+ in LIC, LMIC, and UMIC (data not shown). Moreover, additional analyses found that higher FV-GDR score was statistically less strongly associated with MAR than FGDS in UMIC, but most strongly associated with the likelihood of consuming ≥400 g/day of fruits and vegetables across settings (**Table S14**).

Lastly, higher FGDS and GDQS+ were consistently most strongly associated with consuming ≥3,510 mg/day of potassium across World Bank country income classifications, but both measures were weakly rank correlated with % achieving the guideline for potassium across the 26 available surveys (**Table S6**).

## Discussion

Our secondary analyses of open access quantitative 24-HR data collected from 77,188 non-pregnant females in 44 surveys across 27 countries led to five key findings. First, a simple indicator reflecting non-consumption of either sweet foods or sweet beverages, on a given day, outperformed other measures for better adherence to WHO guidelines for free sugar intake. Second, higher FGDS was robustly associated with greater micronutrient adequacy and higher probabilities of reaching WHO guidelines for fruits and vegetables and potassium intakes across World Bank country income classifications. Third, a uniform FGDS ≥5, in other words Minimum Dietary Diversity for Women, continues to be the most suitable indicator for monitoring minimally acceptable levels of micronutrient adequacy at population-level, while also showing promise to predict consumption of ≥400 g/day of fruits and vegetables globally. Fourth, while higher GDQS+ most strongly associated with reaching the WHO guideline for fibre intake across contexts, the simpler FGDS and NCD-Protect score showed comparable predictive accuracy to GDQS+ in LIC and HIC. Fifth, sweet foods and sweet beverages consumption, FGDS, and NCD-Protect score can all be collected concurrently using the lowest-burden data collection methods, namely the FGDS module and DQQ – in contrast to the semi-quantitative data required for GDQS+.

Similar to our results, a cross-sectional study among Colombian females aged 14-20 years reported that a subgroup increase in the Nova UPF score was associated with higher intakes of sodium [*β* (95% CI): 212 mg/day (163-262)] and greater % energy intake from saturated fatty acids [*β* (95% CI): 0.3 pp (0.1-0.5), respectively] (25). Higher intakes of nutrients to moderate do not always translate to clinically meaningful findings, however, if the absolute or relative intake remains below quantitative reference measures of dietary intake, such as <2,000 mg/day of sodium (15) and <10% energy from saturated fatty acids (37). Furthermore, among females and males aged ≥15 years in Brazil and the United States, higher NCD-Risk score was moderately negatively rank correlated with adherence to recommendations for food groups and nutrients to moderate, namely, total fat, saturated fatty acids, sodium, free sugars, processed meat, and unprocessed red meat (23). HDMI did not include the World Cancer Research Fund and American Institute for Cancer Research recommendation of consuming ≤350-500 g/week of unprocessed red meat as a reference indicator of dietary intake, as available quantitative dietary intake data covered the previous 24-hours only (38). To clarify, consuming >71 g/day of unprocessed red meat, often less than a portion size, on any given day, does not infer that an individual fails to adhere to the weekly intake recommendation.

Likewise to our findings, FGDS has been shown to be equivalent to and often outperform GDQS+ for micronutrient adequacy, for which nutrient intakes, anthropometry, and clinical biomarkers of undernutrition have served as proxies in sub-Saharan African countries (39,40), Mexico (41,42), China (43), India (44), and Brazil (45). The available evidence suggests that the more disaggregated and differentially weighted food groups and intake quantity thresholds used to construct GDQS+ do not provide greater predictive accuracy than the equally weighted non-quantitative count of food groups used for FGDS. Furthermore, analogous to our findings, higher FV-GDR score was correlated with greater absolute intakes of fruits and vegetables in Nigeria and Viet Nam (35). While our analyses indicated that FV-GDR score was more strongly associated with the probability of consuming ≥400 g/day of fruits and vegetables, as compared to FGDS and NCD-Protect score, HDMI considered its narrower focus, which excludes other nutritious food groups, such as pulses, nuts and seeds, and animal-source foods, insufficient for global monitoring of the characteristic nutrient adequacy. Nevertheless, metrics based on a single or limited set of food groups, such as FV-GDR, likely provide more granular insights into diets that are more actionable for policy at national level.

In parallel to our results across World Bank country income classifications, previous multi-country studies among adolescents and adults of both sexes have reported that an FGDS ≥5, widely referred to as Minimum Dietary Diversity for Women (MDD-W) (46), adequately predicts minimally acceptable levels of micronutrient adequacy at population-level as measured by a MAR or a mean probability of adequacy ≥0.60 or ≥0.70 (33,46–49). Our study also indicated that higher mean FGDS, or % reaching MDD-W, reflected higher values of reference metrics of nutrient adequacy across surveys; therefore, a uniform global FGDS indicator threshold might be sufficient to differentiate averages or prevalence of nutrient and food group values from reference metrics of nutrient adequacy. Moreover, our observation that FGDS and NCD-Protect score are comparable predictors of adherence to WHO guidelines for fruits and vegetables, fibre, and potassium intakes is supported by another study indicating high rank correlations between these two measures in Brazil and the United States (23). The consistent positive associations between greater FGDS, GDQS+, and NCD-Protect score and quantitative reference metrics of nutrient adequacy are also in line with a recent study indicating strong agreement and rank correlations between these measures among non-pregnant females in 21 countries (11). Therefore, in combination with our epidemiological findings, construct validity for FGDS, GDQS+, and NCD-Protect score across contexts is highly plausible. Nonetheless, collecting data and reporting statistics on each of these measures is redundant, and the selection of a minimum set of metrics of a healthy diet must additionally be guided by the validity and feasibility of their data collection methods.

Previous research has demonstrated that the non-quantitative open or list-based FDGS module and DQQ are sufficiently accurate in obtaining food group consumption data, as compared to reference measures such as weighed food records (50,51) and quantitative 24-HRs (35,52,53). Furthermore, due to their operational simplicity, the FGDS module and DQQ have already been integrated into multi-topic surveys, such as the Demographic and Health Survey, Gallup World Poll, and Living Standards Measurement Study, Multiple Indicator Cluster Surveys, and national nutrition surveys – which is key to ensuring a critical mass of timely nationally representative data for global monitoring. The coverage and frequency of national dietary intake data being collected, using either the FGDS module or DQQ, is expected to increase in the short term, following the “Prevalence of minimum dietary diversity, by population group” being accepted as an additional SDG 2 indicator during the 2025 Comprehensive Review (54).

In contrast, while the GDQS App has been shown to maintain predictive capacity for nutrient intakes, in comparison with quantitative 24-HRs in Thailand (55), there is limited evidence to support that integration of a standalone dietary assessment application, and portion size estimation tool, is feasible for time-and resource-constrained multi-topic surveys (56). Moreover, the requirement for individuals to visually estimate volumes of food groups consumed, using 10 three-dimensional cubes, further complicates the assessment, especially when telephone-based interviewing is used. In addition, HDMI deemed that the Nova UPF screener was neither comprehensive enough, as moderation goes beyond UPF, nor standardized sufficiently across countries for surveillance purposes. To illustrate, available Nova UPF screeners range from 23 through to 27 UPF subgroups to moderate in Brazil (24) and Ecuador (57), respectively. Furthermore, validation studies of the Nova UPF screener against % energy from UPF are somewhat tautological in nature when assessing the characteristic moderation, or put simply, the independent and dependent variables are linked in a way that is redundant or circular (24,58).

A call-to-action on global surveillance of diets is not newfound (10,59). Prior proposals were to use widely available data on food and energy availability from FAO Food Balance Sheets and household-level apparent food consumption of, for example, UPF from Household Consumption and Expenditure Surveys (60). In contrast, HDMI aimed to build consensus on a suite of valid measures and indicators, such as sweet food or sweet beverage consumption and FGDS, that can be recurrently collected across settings using low-burden data collection methods at the individual-level only. Not only do these data better reflect actual diets at national-level, when surveys are adequately powered, they also allow for assessments of differences across population subgroups within countries, which is essential for targeted and equitable nutrition actions (61). High-frequency global and national data collection platforms, such as the Gallup World Poll and Multiple Indicator Cluster Surveys, and standardized data collection tools, such as the DQQ, can help complement quantitative dietary intake data, which may be less readily available, to monitor adherence to healthy diets and ensure timely action in the context of rapid shifts in diets worldwide.

Our study has several limitations. First, quantitative 24-HR data on FAO/WHO GIFT are heterogeneous in terms of year and season of data collection, sample sizes and weighting, national representativeness, 24-HR methodology, and prevalence of over-and underreporting. Hence, our pooled regression estimates by World Bank country income classifications should be interpreted with caution. Second, our study included only four surveys from high income countries available on FAO/WHO GIFT and was restricted to non-pregnant females. Third, no standalone low-burden data collection methods, such as the DQQ or GDQS App, were used to measure food group consumption, resulting in the best-case scenario to discover associations between measures of a healthy diet and quantitative reference metrics of dietary intake (11). Fourth, self-reported and observed dietary intake data are prone to differential respondent biases and measurement error, stemming from imperfect food composition data and misestimation of dietary components, such as discretionary sodium and free sugar intakes (62). Future multi-country studies should therefore assess the relationships between measures of a healthy diet and nutritional biomarkers, such as urinary sodium (63), fructose, and sucrose (64). Fifth, single 24-HRs tend to inflate the tails of the intake distribution; thus, survey estimates of metrics of a healthy diet should not be interpreted as the usual population-level value (7). Lastly, we were unable to accurately determine whether dietary energy intake was insufficient or excessive in comparison to total energy expenditure, which is particularly relevant for macronutrient balance, due to limited anthropometric and physical activity data (65–67) and lack of data on body composition (68).

Our study also has several strengths. First, our multi-country analyses covered a large sample of diets from 77,118 females across 27 countries covering all World Bank country income classifications. Second, all datasets on FAO/WHO GIFT were harmonized with the EFSA FoodEx2 food classification and description system. This harmonization was aimed at enhancing the consistency and reliability of 24-HRs across contexts (19). Third, the recoding of FoodEx2 base terms to food (sub)groups was conducted independently by at least three experts at FAO, resulting in a high level of coding accuracy. Fourth, our comparative epidemiological analyses assessed the construct validity of measures of a healthy diet within surveys and the cross-context equivalence of observed relationships with reference metrics of dietary intake; the latter has oftentimes been overlooked but is essential for global monitoring purposes (69).

In conclusion, countries regularly collecting nationally representative quantitative 24-HRs, ideally with repeated measurements at least among a subsample, are strongly encouraged to continue, while other countries without such data collection in place should be encouraged to, and supported in, collecting dietary intake data using reference methods every 5 to 10 years. In addition, complementary lower-burden methods, in particular the FGDS module and DQQ, can provide much needed higher-frequency data on population-level consumption of nutritious and unhealthy foods groups. Furthermore, we established that metrics of a healthy diet, which can be derived from these non-quantitative data collection methods being implemented at large-scale, are valid across distinct context and, hence, suitable for timely global monitoring. Specifically, on any given day, non-consumption of either sweet foods or sweet beverages is an acceptable indicator for achieving the WHO guideline of <10% energy from free sugars, while higher nutritious food group diversity, measured by FGDS in particular, is strongly associated with higher micronutrient adequacy and greater probabilities of adhering to WHO guidelines of ≥400 and ≥25 g/day of fruits and vegetables and fibre, respectively, and ≥3,510 mg/day of potassium across contexts.

## Disclaimer

The views expressed in this publication are those of the authors and do not necessarily reflect the views or policies of the Food and Agriculture Organization of the United Nations (FAO) or the World Health Organization (WHO).

## Data availability

All meta-and micro-data supporting the results are publicly available on FAO/WHO Global Individual Food Consumption Data Tool (GIFT): https://www.fao.org/gift-individual-food-consumption/data/en

## Abbreviations

AUC: area under the curve
CI: confidence interval
DQQ: Diet Quality Questionnaire
EAR: estimated average requirement
EFSA: European Food Safety Authority
FAO: Food and Agriculture Organization of the United Nations
FGDS: food group diversity score
FV-GDR: fruits and vegetables-global dietary recommendations
GIFT: Global Individual Food Consumption Data Tool
GDQS+: Global Diet Quality Score Positive
GDQS–: Global Diet Quality Score Negative
HDMI: Healthy Diets Monitoring Initiative
HIC: high income country
24-HR: 24-hour recall and food record
LIC: low income country
LMIC: lower-middle income country
MAR: mean adequacy ratio
NAR: nutrient adequacy ratio
NCD: non-communicable disease
OR: odds ratio
pp: percentage points
ROC: receiver operating characteristics
SDG: Sustainable Development Goal
STROBE-nut: Strengthening the Reporting of Observational Studies in Epidemiology-Nutritional Epidemiology
UNICEF: United Nations Children’s Fund
UMIC: upper-middle income country
UPF: ultra-processed foods
WHO: World Health Organization.

## Ethical approval

All FAO/WHO GIFT data providers signed a License to Redistribute Contribution in which they warrant that: “The data included in any datasets have been collected and compiled in compliance with any legal or regulatory requirements as applicable to the Contributor” and “The data contained in the dataset(s) were collected in full informed consent of the data subject(s).” This study did not include any interaction or intervention with human subjects or include any access to non-anonymized data; hence, no approval was required from an institutional review board.

## Acknowledgments

We thank current and former FAO colleagues who supported the development, maintenance, data harmonisation, and management of FAO/WHO GIFT, namely Giovanni Luca Abblasio, Artur Aghajanyan, Pauline Allemand, Nitin Arora, Agnieszka Balcerzak, Teresa Bevere, Tamari Dakhundaridze, Rita Ferreira de Sousa, Daniela Ichim, Jin Jun Kim, Catherine Leclercq, Emiliana Mbelenga, Maria Julia Miele, Victoria Padula de Quadros, Marina Prieto Bercianos, Doris Rittenschober, Valeria Scrilatti, and Jacqueline Tereza da Silva. We are sincerely grateful to all data owners who have contributed and shared their data through FAO/WHO GIFT.

We also thank Nancy Aburto (FAO) for reviewing an earlier draft and the members of HDMI, in particular those who provided inputs to, and reviewed, the research protocol, namely Victor Aguayo (UNICEF), Francesco Branca (WHO), Elaine Borghi (WHO), Chika Hayashi (UNICEF), Vrinda Mehra (UNICEF), Lynnette Neufeld (FAO), and Kuntal Saha (WHO).

## Author contributions

The authors’ responsibilities were as follows – GTH-C: designed research; GTH-C: curated and managed data; GTH-C: analysed data and performed statistical analysis; GTH-C: wrote the paper; GTH-C and BAH: have primary responsibility for final content; and all authors: read, edited, and approved the final manuscript.

