## Supplementary materials for "Low-burden metrics for monitoring healthy diets across contexts: A multi-country validation analysis using quantitative 24-hour dietary intake data"

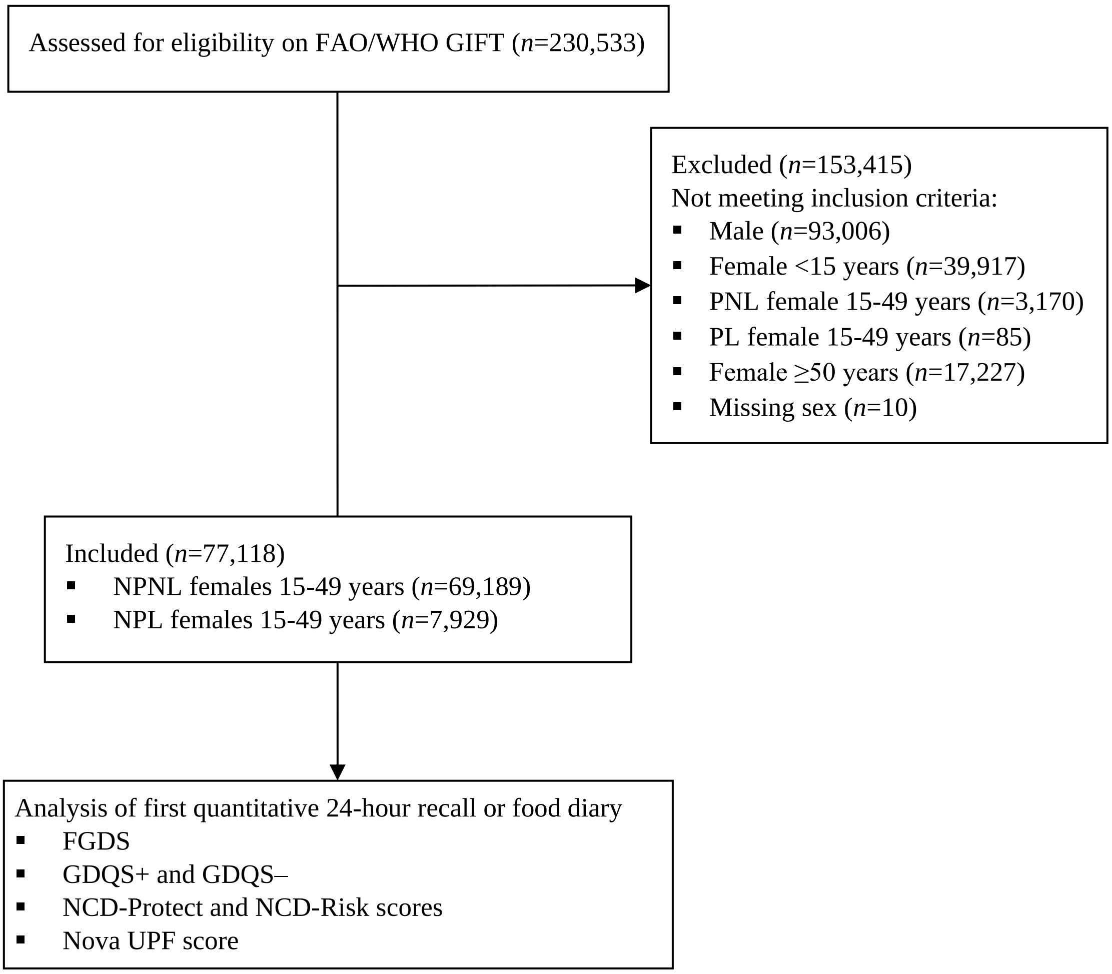


**Figure S1. Study flowchart.** FAO, Food and Agriculture Organization of the United Nations; FGDS, food group diversity score; GDQS–, Global Diet Quality Score Negative; GDQS+, Global Diet Quality Score Positive; GIFT, Global Individual Food Consumption Data Tool; NCD, non-communicable disease; NPL; non-pregnant lactating; NPNL, non-pregnant non-lactating; PL, pregnant lactating; PNL, pregnant non-lactating; UPF, ultra-processed food; WHO, World Health Organization.

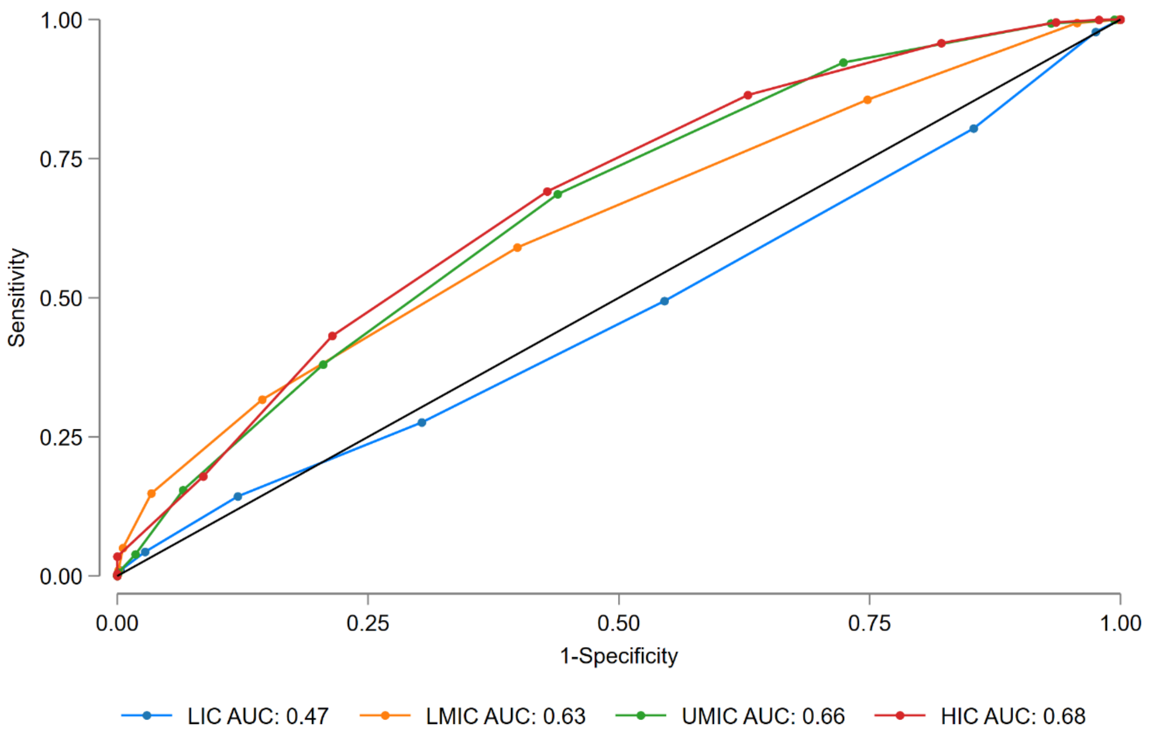


**Figure S2. Receiver operating characteristic curves of the food group diversity score (0–10 points) indicating predictions for 10-35% of dietary energy intake from protein among non-pregnant females (18–49 years), by World Bank country income classification.** AUC, area under the curve; HIC, high income country; LIC, low income country; LMIC, lower-middle income country; UMIC, upper-middle income country.

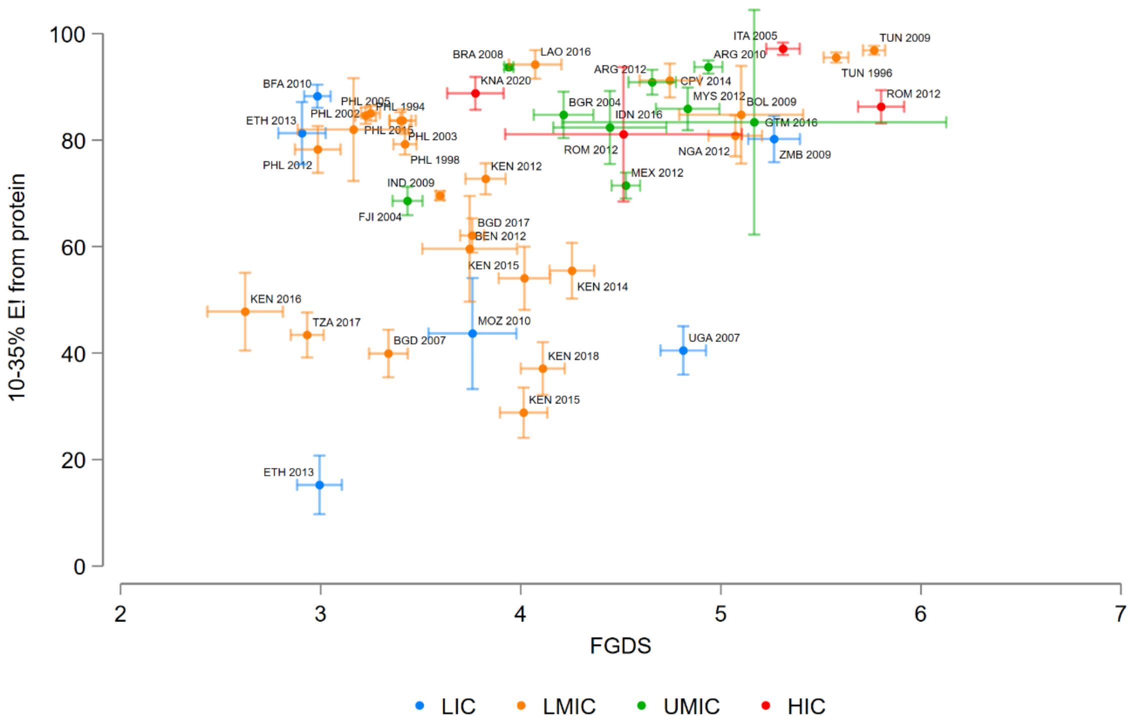


**Figure S3. Range plot with means and 95% confidence intervals of food group diversity score (FGDS) and prevalence of 10-35% of dietary energy intake (E!) from protein among non-pregnant females (18–49 years), by survey available open access on FAO/WHO GIFT.** ARG, Republic of Argentina; BEN, Republic of Benin; BFA, Burkina Faso; BGD, People's Republic of Bangladesh; BGR, Republic of Bulgaria; BOL, Plurinational State of Bolivia; BRA, Federative Republic of Brazil; CPV, Republic of Cabo Verde; ETH, Federal Democratic Republic of Ethiopia; FAO, Food and Agriculture Organization of the United Nations; FJI, Republic of Fiji; GIFT, Global Individual Food Consumption Data Tool; GTM, Republic of Guatemala; HIC, high income country; IDN, Republic of Indonesia; IND, Republic of India; ITA, Republic of Italy; KEN, Republic of Kenya; KNA, Saint Kitts and Nevis; LAO, Lao People’s Democratic Republic; LIC, low income country; LMIC, lower-middle income country; MEX, United Mexican States; MOZ, Republic of Mozambique; MYS, Malaysia; NGA, Federal Republic of Nigeria; PHL, Republic of the Philippines; ROM, Romania; TUN, Tunisian Republic; TZA, United Republic of Tanzania; UGA, Republic of Uganda; UMIC, upper-middle income country; WHO, World Health Organization; ZMB, Republic of Zambia.

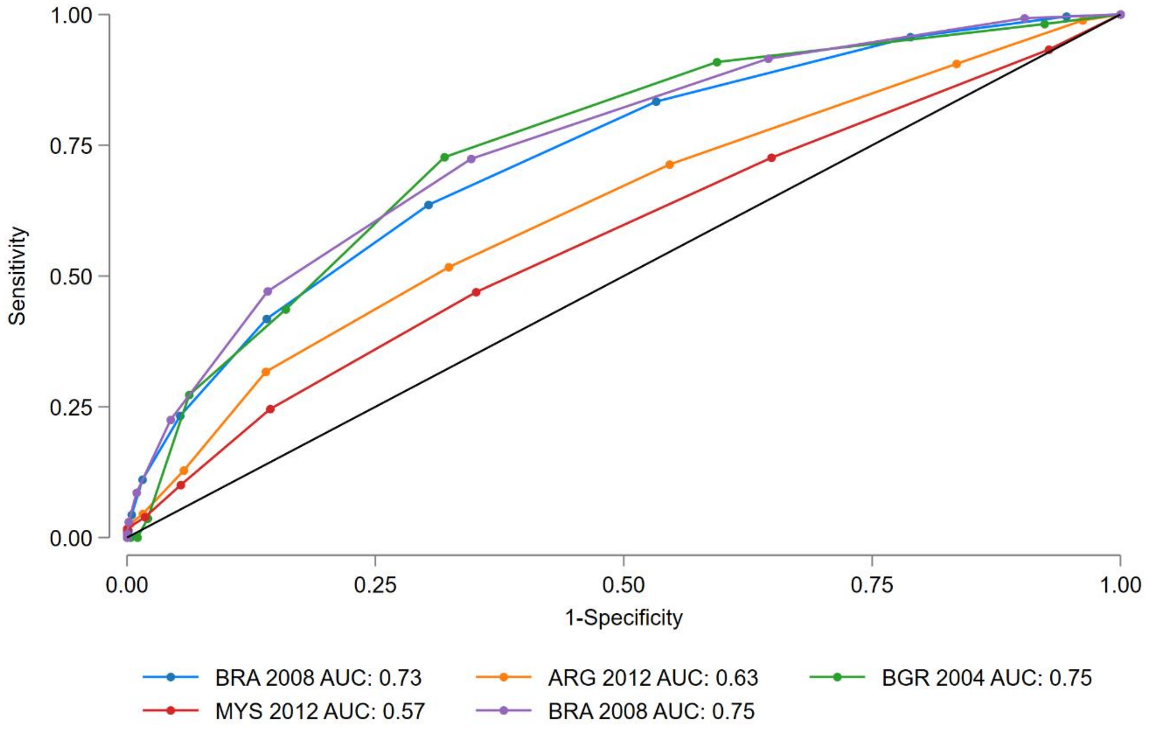


**Figure S4. Receiver operating characteristic curves of the Nova ultra-processed food score (0–23 points) indicating predictions for ≥10% of dietary energy intake from free sugars among non-pregnant females (15–49 years) from five surveys in four upper-middle income countries.** ARG, Argentina; AUC, area under the curve; BGR, Bulgaria; BRA, Brazil; MYS, Malaysia.

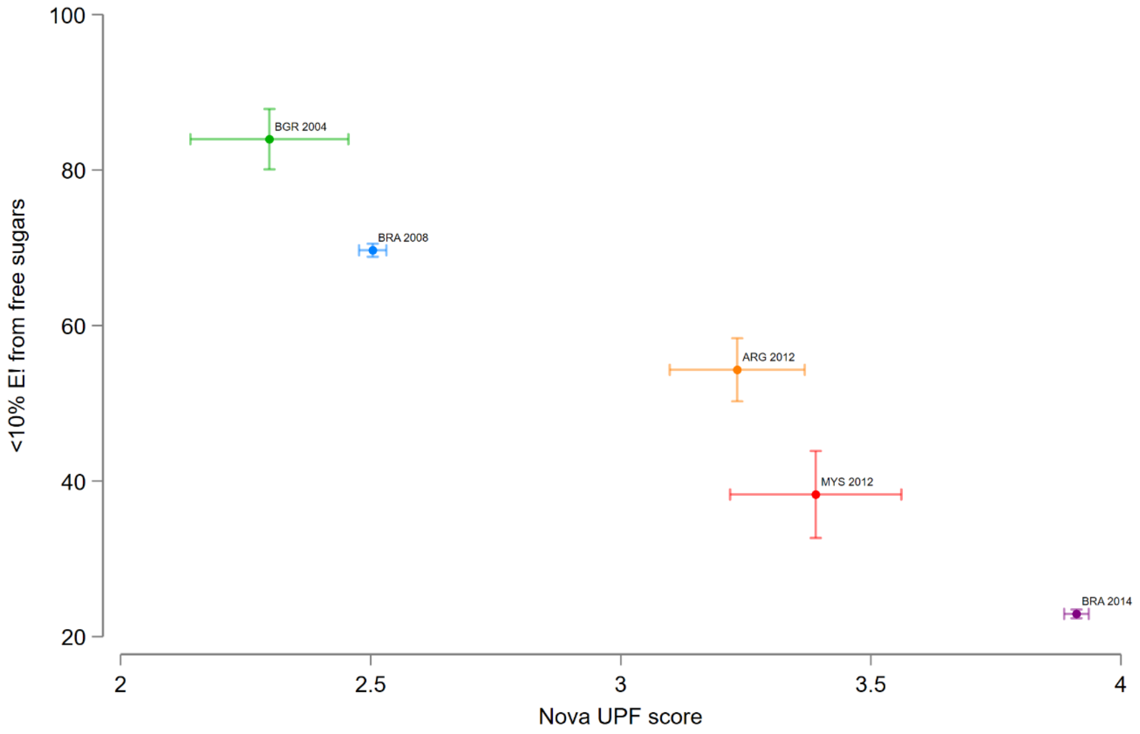


**Figure S5. Range plot with means and 95% confidence intervals of Nova ultra-processed food (UPF) score and prevalence of <10% of dietary energy intake from free sugars among non-pregnant females (15–49 years) from five surveys available open access on FAO/WHO GIFT in four upper-middle income countries.** ARG, Republic of Argentina; BGR, Republic of Bulgaria; BRA, Federative Republic of Brazil; FAO, Food and Agriculture Organization of the United Nations; GIFT, Global Individual Food Consumption Data Tool; MYS, Malaysia; WHO, World Health Organization.

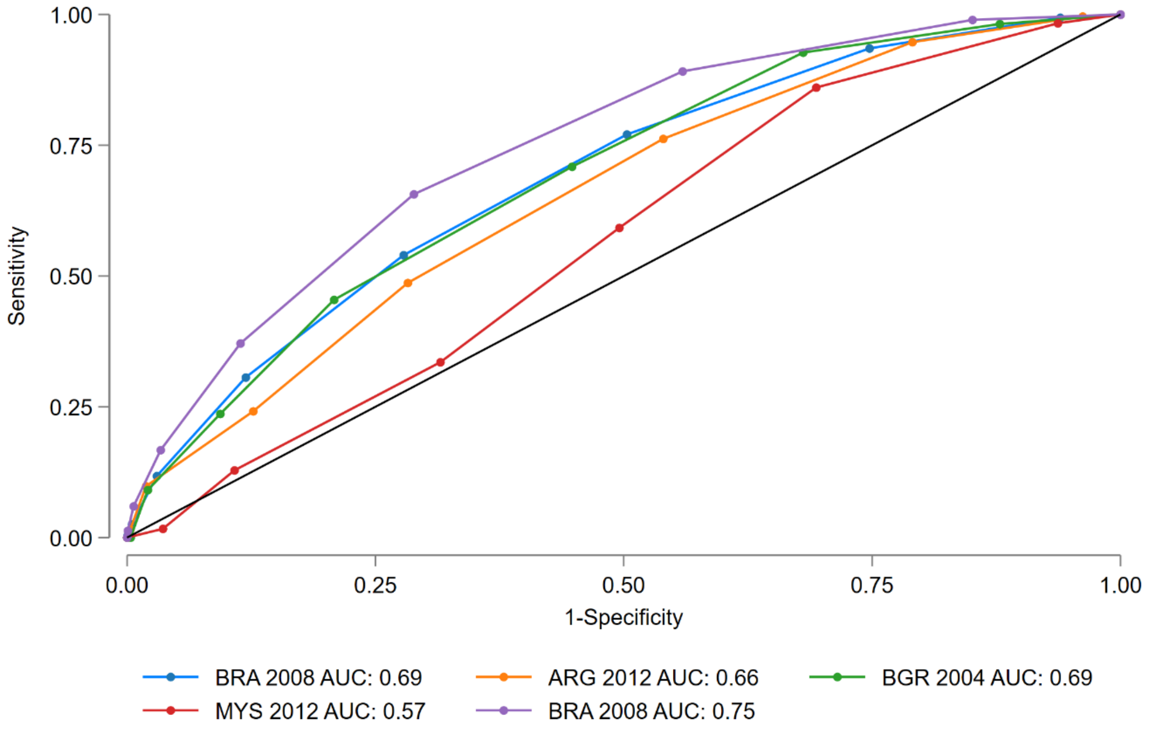


**Figure S6. Receiver operating characteristic curves of the non-communicable disease-Risk score (0–9 points) indicating predictions for ≥10% of dietary energy intake from free sugars among non-pregnant females (15–49 years) from five surveys in four upper-middle income countries.** ARG, Argentina; AUC, area under the curve; BGR, Bulgaria; BRA, Brazil; MYS, Malaysia.

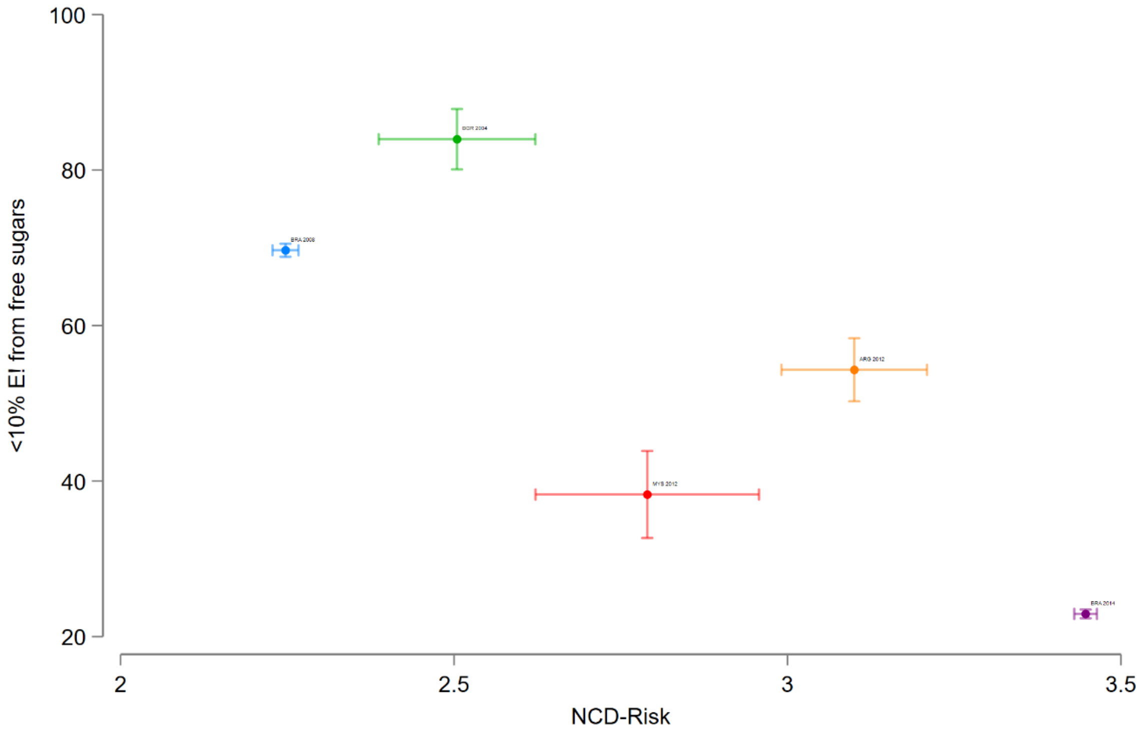


**Figure S7. Range plot with means and 95% confidence intervals of non-communicable disease (NCD)-Risk score and prevalence of <10% of dietary energy intake from free sugars among non-pregnant females (15–49 years) from five surveys available open access on FAO/WHO GIFT in four upper-middle income countries.** ARG, Republic of Argentina; BGR, Republic of Bulgaria; BRA, Federative Republic of Brazil; FAO, Food and Agriculture Organization of the United Nations; GIFT, Global Individual Food Consumption Data Tool; MYS, Malaysia; WHO, World Health Organization.

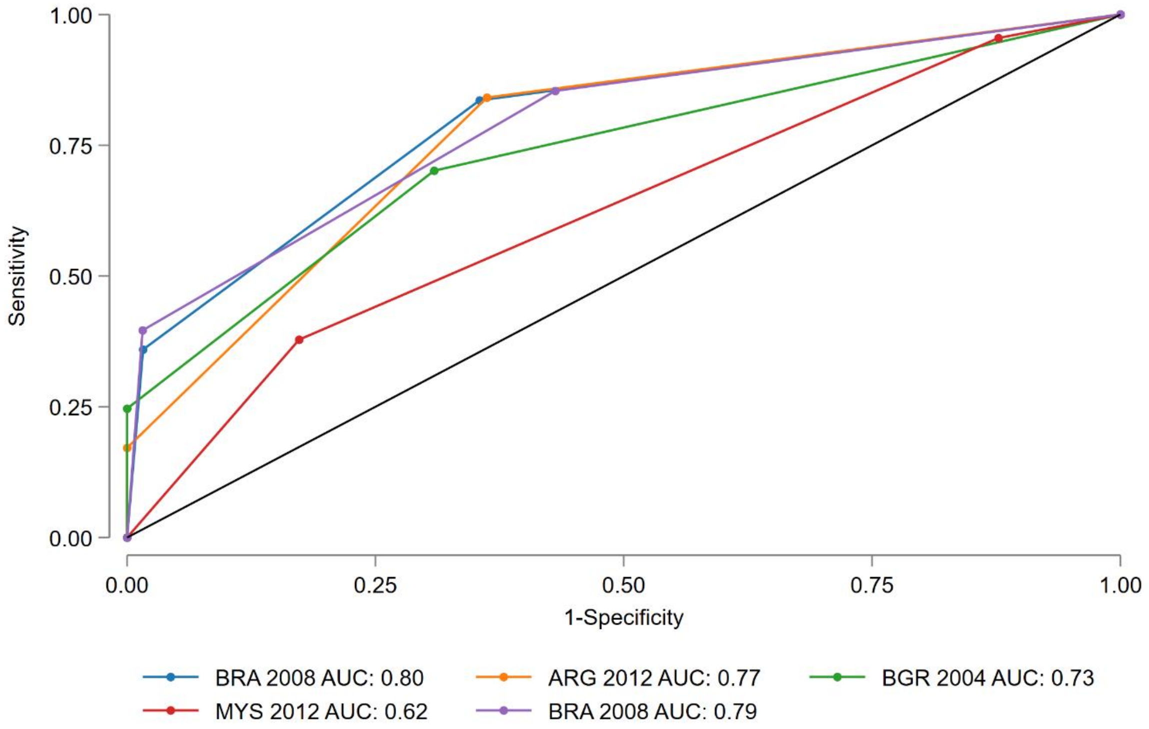


**Figure S8. Receiver operating characteristic curves of the sweet food and sweet beverage score (0–2 points) indicating predictions for <10% of dietary energy intake from free sugars among non-pregnant females (15–49 years) from five surveys in four upper-middle income countries.** ARG, Argentina; AUC, area under the curve; BGR, Bulgaria; BRA, Brazil; MYS, Malaysia.

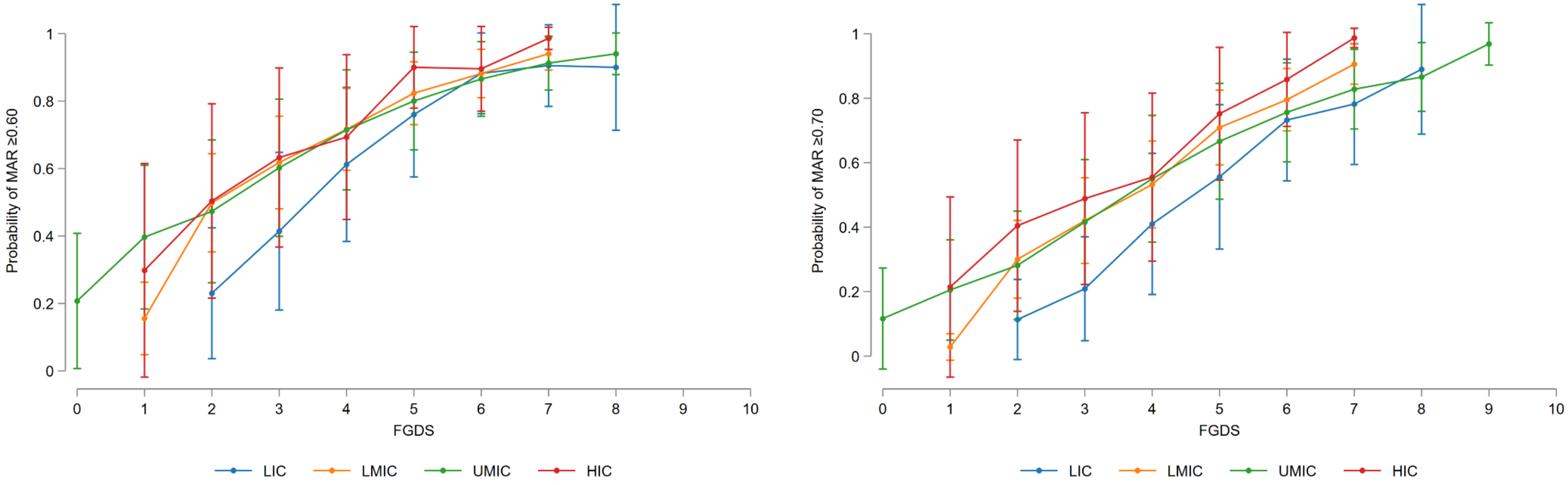


**Figure S9. Relationship between the probability of mean adequacy ratio (MAR) of six micronutrients (vitamin A, vitamin C, folate, calcium, iron, and zinc) ≥0.60 (left) or ≥0.70 and food group diversity score (FGDS) among non-pregnant females (15–49 years), by World Bank country income classification.** Error bars represent the 95% confidence intervals around the predicted mean probabilities from mixed effect logistic regression models with random intercepts for survey and random slopes for FGDS. HIC, high income country; LIC, low income country; LMIC, lower-middle income country; UMIC, upper-middle income country.

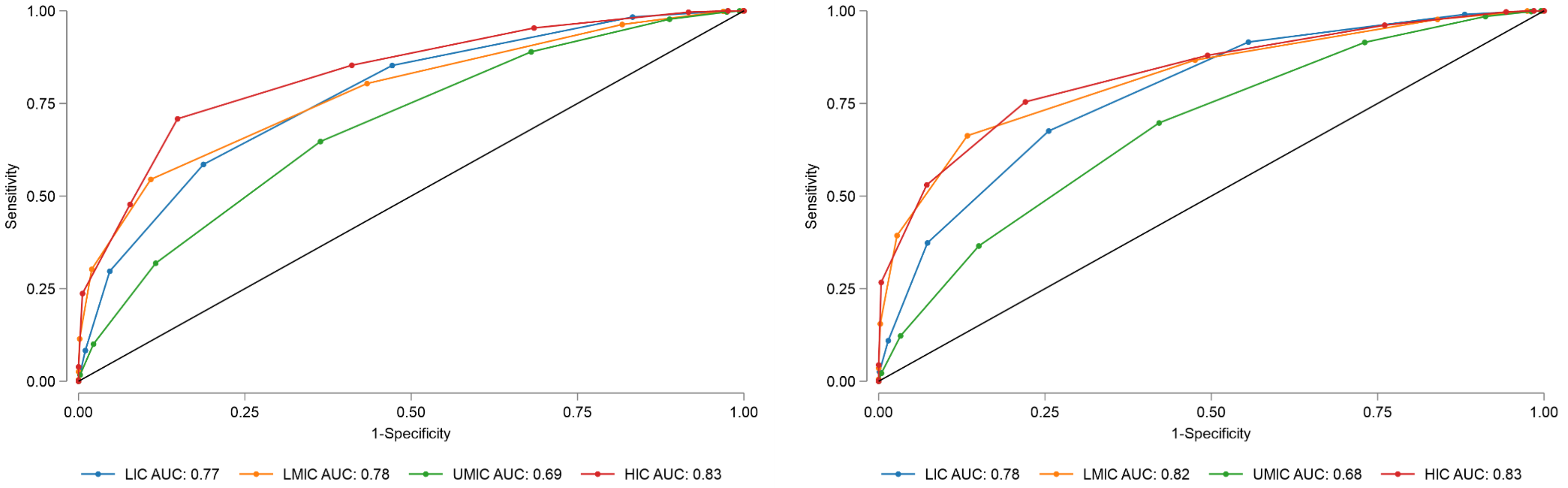


**Figure S10. Receiver operating characteristic curves of the food group diversity score (0–10 points) indicating predictions for mean adequacy ratio of six micronutrients (vitamin A, vitamin C, folate, calcium, iron, and zinc) ≥0.60 (left) or ≥0.60 (right) among non-pregnant females (15–49 years), by World Bank country income classification.** AUC, area under the curve; HIC, high income country; LIC, low income country; LMIC, lower-middle income country; UMIC, upper-middle income country.

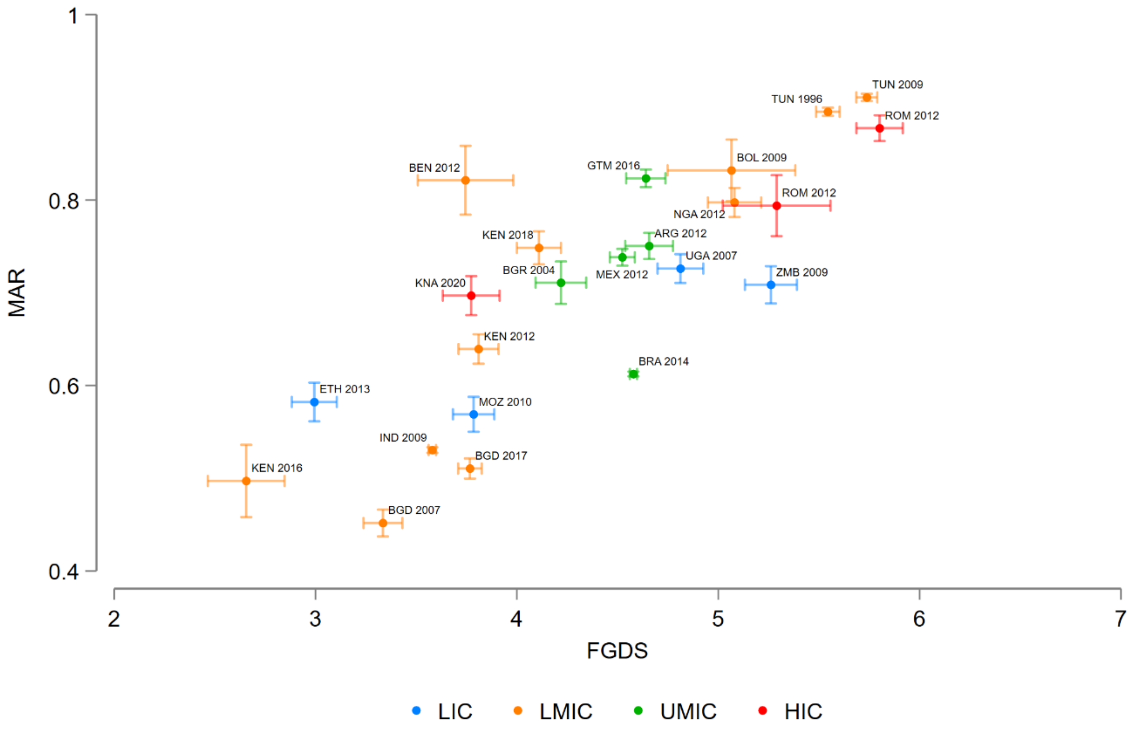


**Figure S11. Range plot with means and 95% confidence intervals of food group diversity score (FGDS) and mean adequacy ratio (MAR) of six micronutrients (vitamin A, vitamin C, folate, calcium, iron, and zinc) among non-pregnant females (15–49 years), by survey available open access on FAO/WHO GIFT.** ARG, Republic of Argentina; BEN, Republic of Benin; BGD, People's Republic of Bangladesh; BGR, Republic of Bulgaria; BOL, Plurinational State of Bolivia; BRA, Federative Republic of Brazil; ETH, Federal Democratic Republic of Ethiopia; FAO, Food and Agriculture Organization of the United Nations; GIFT, Global Individual Food Consumption Data Tool; GTM, Republic of Guatemala; HIC, high income country; IND, Republic of India; KEN, Republic of Kenya; KNA, Saint Kitts and Nevis; LIC, low income country; LMIC, lower-middle income country; MEX, United Mexican States; MOZ, Republic of Mozambique; NGA, Federal Republic of Nigeria; ROM, Romania; TUN, Tunisian Republic; UGA, Republic of Uganda; UMIC, upper-middle income country; WHO, World Health Organization; ZMB, Republic of Zambia.

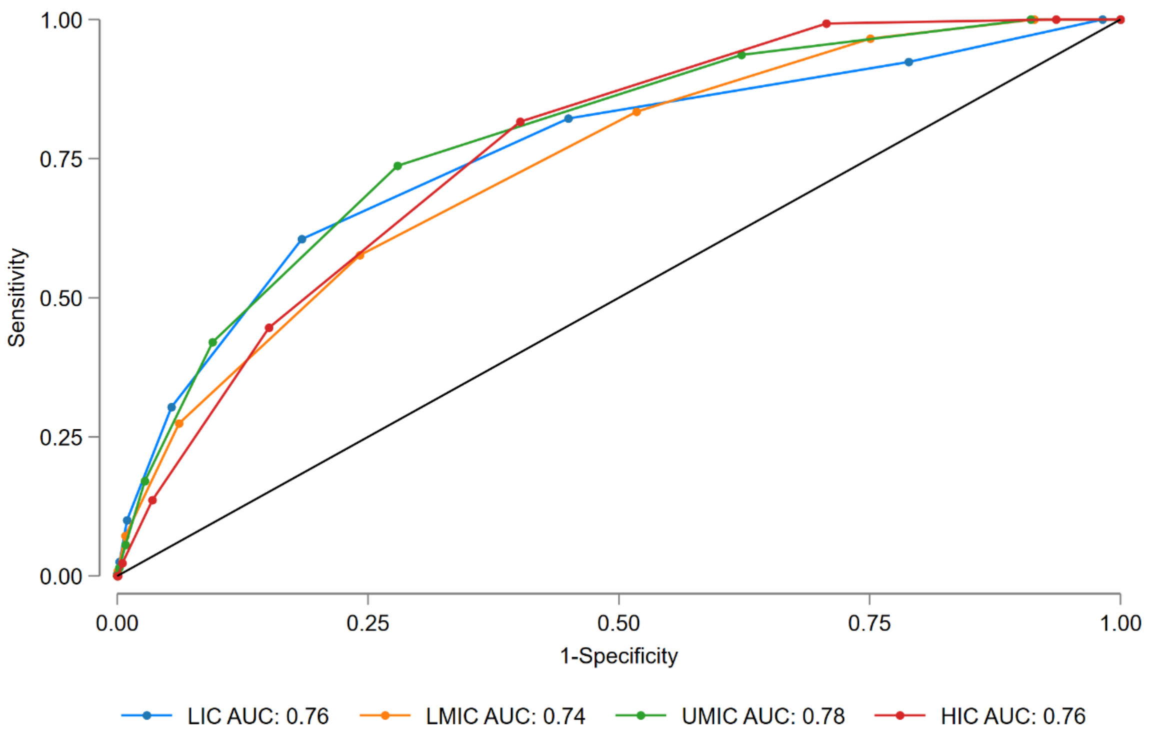


**Figure S12. Receiver operating characteristic curves of the non-communicable disease-Protect score (0–9 points) indicating predictions for ≥400 g/day of fruits and vegetables among non-pregnant females (15–49 years), by World Bank country income classification.** AUC, area under the curve; HIC, high income country; LIC, low income country; LMIC, lower-middle income country; UMIC, upper-middle income country.

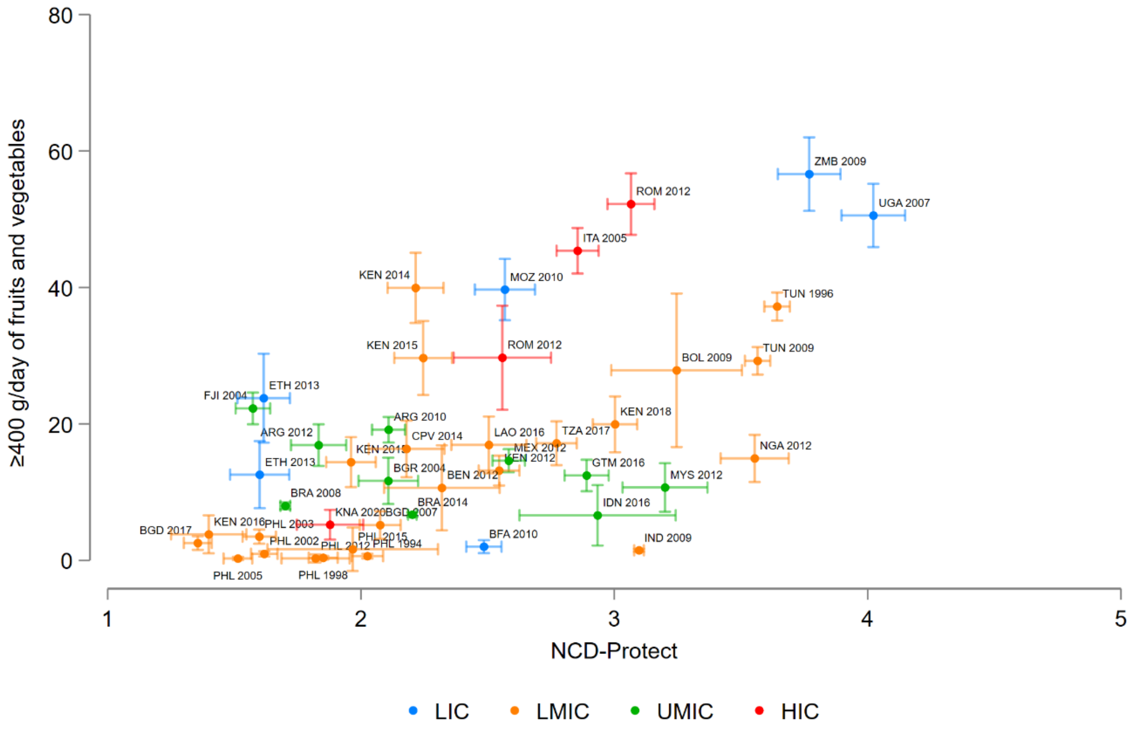


**Figure S13. Range plot with means and 95% confidence intervals of non-communicable disease (NCD)-Protect score and prevalence of ≥400 g/day of fruits and vegetables among non-pregnant females (15–49 years), by survey available open access on FAO/WHO GIFT.** ARG, Republic of Argentina; BEN, Republic of Benin; BFA, Burkina Faso; BGD, People's Republic of Bangladesh; BGR, Republic of Bulgaria; BOL, Plurinational State of Bolivia; BRA, Federative Republic of Brazil; CPV, Republic of Cabo Verde; ETH, Federal Democratic Republic of Ethiopia; FAO, Food and Agriculture Organization of the United Nations; FJI, Republic of Fiji; GIFT, Global Individual Food Consumption Data Tool; GTM, Republic of Guatemala; HIC, high income country; IDN, Republic of Indonesia; IND, Republic of India; ITA, Republic of Italy; KEN, Republic of Kenya; KNA, Saint Kitts and Nevis; LAO, Lao People’s Democratic Republic; LIC, low income country; LMIC, lower-middle income country; MEX, United Mexican States; MOZ, Republic of Mozambique; MYS, Malaysia; NGA, Federal Republic of Nigeria; PHL, Republic of the Philippines; ROM, Romania; TUN, Tunisian Republic; TZA, United Republic of Tanzania; UGA, Republic of Uganda; UMIC, upper-middle income country; WHO, World Health Organization; ZMB, Republic of Zambia.

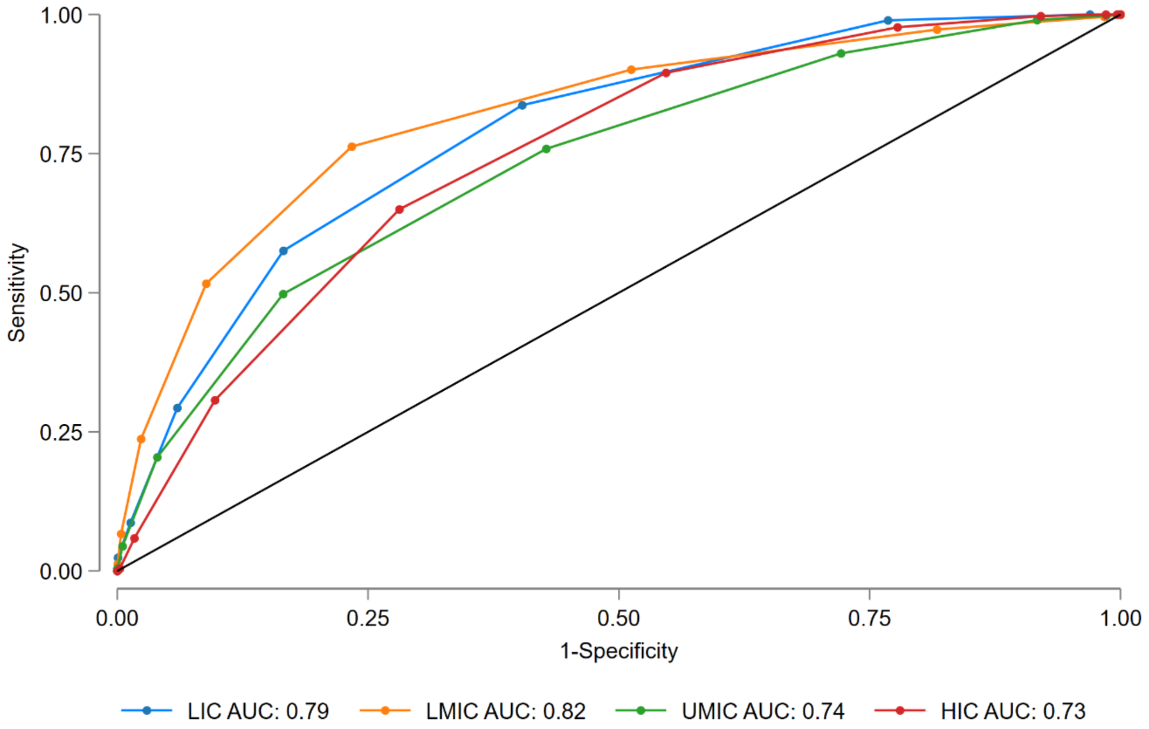


**Figure S14. Receiver operating characteristic curves of the food group diversity score (0–10 points) indicating predictions for ≥400 g/day of fruits and vegetables among non-pregnant females (15–49 years), by World Bank country income classification.** AUC, area under the curve; HIC, high income country; LIC, low income country; LMIC, lower-middle income country; UMIC, upper-middle income country.

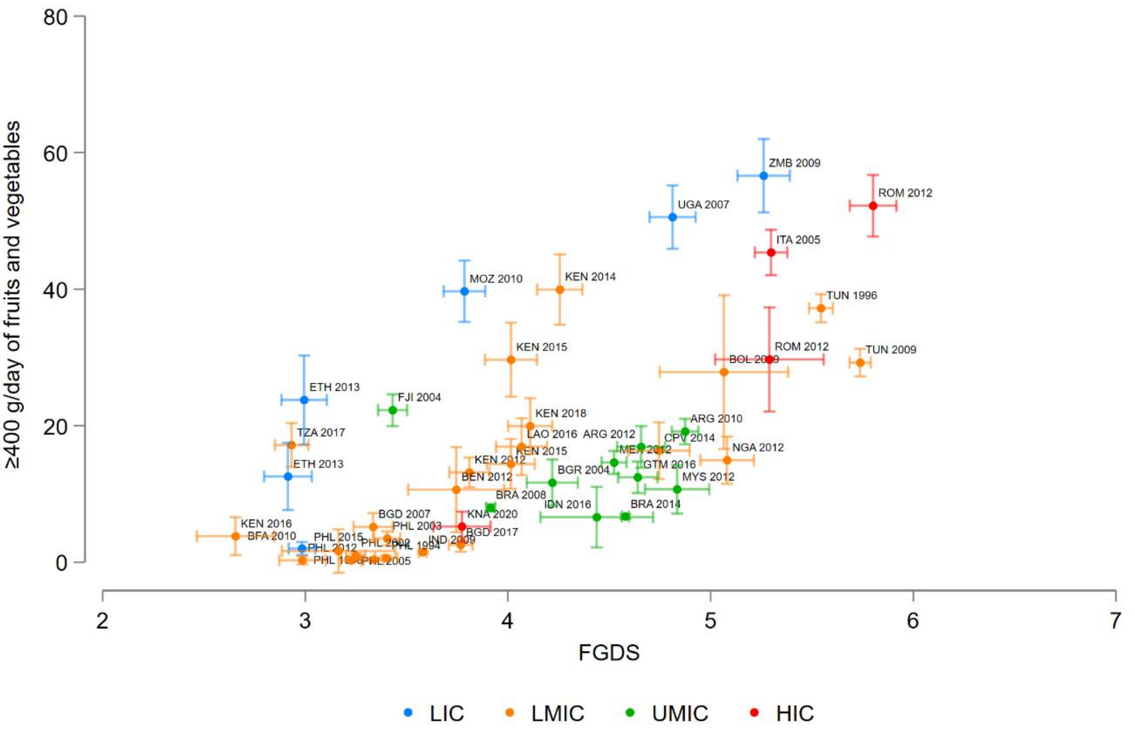


**Figure S15. Range plot with means and 95% confidence intervals of food group diversity score (FGDS) and prevalence of ≥400 g/day of fruits and vegetables among non-pregnant females (15–49 years), by survey available open access on FAO/WHO GIFT.** ARG, Republic of Argentina; BEN, Republic of Benin; BFA, Burkina Faso; BGD, People's Republic of Bangladesh; BGR, Republic of Bulgaria; BOL, Plurinational State of Bolivia; BRA, Federative Republic of Brazil; CPV, Republic of Cabo Verde; ETH, Federal Democratic Republic of Ethiopia; FAO, Food and Agriculture Organization of the United Nations; FJI, Republic of Fiji; GIFT, Global Individual Food Consumption Data Tool; GTM, Republic of Guatemala; HIC, high income country; IDN, Republic of Indonesia; IND, Republic of India; ITA, Republic of Italy; KEN, Republic of Kenya; KNA, Saint Kitts and Nevis; LAO, Lao People’s Democratic Republic; LIC, low income country; LMIC, lower-middle income country; MEX, United Mexican States; MOZ, Republic of Mozambique; MYS, Malaysia; NGA, Federal Republic of Nigeria; PHL, Republic of the Philippines; ROM, Romania; TUN, Tunisian Republic; TZA, United Republic of Tanzania; UGA, Republic of Uganda; UMIC, upper-middle income country; WHO, World Health Organization; ZMB, Republic of Zambia.

**Table S1. Surveys including non-pregnant females aged 15-49 years on FAO/WHO GIFT, by World Bank income classification**

| Survey, country and years | Sample size |
|---|---|
| **Low income** | 2,408 |
| Burkina Faso 2010 (1) | 841 |
| Ethiopia 2013 (2) | 175 |
| Ethiopia (South) 2013 (3) | 164 |
| Mozambique 2010 (4) | 456 |
| Uganda 2007 (5) | 447 |
| Zambia 2009 (6) | 325 |
| **Lower-middle income** | 32,766 |
| Bangladesh 2007-2008 (7) | 463 |
| Bangladesh 2017-2018[1] (8) | 939 |
| Benin 2012 (9) | 94 |
| Bolivia 2009-2012 (10) | 61 |
| Cabo Verde 2014 (11) | 306 |
| India 2009-2012[1] (12) | 12,598 |
| Indonesia 2016 | 121 |
| Kenya 2012 (13) | 919 |
| Kenya 2014 (Lean season) (14) | 348 |
| Kenya 2015 (15) | 273 |
| Kenya 2015 (Plenty season) (14) | 354 |
| Kenya 2016 | 183 |
| Kenya 2018 (16) | 366 |
| Lao PDR 2016-2017[1] (17) | 313 |
| Nigeria 2011 (18) | 408 |
| Philippines 1994-1996 (19) | 2,054 |
| Philippines 1998-2000 (19) | 2,286 |
| Philippines 2002-2003 (19) | 2,488 |
| Philippines 2002-2003[1] (20) | 1,205 |
| Philippines 2005 (19) | 2,107 |
| Philippines 2012-2013 (19) | 340 |
| Philippines 2015-2016 (19) | 61 |
| Tanzania 2016 (21) | 530 |
| Tunisia 1996-1997[1] (22) | 2,118 |
| Tunisia 2009-2010 (23) | 1,952 |
| **Upper-middle income** | 40,077 |
| Argentina 2011 (24) | 1,697 |
| Argentina 2012-2013 (25) | 580 |
| Brazil 2008-2009[1] (26) | 11,594 |
| Brazil 2013-2014[1] (27) | 21,739 |
| Bulgaria 2004 (28) | 343 |
| Fiji 2004 (29) | 1,230 |
| Guatemala 2016 | 787 |
| Indonesia 2016 | 121 |
| Malaysia 2012-2013 (30) | 290 |
| Mexico 2012[1] (31) | 1,696 |
| **High income** | 1,867 |
| Italy 2005-2006[1] (32) | 855 |
| Romania (Adult: 19-49 years) 2012[1] (33) | 473 |
| Romania (Child: 15-18 years) 2012[1] (34) | 138 |
| Saint-Kitts and Nevis 2020-2021 (35) | 401 |
| **Total** | 77,118 |

[1]Data from nationally representative surveys for specific population groups. FAO, Food and Agriculture Organization; GIFT, Global Individual Food consumption data Tool; PDR; People's Democratic Republic; WHO, World Health Organization.

#### Table S2. Descriptive statistics of demographic variables and healthy diet metrics among non-pregnant females aged 15-49 years, by World Bank income classification and nationally representative surveys available open access on FAO/WHO GIFT[1]

| Survey | Sample size | Age (years) | Weight (kg) | Height (cm) | BMI (kg/m²)[2] | FGDS (0-10) | MDD-W (%) | GDQS (0-49) | GDQS+ (0-32) | GDQS− (0-17) | GDQS ≥15 | GDQS ≥23 | GDR score (0-18) | GDR indicator (%) | NCD-Protect (0-9) | NCD-Risk (0-9) | Nova UPF score (0-23) |
|---|---|---|---|---|---|---|---|---|---|---|---|---|---|---|---|---|---|
| **LIC** | 2,408 | 28.3 (8.58)<br>28 (22, 35) | 53.5 (8.75)<br>52.6 (47.9, 57.8)<br>*n*=2,129 | 159 (6.89)<br>159 (154, 163)<br>*n*=2,129 | 21.5 (3.03)<br>20.9 (19.4, 22.8)<br>*n*=1,640 | 3.78 (1.29)<br>4 (3, 5) | 28.0 | 17.8 (3.66)<br>18 (15.25, 20.25) | 6.88 (3.53)<br>6.5 (4.25, 9) | 10.9 (1.44)<br>11 (10, 12) | 78.6 | 8.89 | 11.1 (1.51)<br>11 (10, 12) | 87.6 | 2.84 (1.39)<br>3 (1, 4) | 0.72 (0.81)<br>1 (0, 1) | 0.37 (0.63)<br>0 (0, 1) |
| **LMIC** | 32,766 | 31.8 (10.1)<br>32 (23, 40) | 51.9 (12.7)<br>49.7 (42.7, 58.8)<br>*n*=29,499 | 153 (6.36)<br>152 (148, 157)<br>*n*=29,502 | 22.6 (4.89)<br>21.8 (19.0, 25.5)<br>*n*=25,019 | 3.80 (1.38)<br>4 (3, 5) | 27.3 | 17.4 (3.60)<br>17.25 (15, 19.75) | 6.15 (3.22)<br>6 (4, 8.25) | 11.2 (1.68)<br>11 (10, 12) | 75.9 | 7.01 | 10.2 (1.93)<br>11 (9, 12) | 70.9 | 2.59 (1.40)<br>3 (2, 4) | 1.37 (1.17)<br>1 (1, 2) | 0.92 (1.26)<br>0 (0, 2) |
| BGD 2017 | 939 | 28.0 (8.49)<br>26 (22, 34) | 52.0 (11.0)<br>50.0 (45.0, 59.0) | 152 (5.89)<br>152 (148, 156) | 23.0 (4.24)<br>22.6 (20.0, 25.5) | 3.77 (0.92)<br>4 (3, 4) | 19.1 | 17.1 (2.6)<br>16.5 (15.25, 18.75) | 6.02 (2.38)<br>5.5 (4.25, 7.75) | 11.1 (0.89)<br>11 (10, 12) | 79.0 | 1.92 | 10.1 (1.04)<br>10 (9, 11) | 71.8 | 1.36 (0.86)<br>1 (1, 2) | 0.26 (0.50)<br>0 (0, 0) | 0.04 (0.20)<br>0 (0, 0) |
| IND 2009 | 12,598 | 30.7 (9.67)<br>30 (23, 39) | 46.6 (9.60)<br>44.7 (39.9, 51.6) | 152 (5.76)<br>152 (148, 156) | 20.5 (3.92)<br>19.8 (17.7, 22.7) | 3.58 (1.06)<br>4 (3, 4) | 16.8 | 18.0 (3.01)<br>18 (16, 20) | 6.14 (2.56)<br>6.25 (4.25, 8) | 11.9 (1.39)<br>12 (11, 13) | 85.2 | 5.97 | 11.2 (1.16)<br>11 (10, 12) | 92.7 | 3.10 (1.09)<br>3 (2, 4) | 0.93 (0.53)<br>1 (1, 1) | 0.12 (0.33)<br>0 (0, 0) |
| LAO 2016 | 313 | 30.0 (8.71)<br>28 (23, 36) | 53.7 (9.50)<br>51.9 (47.1, 58.8) | 152 (5.16)<br>152 (149, 156) | 23.4 (3.85)<br>22.9 (20.8, 25.5) | 4.07 (1.13)<br>4 (3, 5) | 35.1 | 15.9 (3.1)<br>15.5 (14, 18) | 4.98 (2.86)<br>4.5 (2.75, 7.25) | 10.9 (1.37)<br>11 (10, 12) | 60.4 | 0.96 | 9.40 (1.74)<br>9 (8, 11) | 49.8 | 2.50 (1.34)<br>2 (2, 3) | 2.11 (1.58)<br>2 (1, 3) | 0.41 (0.73)<br>0 (0, 1) |
| PHL 2003 | 1,205 | 29.8 (6.78)<br>28 (23, 34) | NA | NA | NA | 3.40 (1.15)<br>3 (3, 4) | 16.4 | 15.8 (2.73)<br>15.5 (14, 17.5) | 4.49 (2.60)<br>4.25 (2.25, 6) | 11.3 (1.11)<br>12 (11, 12) | 61.9 | 1.41 | 9.45 (1.48)<br>10 (9, 10) | 50.4 | 1.60 (1.17)<br>2 (1, 2) | 1.14 (1.06)<br>1 (0, 2) | 0.25 (0.49)<br>0 (0, 0) |
| TUN 1996[3] | 2,118 | 29.6 (9.67)<br>30 (21, 39) | 61.0 (13.3)<br>58.7 (51.0, 69.2) | 158 (6.29)<br>157 (153, 162) | 25.3 (5.29)<br>24.8 (21.4, 28.9) | 5.55 (1.35)<br>6 (5, 6) | 80.6 | 20.2 (3.38)<br>20.25 (17.75, 22.75) | 9.79 (3.39)<br>9.75 (7.5, 12.5) | 10.4 (1.34)<br>10 (9, 11) | 93.9 | 24.6 | 11.2 (1.35)<br>11 (10, 12) | 90.6 | 3.61 (1.19)<br>4 (3, 4) | 1.42 (0.79)<br>1 (1, 2) | 1.23 (0.62)<br>1 (1, 1) |
| **UMIC** | 40,077 | 22.3 (9.68)<br>17 (16, 28) | 58.9 (12.6)<br>56.6 (50.1, 65.2)<br>*n*=37,937 | 160 (6.67)<br>160 (155, 164)<br>*n*=37,936 | 25.4 (5.12)<br>24.4 (21.8, 27.9)<br>*n*=13,319 | 4.36 (1.35)<br>4 (3, 5) | 45.7 | 14.8 (3.83)<br>15 (12, 17.25) | 5.70 (3.02)<br>6 (4, 8) | 9.05 (2.52)<br>9 (7, 11) | 50.6 | 1.58 | 8.07 (1.96)<br>8 (7, 9) | 23.3 | 2.07 (1.27)<br>2 (1, 3) | 3.00 (1.63)<br>3 (2, 4) | 3.30 (1.87)<br>3 (2, 4) |
| BGR 2004[3] | 343 | 27.0 (9.96)<br>25 (18, 33) | 60.5 (11.9)<br>58.0 (52.0, 67.0) | 164 (6.81)<br>164 (160, 168) | 23.5 (4.82)<br>22.1 (20.0, 26.1) | 4.22 (1.19)<br>4 (4, 5) | 40.2 | 14.9 (3.54)<br>14.75 (12.25, 17.5) | 5.26 (2.69)<br>5 (3.25, 7.25) | 9.66 (2.54)<br>10 (8, 12) | 49.9 | 2.04 | 8.60 (1.85)<br>9 (7, 10) | 32.4 | 2.11 (1.11)<br>2 (1, 3) | 2.50 (1.55)<br>2 (1, 3) | 2.30 (1.49)<br>2 (1, 3) |
| BRA 2008[3] | 11,594 | 31.3 (9.97)<br>30 (23, 39) | 62.9 (13.0)<br>60.2 (53.2, 68.9) | 160 (6.98)<br>159 (154, 164) | 25.1 (4.93)<br>24.2 (21.7, 27.6) | 3.94 (1.21)<br>4 (3, 5) | 31.1 | 15.0 (3.67)<br>15.25 (13, 18) | 5.04 (2.63)<br>5 (4, 6.25) | 10.0 (2.42)<br>10 (8, 12) | 56.0 | 1.20 | 8.38 (1.79)<br>9 (7, 10) | 26.7 | 1.75 (1.08)<br>2 (1, 2) | 2.36 (1.48)<br>2 (1, 3) | 2.72 (1.52)<br>2 (1, 3) |
| BRA 2014[3] | 21,739 | 15.9 (0.81)<br>16 (15, 17) | 56.7 (11.8)<br>54.2 (48.4, 61.6) | 161 (6.15)<br>160 (156, 165) | NA | 4.60 (1.34)<br>5 (4, 5) | 54.7 | 14.4 (3.84)<br>14.25 (11.75, 17.25) | 6.14 (3.08)<br>6.25 (4, 8.25) | 8.30 (2.48)<br>8 (7, 10) | 46.8 | 1.27 | 7.71 (1.94)<br>8 (6, 9) | 17.6 | 2.20 (1.26)<br>2 (1, 3) | 3.50 (1.54)<br>3 (2, 5) | 3.96 (1.84)<br>4 (3, 5) |
| FJI 2004 | 1,230 | 30.0 (8.01)<br>30 (23, 37) | NA | NA | NA | 3.43 (1.29)<br>3 (3, 4) | 19.6 | 14.7 (3.63)<br>14.25 (12, 17) | 4.58 (3.08)<br>4.25 (2, 6.5) | 10.1 (1.76)<br>10 (9, 12) | 46.6 | 1.71 | 9.08 (1.68)<br>9 (8, 10) | 39.2 | 1.57 (1.22)<br>1 (1, 2) | 1.49 (1.11)<br>1 (1, 2) | 0.97 (1.03)<br>1 (0, 2) |
| MEX 2012[3] | 1,696 | 31.2 (10.2)<br>26 (18, 37) | 66.6 (14.8)<br>62.5 (53.5, 73.0) | 155 (7.20)<br>155 (150, 160) | 28.6 (5.58)<br>28.0 (24.8, 32.0) | 4.65 (1.31)<br>5 (4, 5) | 54.8 | 15.1 (3.47)<br>14.5 (12.25, 17.25) | 5.38 (2.83)<br>5 (3.25, 6.75) | 9.75 (2.10)<br>10 (8, 11) | 49.0 | 1.42 | 8.49 (1.96)<br>8 (7, 10) | 30.0 | 2.74 (1.38)<br>2 (2, 3) | 3.25 (1.52)<br>3 (2, 4) | 2.71 (1.74)<br>3 (1, 4) |
| **HIC** | 1,867 | 33.7 (9.79)<br>35 (26, 42) | 65.2 (15.1)<br>62.0 (55.0, 72.0)<br>*n*=1,852 | 164 (6.70)<br>165 (160, 168)<br>*n*=1,866 | 24.6 (5.56)<br>23.2 (20.8, 26.8)<br>*n*=1,628 | 5.10 (1.49)<br>5 (4, 6) | 67.7 | 16.7 (3.86)<br>16.5 (14.25, 19.25) | 6.80 (3.18)<br>6.5 (4.5, 8.75) | 9.91 (2.14)<br>10 (9, 11) | 68.4 | 5.41 | 8.87 (1.89)<br>9 (8, 10) | 37.7 | 2.68 (1.28)<br>3 (2, 4) | 2.80 (1.49)<br>3 (2, 4) | 2.67 (1.39)<br>3 (2, 3) |
| ITA 2005 | 855 | 34.0 (9.62)<br>35 (26, 42) | 60.4 (10.2)<br>59 (54, 65) | 164 (6.34)<br>165 (160, 169) | 22.5 (3.53)<br>21.9 (20.2, 23.9) | 5.30 (1.20)<br>5 (4, 6) | 75.0 | 18.1 (3.64)<br>18 (15.5, 20.5) | 7.82 (3.12)<br>7.5 (5.5, 10) | 10.3 (1.97)<br>10 (9, 12) | 81.2 | 9.24 | 8.76 (1.91)<br>9 (8, 10) | 34.5 | 2.85 (1.24)<br>3 (2, 4) | 3.10 (1.43)<br>3 (2, 4) | 2.82 (1.42)<br>3 (2, 4) |
| KNA 2020[3] | 401 | 35.7 (8.40)<br>36 (29, 43) | 81.1 (18.8)<br>78.0 (68.0, 90.7) | 165 (7.51)<br>165 (160, 168) | 30.1 (6.99)<br>29.1 (25.5, 33.6) | 3.78 (1.44)<br>4 (3, 5) | 29.9 | 14.4 (3.78)<br>14 (12, 17) | 4.72 (3.03)<br>4.25 (2, 6.75) | 9.67 (2.09)<br>10 (8, 11) | 44.4 | 0.75 | 8.62 (2.08)<br>9 (7, 10) | 34.7 | 1.87 (1.34)<br>2 (1, 3) | 2.25 (1.46)<br>2 (1, 3) | 2.84 (1.45)<br>3 (2, 4) |
| ROM 2012 (adults) | 473 | 36.4 (7.36)<br>37 (32, 42) | 64.1 (10.7)<br>62 (56, 70) | 165 (6.21)<br>165 (162, 168) | 23.6 (3.82)<br>23.0 (20.8, 25.7) | 5.80 (1.28)<br>6 (5, 7) | 86.1 | 16.5 (3.3)<br>16.5 (14.5, 18.5) | 6.92 (2.62)<br>6.75 (5, 8.75) | 9.59 (2.26)<br>10 (8, 11) | 67.9 | 3.38 | 9.30 (1.73)<br>9 (8, 10) | 46.1 | 3.07 (1.03)<br>3 (2, 4) | 2.77 (1.47)<br>3 (2, 4) | 2.29 (1.26)<br>2 (1, 3) |
| ROM 2012 (children) | 138 | 16.5 (1.12)<br>17 (16, 18) | 53.9 (8.56)<br>52 (49, 58) | 164 (8.38)<br>165 (160, 168) | NA | 5.29 (1.60)<br>5 (4, 7) | 70.3 | 15.6 (3.3)<br>15.75 (13.5, 17.5) | 6.09 (2.78)<br>6 (4, 8.25) | 9.52 (2.59)<br>10 (8, 11) | 60.9 | 2.17 | 8.89 (1.57)<br>9 (8, 10) | 37.7 | 2.56 (1.15)<br>3 (2, 3) | 2.67 (1.54)<br>2 (2, 4) | 2.54 (1.21)<br>3 (2, 3) |
| **Total** | 77,118 | 26. 8 (10.9)<br>24 (16, 36) | 56.0 (13.2)<br>54.1 (46.9, 62.8)<br>*n*=71,417 | 157 (7.49)<br>157 (152, 162)<br>*n*=71,433 | 23.5 (5.11)<br>22.7 (19.9, 26.3)<br>*n*=41,606 | 4.12 (1.41)<br>4 (3, 5) | 37.8 | 16.0 (3.95)<br>16 (13.25, 18.5) | 5.96 (3.14)<br>6 (4, 8) | 10.0 (2.41)<br>10 (9, 12) | 62.6 | 4.21 | 9.1 (2.22)<br>9 (8, 11) | 45.9 | 2.32 (1.36)<br>2 (1, 3) | 2.23 (1.66)<br>2 (1, 3) | 2.18 (2.00)<br>2 (0, 3) |

[1]Values are mean (SD) and median ($P^{25}$, $P^{75}$) or percentage. BGD, People's Republic of Bangladesh; BGR, Republic of Bulgaria; BMI, body mass index; BRA, Federative Republic of Brazil; FAO, Food and Agriculture Organization of the United Nations; FJI, Republic of Fiji; GDQS, Global Diet Quality Score; GDQS−, Global Quality Score Negative; GDQS+, Global Quality Score Positive; GDR, Global Dietary Recommendations; GIFT, Global Individual Food Consumption Data Tool; HIC, high income countries; IND, Republic of India; ITA, Republic of Italy; KNA, Saint Kitts and Nevis; LAO, Lao People's Democratic Republic; LIC, low income countries; LMIC, lower-middle income countries; MDD-W, Minimum Dietary Diversity for Women; MEX, United Mexican States; NA, not applicable; NCD, non-communicable disease; PHL, Republic of the Philippines; ROM, Romania; TUN, Tunisian Republic; UPF, ultra-processed food; UMIC, upper-middle income countries; WHO, World Health Organization.
[2]Among non-pregnant females aged 20-49 years.
[3]Values for mean (SD) and percentages are adjusted for survey weights, while medians are unadjusted.

#### Table S3. Descriptive statistics of primary quantitative reference metrics of dietary intake among non-pregnant females aged 15-49 years, by World Bank income classification and nationally representative surveys available open access on FAO/WHO GIFT[1]

| Characteristic | Sample size[2] | Macronutrient balance | | | Moderation | | | Nutrient adequacy | |
|---|---|---|---|---|---|---|---|---|---|
| **Survey** | | **40-70% E! from CHO** | **10-35% E! from protein[3]** | **15-30% E! from lipids[4]** | **<2,000 mg/day sodium[5]** | **<10% E! from free sugars** | **≤10% E! from SAFA** | **MAR of six micronutrients[6]** | **≥400 g/day of fruits and vegetables** |
| **LIC** | NA | 39.0<br>*n*=2,408 | 68.1<br>*n*=2,033 | 40.0<br>*n*=1,914 | 74.0<br>*n*=448 | NA | 72.8<br>*n*=456 | 0.65 (0.20)<br>0.67 (0.52, 0.80)<br>*n*=1,392 | 27.8 |
| **LMIC** | NA | 46.8<br>*n*=32,766 | 75.4<br>*n*=30,295 | 39.0<br>*n*=28,095 | 47.6<br>*n*=20,317 | NA | 84.5<br>*n*=7,850 | 0.62 (0.23)<br>0.60 (0.45, 0.81)<br>*n*=20,101 | 7.46 |
| BGD 2017 | 939 | 51.2 | 62.0<br>*n*=878 | 55.8<br>*n*=805 | 55.9<br>*n*=915 | NA | 99.3 | 0.51 (0.17)<br>0.48 (0.39, 0.61) | 2.56 |
| IND 2009 | 12,598 | 35.3 | 69.6<br>*n*=11,369 | 36.0<br>*n*=10,664 | 56.8<br>*n*=12,178 | NA | NA | 0.53 (0.17)<br>0.53 (0.41, 0.64) | 1.48 |
| LAO 2016 | 313 | 30.7 | 94.2<br>*n*=292 | 21.7<br>*n*=272 | 26.5<br>*n*=310 | NA | NA | NA | 16.9 |
| PHL 2003 | 1,205 | 29.9 | 83.7<br>*n*=1,181 | 26.8<br>*n*=1,124 | NA | NA | NA | NA | 3.49 |
| TUN 1996[7] | 2,118 | 91.2 | 96.6<br>*n*=1,840 | 65.5<br>*n*=1,688 | 2.50<br>*n*=2,022 | NA | 91.2 | 0.90 (0.10)<br>0.92 (0.83, 0.98) | 40.4 |
| **UMIC** | NA | 78.8<br>*n*=40,077 | 89.5<br>*n*=15,655 | 49.7<br>*n*=14,451 | 76.2<br>*n*=30,333 | 39.9<br>*n*=34,546 | 53.3<br>*n*=38,436 | 0.63 (0.20)<br>0.65 (0.51, 0.78)<br>*n*=25,145 | 8.75 |
| BGR 2004[7] | 343 | 67.6 | 84.4<br>*n*=262 | 12.7<br>*n*=237 | 42.9 | 84.0 | 24.8 | 0.71 (0.22)<br>0.75 (0.59, 0.88) | 11.7 |
| BRA 2008[7] | 11,594 | 85.4 | 94.4<br>*n*=10,479 | 53.5<br>*n*=9,758 | 77.6<br>*n*=11,188 | 67.2 | 52.4 | NA | 7.75 |
| BRA 2014[7] | 21,739 | 78.6 | NA | NA | 81.3<br>*n*=14,273 | 20.9 | 55.9 | 0.61 (0.19)<br>0.63 (0.49, 0.75) | 6.22 |
| FJI 2004 | 1,230 | 54.9 | 68.6<br>*n*=1,155 | 40.5<br>*n*=1,105 | NA | NA | NA | NA | 22.3 |
| MEX 2012[7] | 1,696 | 78.4 | 72.2<br>*n*=1,310 | 40.9<br>*n*=1,070 | 46.6<br>*n*=1,564 | NA | 43.6 | 0.75 (0.19)<br>0.76 (0.61, 0.89) | 17.4 |
| **HIC** | NA | 73.4<br>*n*=1,867 | 91.8<br>*n*=1,716 | 23.3<br>*n*=1,643 | 38.9<br>*n*=980 | NA | 46.9<br>*n*=1,866 | 0.79 (0.20)<br>0.85 (0.69, 0.96)<br>*n*=1,012 | 37.3 |
| ITA 2005 | 855 | 82.9 | 97.1<br>*n*=805 | 18.4<br>*n*=782 | NA | NA | 40.6 | NA | 45.4 |
| KNA 2020[7] | 401 | 69.0 | 89.1 | 42.1<br>*n*=390 | 60.1 | NA | 56.4 | 0.70 (0.22)<br>0.73 (0.56, 0.86) | 5.24 |
| ROM 2012 (adults) | 473 | 61.5 | 86.3 | 15.5<br>*n*=471 | 21.6 | NA | 51.2 | 0.88 (0.15)<br>0.93 (0.82, 0.99) | 52.2 |
| ROM 2012 (children) | 138 | 67.4 | 81.1<br>*n*=37 | NA | 34.0<br>*n*=106 | NA | 43.5 | 0.79 (0.20)<br>0.84 (0.70, 0.95) | 29.7 |
| **Total** | NA | 63.8<br>*n*=77,118 | 80.1<br>*n*=49,699 | 41.8<br>*n*=46,103 | 63.9<br>*n*=52,078 | 39.9<br>*n*=34,546 | 58.3<br>*n*=48,608 | 0.63 (0.21)<br>0.63 (0.48, 0.79)<br>*n*=47,650 | 9.49 |

[1]Values are mean (SD) and median ($P^{25}$, $P^{75}$) or percentage. BGD, People's Republic of Bangladesh; BGR, Republic of Bulgaria; BRA, Federative Republic of Brazil; CHO, carbohydrates; E!, energy (kcal/day); FAO, Food and Agriculture Organization of the United Nations; FJI, Republic of Fiji; GIFT, Global Individual Food Consumption Data Tool; HIC, high income countries; IND, Republic of India; ITA, Republic of Italy; KNA, Saint Kitts and Nevis; LAO, Lao People's Democratic Republic; LIC, low income countries; LMIC, lower-middle income countries; MAR, mean adequacy ratio; MEX, United Mexican States; NA, not applicable; PHL, Republic of the Philippines; ROM, Romania; SAFA, saturated fatty acids; TUN, Tunisian Republic; UMIC, upper-middle income countries; WHO, World Health Organization.
[2]For World Bank income classifications, sample sizes are reported separately under each primary quantitative reference metrics of dietary intake, due to heterogeneous data availability.
[3]Among individuals aged 18 years and older.
[4]Among individuals aged 20 years and older.
[5]Among individuals aged 16 years and older.
[6]Vitamin A, vitamin C, folate, calcium, iron, and zinc. The selection of micronutrients was informed by the prevalence of nutrient deficiencies globally and the availability of dietary intake data from surveys on FAO/WHO GIFT (i.e., leading to the exclusion of vitamin D, iodine, and vitamin K in the composite measure).
[7]Values for mean (SD) and percentages are adjusted for survey weights, while medians are unadjusted.

#### Table S4. Descriptive statistics of secondary quantitative reference metrics of dietary intake among non-pregnant females aged 15-49 years, by World Bank income classification and nationally representative surveys available open access on FAO/WHO GIFT[1]

| Characteristic | Sample size[2] | Macronutrient balance | | | Moderation | | | | Nutrient adequacy | | |
|---|---|---|---|---|---|---|---|---|---|---|---|
| **Survey** | | **5-10% E! from *n*-6 PUFA** | **0.6-1.2% E! from *n*-3 PUFA** | **≥10% E! from PUFA** | **≤1% E! from TFA** | **<5% E! from free sugars** | **≥25 g/day of dietary fibre** | **% E! from UPF** | **MAR of 11 micronutrients[3]** | **≥3,510 mg/day of potassium[4]** | **≥250 mg/day of n-3 PUFA** |
| **LIC** | NA | NA | NA | 2.85<br>*n*=456 | NA | NA | 47.5<br>*n*=945 | 16.1 (24.6)<br>0.69 (0.00, 28.3)<br>*n*=2,408 | 0.68 (0.18)<br>0.71 (0.56, 0.82)<br>*n*=1,228 | 14.3<br>*n*=448 | NA |
| **LMIC** | NA | NA | NA | 26.7<br>*n*=8,380 | 96.7<br>*n*=306 | NA | 41.0<br>*n*=30,732 | 13.6 (19.7)<br>1.08 (0.00, 23.9)<br>*n*=32,766 | 0.79 (0.19)<br>0.85 (0.68, 0.94)<br>*n*=7,137 | 15.1<br>*n*=8,195 | NA |
| BGD 2017 | 939 | NA | NA | 35.9 | NA | NA | 23.9 | 0.52 (1.96)<br>0.00 (0.00, 0.00) | 0.61 (0.13)<br>0.60 (0.51, 0.70)<br>*n*=939 | 0.66<br>*n*=915 | NA |
| IND 2009 | 12,598 | NA | NA | NA | NA | NA | 59.3 | 1.00 (2.97)<br>0.00 (0.00, 0.00) | NA | NA | NA |
| LAO 2016 | 313 | NA | NA | NA | NA | NA | 1.28 | 6.04 (9.84)<br>1.62 (0.00, 7.79) | NA | NA | NA |
| PHL 2003 | 1,205 | NA | NA | NA | NA | NA | NA | 2.11 (4.00)<br>0.00 (0.00, 3.15) | NA | NA | NA |
| TUN 1996[5] | 2,118 | NA | NA | 49.1 | NA | NA | 77.4 | 33.1 (17.0)<br>33.1 (21.1, 43.9) | 0.89 (0.10)<br>0.90 (0.83, 0.96) | 16.6<br>*n*=2,022 | NA |
| **UMIC** | NA | 36.8<br>*n*=13,870 | 53.8<br>*n*=11,594 | 5.71<br>*n*=38,436 | 62.5<br>*n*=37,306 | 25.3<br>*n*=34,546 | 19.5<br>*n*=39,956 | 46.8 (23.0)<br>46.5 (29.8, 63.8)<br>*n*=40,077 | 0.65 (0.20)<br>0.67 (0.52, 0.81)<br>*n*=25,145 | 7.70<br>*n*=30,966 | 98.4<br>*n*=11,594 |
| BGR 2004[5] | 343 | NA | NA | 65.0 | NA | 60.9 | 18.7 | 41.6 (21.0)<br>41.8 (25.6, 57.5) | 0.74 (0.20)<br>0.77 (0.61, 0.90) | 9.62 | NA |
| BRA 2008[5] | 11,594 | 42.5 | 53.8 | 4.94 | 43.0 | 46.9 | 19.2 | 39.6 (20.7)<br>36.1 (22.2, 51.7) | NA | 9.26<br>*n*=11,188 | 98.4 |
| BRA 2014[5] | 21,739 | NA | NA | 3.72 | 67.3 | 9.80 | 22.1 | 51.9 (22.5)<br>52.8 (35.5, 69.3) | 0.63 (0.20)<br>0.65 (0.50, 0.79) | 5.39<br>*n*=14,273 | NA |
| FJI 2004 | 1,230 | NA | NA | NA | NA | NA | 25.2 | 27.7 (22.2)<br>26.5 (6.80, 43.1) | NA | 28.8<br>*n*=1,213 | NA |
| MEX 2012[5] | 1,696 | 11.6 | NA | 15.0 | 94.3 | NA | 10.3 | 52.1 (18.5)<br>54.0 (41.4, 66.6) | 0.74 (0.19)<br>0.76 (0.60, 0.90) | 5.48<br>*n*=1,564 | NA |
| **HIC** | NA | 3.76<br>*n*=611 | NA | 36.6<br>*n*=1011 | 99.4<br>*n*=611 | NA | 17.9<br>*n*=1,867 | 35.0 (20.9)<br>31.4 (19.7, 47.1)<br>*n*=1,867 | 0.81 (0.19)<br>0.88 (0.72, 0.96)<br>*n*=1,012 | 13.9<br>*n*=1,819 | NA |
| ITA 2005 | 855 | NA | NA | NA | NA | NA | 13.0 | 28.6 (14.4)<br>27.9 (17.9, 37.6) | NA | 21.3<br>*n*=839 | NA |
| KNA 2020[5] | 401 | NA | NA | 17.1<br>*n*=400 | NA | NA | 6.95 | 52.7 (24.2)<br>55.0 (34.2, 70.4) | 0.74 (0.20)<br>0.78 (0.61, 0.90) | 2.81 | NA |
| ROM 2012 (adults) | 473 | 2.75 | NA | 51.0 | 99.2 | NA | 33.8 | 31.0 (19.0)<br>27.2 (16.0, 42.5) | 0.87 (0.15)<br>0.93 (0.83, 0.98) | 11.4 | NA |
| ROM 2012 (children) | 138 | 7.25 | NA | 44.2 | 100 | NA | 26.1 | 36.0 (21.7)<br>33.2 (20.1, 51.4) | 0.81 (0.19)<br>0.88 (0.70, 0.95) | 7.55<br>*n*=106 | NA |
| **Total** | NA | 35.4<br>*n*=14,481 | 53.8<br>*n*=11,594 | 9.98<br>*n*=48,283 | 63.3<br>*n*=38,223 | NA | 28.8<br>*n*=73,500 | 31.5 (27.1)<br>29.3 (1.65, 52.1)<br>*n*=77,118 | 0.69 (0.21)<br>0.71 (0.55, 0.85)<br>*n*=34,522 | 9.51<br>*n*=41,428 | 98.4<br>*n*=11,594 |

[1]Values are mean (SD) and median ($P^{25}$, $P^{75}$) or percentage. BGD, People's Republic of Bangladesh; BGR, Republic of Bulgaria; BRA, Federative Republic of Brazil; E!, energy (kcal/day); FAO, Food and Agriculture Organization of the United Nations; FJI, Republic of Fiji; GIFT, Global Individual Food Consumption Data Tool; HIC, high income countries; IND, Republic of India; ITA, Republic of Italy; KNA, Saint Kitts and Nevis; LAO, Lao People's Democratic Republic; LIC, low income countries; LMIC, lower-middle income countries; MEX, United Mexican States; NA, not applicable; PHL, Republic of the Philippines; PUFA, poly-unsaturated fatty acids; ROM, Romania; TFA, trans-fatty acids; TUN, Tunisian Republic; UPF, ultra-processed food; UMIC, upper-middle income countries; WHO, World Health Organization.
[2]For World Bank income classifications, sample sizes are reported separately under each primary quantitative reference metrics of dietary intake, due to heterogeneous data availability.
[3]Vitamin A, thiamine, riboflavin, niacin, vitamin B6, vitamin B12, vitamin C, folate, calcium, iron, and zinc.
[4]Among individuals aged 16 years and older.
[5]Values for mean (SD) and percentages are adjusted for survey weights, while medians are unadjusted.

### Table S5. Associations between mean-standardized healthy diet measures and primary quantitative reference metrics of dietary intake among non-pregnant females aged 15-49 years, by World Bank income classification using surveys available open access on FAO/WHO GIFT[1]

| Characteristic | Macronutrient balance | | | Moderation | | | Nutrient adequacy | |
|---|---|---|---|---|---|---|---|---|
| **Outcome** | **40-70% E! from CHO** | **10-35% E! from protein[2]** | **15-30% E! from lipids[3]** | **<2,000 mg/day sodium[4]** | **<10% E! from free sugars** | **≤10% E! from SAFA** | **MAR of six micronutrients[5,6]** | **≥400 g/day of fruits and vegetables** |
| **LIC, *n*** | 2,408 | 2,033 | 1,914 | 448 | NA | 456 | 1,392 | 2,408 |
| FGDS | 1.78 (1.58, 2.02) | 1.62 (1.39, 1.89) | 1.67 (1.46, 1.91) | 0.24 (0.16, 0.35) | NA | 1.07 (0.82, 1.38) | 0.11 (0.09, 0.13), 0.15 | 2.27 (1.97, 2.63) |
| GDQS+ | 1.75 (1.58, 1.93) | 1.45 (1.28, 1.64) | 1.71 (1.53, 1.90) | 0.40 (0.30, 0.54) | NA | 1.16 (0.91, 1.49) | 0.10 (0.06, 0.13), 0.15 | 1.50 (1.33, 1.69) |
| GDQS− | 0.97 (0.88, 1.08) | 1.35 (1.19, 1.53) | 0.89 (0.80, 0.99) | 1.53 (1.19, 1.96) | NA | 0.84 (0.69, 1.03) | -0.02 (-0.04, -0.01), 0.18 | 1.09 (0.97, 1.21) |
| NCD-Protect score | 1.19 (1.07, 1.32) | 1.11 (0.97, 1.28) | 1.41 (1.25, 1.59) | 0.50 (0.37, 0.67) | NA | 1.23 (0.98, 1.55) | 0.10 (0.08, 0.12), 0.16 | 2.61 (2.27, 3.01) |
| 9−NCD-Risk score[7] | 0.78 (0.71, 0.85) | 0.98 (0.87, 1.10) | 0.82 (0.75, 0.91) | 1.63 (1.26, 2.10) | NA | 0.88 (0.73, 1.06) | -0.01 (-0.02, 0.00), 0.18 | 1.01 (0.90, 1.12) |
| 23−Nova UPF score[7] | 0.87 (0.79, 0.95) | 1.01 (0.90, 1.13) | 0.78 (0.71, 0.86) | 1.37 (1.16, 1.61) | NA | 0.96 (0.82, 1.11) | -0.02 (-0.04, -0.01), 0.18 | 1.06 (0.96, 1.17) |
| **LMIC, *n*** | 32,766 | 30,295 | 28,095 | 20,317 | NA | 7,850 | 20,101 | 32,766 |
| FGDS | 1.60 (1.55, 1.64) | 1.76 (1.69, 1.82) | 1.36 (1.32, 1.40) | 0.53 (0.50, 0.55) | NA | 0.72 (0.67, 0.78) | 0.11 (0.08, 0.14), 0.15 | 2.29 (2.16, 2.42) |
| GDQS+ | 1.61 (1.57, 1.66) | 2.48 (2.38, 2.58) | 1.34 (1.30, 1.38) | 0.56 (0.54, 0.58) | NA | 0.91 (0.85, 0.98) | 0.10 (0.07, 0.12), 0.14 | 1.82 (1.73, 1.92) |
| GDQS− | 1.04 (1.01, 1.07) | 1.38 (1.34, 1.42) | 1.36 (1.32, 1.40) | 1.33 (1.28, 1.38) | NA | 0.94 (0.87, 1.01) | -0.02 (-0.04, 0.00), 0.16 | 1.02 (0.96, 1.08) |
| NCD-Protect score | 1.17 (1.14, 1.20) | 1.32 (1.27, 1.36) | 1.34 (1.30, 1.38) | 0.62 (0.59, 0.64) | NA | 1.03 (0.96, 1.13) | 0.10 (0.06, 0.14), 0.15 | 2.56 (2.40, 2.72) |
| 9−NCD-Risk score[7] | 0.57 (0.56, 0.59) | 0.92 (0.89, 0.95) | 0.71 (0.69, 0.73) | 1.01 (0.95, 1.08) | NA | 1.72 (1.58, 1.88) | -0.05 (-0.08, -0.01), 0.16 | 0.98 (0.92, 1.04) |
| 23−Nova UPF score[7] | 0.68 (0.66, 0.71) | 0.98 (0.93, 1.02) | 0.77 (0.74, 0.80) | 1.19 (1.08, 1.31) | NA | 1.61 (1.46, 1.77) | -0.01 (-0.05, 0.02), 0.16 | 1.13 (1.05, 1.22) |
| **UMIC, *n*** | 40,077 | 15,655 | 14,451 | 30,333 | 34,546 | 38,436 | 25,145 | 40,077 |
| FGDS | 1.27 (1.24, 1.30) | 2.12 (2.00, 2.26) | 1.08 (1.04, 1.11) | 0.88 (0.86, 0.91) | 0.85 (0.84, 0.87)[8] | 0.93 (0.91, 0.95) | 0.10 (0.08, 0.12), 0.17 | 2.91 (2.79, 3.03) |
| GDQS+ | 1.18 (1.15, 1.21) | 1.71 (1.61, 1.82) | 1.22 (1.18, 1.26) | 0.97 (0.95, 1.00) | 1.00 (0.98, 1.02)[8] | 1.30 (1.28, 1.33) | 0.08 (0.08, 0.09), 0.17 | 2.63 (2.53, 2.74) |
| GDQS− | 0.83 (0.81, 0.85) | 1.19 (1.12, 1.25) | 1.02 (0.99, 1.05) | 1.59 (1.54, 1.63) | 3.06 (2.97, 3.14)[8] | 1.01 (0.99, 1.03) | -0.05 (-0.05, -0.05), 0.18[9] | 1.07 (1.03, 1.11) |
| NCD-Protect score | 1.08 (1.05, 1.11) | 1.30 (1.22, 1.37) | 1.15 (1.11, 1.19) | 1.00 (0.97, 1.03) | 0.88 (0.86, 0.90)[8] | 1.09 (1.07, 1.11) | 0.07 (0.06, 0.07), 0.18 | 3.01 (2.90, 3.13) |
| 9−NCD-Risk score[7] | 0.77 (0.75, 0.79) | 0.81 (0.76, 0.86) | 1.22 (1.18, 1.27) | 2.09 (2.02, 2.15) | 2.91 (2.82, 2.99)[8] | 1.39 (1.36, 1.43) | -0.04 (-0.04, -0.03), 0.19 | 1.07 (1.03, 1.11) |
| 23−Nova UPF score[7] | 0.67 (0.65, 0.69) | 0.87 (0.82, 0.93) | 1.16 (1.12, 1.20) | 1.95 (1.89, 2.02) | 3.52 (3.40, 3.64)[8] | 1.36 (1.33, 1.39) | -0.05 (-0.06, -0.04), 0.18 | 1.05 (1.01, 1.09) |
| **HIC, *n*** | 1,867 | 1,716 | 1,643 | 980 | NA | 1,866 | 1,012 | 1,867 |
| FGDS | 0.97 (0.87, 1.09) | 2.40 (1.94, 2.97) | 0.72 (0.63, 0.83) | 0.55 (0.47, 0.64) | NA | 0.78 (0.70, 0.86) | 0.11 (0.09, 0.12), 0.16 | 2.42 (2.10, 2.78) |
| GDQS+ | 0.95 (0.84, 1.06) | 1.88 (1.50, 2.36) | 0.83 (0.73, 0.95) | 0.69 (0.58, 0.81) | NA | 1.07 (0.97, 1.18) | 0.07 (0.05, 0.10), 0.18 | 2.50 (2.19, 2.86) |
| GDQS− | 0.76 (0.68, 0.85) | 1.20 (1.01, 1.42) | 0.95 (0.84, 1.07) | 1.62 (1.40, 1.88) | NA | 0.96 (0.87, 1.05) | -0.05 (-0.06, -0.04), 0.18 | 0.96 (0.87, 1.07) |
| NCD-Protect score | 1.15 (1.03, 1.29) | 1.02 (0.84, 1.24) | 1.05 (0.93, 1.19) | 0.84 (0.72, 0.97) | NA | 1.04 (0.94, 1.14) | 0.07 (0.06, 0.10), 0.17 | 2.85 (2.49, 3.27) |
| 9−NCD-Risk score[7] | 0.91 (0.81, 1.01) | 0.69 (0.57, 0.83) | 1.25 (1.10, 1.41) | 1.89 (1.62, 2.21) | NA | 1.44 (1.31, 1.59) | -0.06 (-0.09, -0.03), 0.18 | 1.01 (0.91, 1.12) |
| 23−Nova UPF score[7] | 0.75 (0.67, 0.84) | 0.99 (0.82, 1.18) | 1.12 (0.99, 1.26) | 2.22 (1.87, 2.64) | NA | 1.26 (1.15, 1.39) | -0.07 (-0.09, -0.05), 0.17 | 1.06 (0.95, 1.17) |

[1]Healthy diet measures were transformed to age range specific *z*-scores using the mean and SD within each World Bank income classification. Unless otherwise stated, values are the odds ratios (95% CIs) of a one-SD increment in a healthy diet measure from mixed effects logistic regression models with a random intercept for each survey. CHO, carbohydrates; E!, energy (kcal/day); FAO, Food and Agriculture Organization of the United Nations; GDQS−, Global Quality Score Negative; GDQS+, Global Quality Score Positive; GIFT, Global Individual Food consumption data Tool; HIC, high income countries; LIC, low income countries; LMIC, lower-middle income countries; MAR, mean adequacy ratio; NA, not applicable; NCD, non-communicable disease; SAFA, saturated fatty acids; UPF, ultra-processed food; UMIC, upper-middle income countries; WHO, World Health Organization.
[2]Among non-pregnant females aged 18-49 years.
[3]Among non-pregnant females aged 20-49 years.
[4]Among non-pregnant females aged 16-49 years.
[5]Vitamin A, vitamin C, folate, calcium, iron, and zinc. The selection of micronutrients was informed by the prevalence of nutrient deficiencies globally and the availability of dietary intake data from surveys on FAO/WHO GIFT (i.e., leading to the exclusion of vitamin D, iodine, and vitamin K in the composite measure).
[6]Values are coefficients (95% CIs) and SD of the residuals from mixed effects linear regression models with a random intercept for each survey and a random slope for the healthy diet measure.
[7]Likewise to GDQS−, higher scores of the complement of NCD-Risk and Nova UPF scores reflect lower consumption of food groups associated with increased risk for diet-related NCDs.
[8]Logistic regression due to failure of model convergence with a random intercept for each survey.
[9]Mixed effects linear regression model with a random intercept for each survey only due to failure of model convergence with a random slope for measures of healthy diets.

#### Table S6. Associations between mean-standardized healthy diet measures and secondary quantitative reference metrics of dietary intake among non-pregnant females aged 15-49 years, by World Bank income classification using surveys available open access on FAO/WHO GIFT[1]

| **Characteristic** | **Macronutrient balance** | | | **Moderation** | | | | **Nutrient adequacy** | | |
|---|---|---|---|---|---|---|---|---|---|---|
| **Outcome** | **5-10% E! from *n*-6 PUFA** | **0.6-1.2% E! from *n*-3 PUFA** | **≥10% E! from PUFA** | **≤1% E! from TFA** | **<5% E! from free sugars** | **≥25 g/day of dietary fibre** | **pp E! from UPF[2]** | **MAR of 11 micronutrients[2,3]** | **≥3,510 mg/day of potassium[4]** | **≥250 mg/day of *n*-3 PUFA** |
| **LIC, *n*** | NA | NA | 456 | NA | NA | 945 | 2,408 | 1,228 | 448 | NA |
| FGDS | NA | NA | 0.90 (0.45, 1.79) | NA | NA | 2.78 (2.22, 3.48) | -2.23 (-4.46, 0.00), 13.1 | 0.11 (0.10, 0.12), 0.14 | 2.74 (1.91, 3.94) | NA |
| GDQS+ | NA | NA | 1.44 (0.79, 2.62) | NA | NA | 3.07 (2.48, 3.81) | -1.43 (-3.12, 0.27), 13.1 | 0.09 (0.08, 0.10), 0.15 | 2.56 (1.82, 3.60) | NA |
| GDQS− | NA | NA | 0.79 (0.48, 1.30) | NA | NA | 0.71 (0.61, 0.84) | -1.59 (-3.17, 0.00), 13.1 | -0.03 (-0.03, -0.02), 0.17 | 0.83 (0.65, 1.06) | NA |
| NCD-Protect score | NA | NA | 0.85 (0.46, 1.56) | NA | NA | 2.84 (2.30, 3.50) | -2.06 (-3.29, -0.83), 13.1 | 0.08 (0.07, 0.09), 0.15 | 2.35 (1.73, 3.19) | NA |
| 9−NCD-Risk score[5] | NA | NA | 0.84 (0.54, 1.32) | NA | NA | 1.00 (0.87, 1.16) | 0.52 (-1.87, 2.92), 13.0 | -0.02 (-0.03, -0.02), 0.17 | 0.90 (0.71, 1.14) | NA |
| 23−Nova UPF score[5] | NA | NA | 0.89 (0.63, 1.25) | NA | NA | 0.98 (0.87, 1.11) | -3.41 (-5.51, -1.31), 12.8 | -0.03 (-0.03, -0.02), 0.17 | 1.01 (0.82, 1.24) | NA |
| **LMIC, *n*** | NA | NA | 8,380 | 306 | NA | 30,732 | 32,766 | 7,137 | 8,195 | NA |
| FGDS | NA | NA | 1.13 (1.06, 1.20) | 1.79 (1.11, 2.87) | NA | 1.73 (1.69, 1.77)[6] | -0.92 (-2.41, 0.57), 13.2 | 0.10 (0.07, 0.13), 0.11 | 2.15 (1.99, 2.33) | NA |
| GDQS+ | NA | NA | 1.25 (1.18, 1.32) | 1.42 (1.12, 1.80) | NA | 3.12 (3.02, 3.22)[6] | -1.78 (-3.51, 0.00), 13.1 | 0.08 (0.05, 0.11), 0.11 | 1.97 (1.84, 2.12) | NA |
| GDQS− | NA | NA | 1.10 (1.03, 1.17) | 0.99 (0.73, 1.36) | NA | 1.32 (1.29, 1.35)[6] | -4.29 (-6.74, -1.84), 12.0 | -0.03 (-0.05, -0.01), 0.13 | 0.74 (0.68, 0.79) | NA |
| NCD-Protect score | NA | NA | 1.14 (1.06, 1.21) | 1.96 (1.10, 3.49) | NA | 2.76 (2.69, 2.85)[6] | -2.02 (-3.27, -0.77), 13.2 | 0.08 (0.04, 0.13), 0.12 | 2.11 (1.94, 2.30) | NA |
| 9−NCD-Risk score[5] | NA | NA | 1.22 (1.14, 1.32) | 1.02 (0.66 1.57) | NA | 1.67 (1.63, 1.72)[6] | -4.00 (-5.60, -2.40), 12.5 | -0.05 (-0.09, -0.01), 0.13 | 0.85 (0.78, 0.93) | NA |
| 23−Nova UPF score[5] | NA | NA | 1.41 (1.29, 1.54) | 0.88 (0.57, 1.35) | NA | 2.16 (2.09, 2.22)[6] | -13.5 (-17.7, -9.35), 10.9 | -0.01 (-0.05, 0.03), 0.13 | 0.89 (0.80, 0.99) | NA |
| **UMIC, *n*** | 13,870 | 11,594 | 38,436 | 37,306 | 34,546 | 39,56 | 40,077 | 25,145 | 30,966 | 11,594 |
| FGDS | 1.03 (0.99, 1.07) | 1.34 (1.28, 1.39) | 0.77 (0.73, 0.81) | 1.10 (1.08, 1.11) | 0.77 (0.75, 0.79)[6] | 2.00 (1.93, 2.06) | -4.00 (-5.33, 2.67), 20.8 | 0.07 (0.07, 0.07), 0.18[7] | 1.96 (1.87, 2.05) | 7.24 (5.92, 8.85) |
| GDQS+ | 1.45 (1.39, 1.51) | 1.42 (1.36, 1.49) | 0.96 (0.92, 1.01) | 1.08 (1.07, 1.09) | 0.92 (0.90, 0.94)[6] | 2.68 (2.59, 2.76) | -4.73 (-6.43, -3.03), 20.5 | 0.06 (0.06, 0.06), 0.19[7] | 2.14 (2.04, 2.24) | 6.82 (5.52, 8.41) |
| GDQS− | 1.06 (1.02, 1.10) | 1.29 (1.24, 1.35) | 1.01 (0.97, 1.06) | 1.05 (1.04, 1.06) | 4.03 (3.89, 4.19)[6] | 0.79 (0.76, 0.80) | -6.06 (-7.61, -4.50), 20.1 | -0.05 (-0.05, -0.05), 0.19[7] | 0.58 (0.56, 0.61) | 0.60 (0.51, 0.70) |
| NCD-Protect score | 0.98 (0.94, 1.02) | 1.23 (1.18, 1.29) | 0.82 (0.78, 0.86) | 1.09 (1.07, 1.11) | 0.80 (0.78, 0.83)[6] | 1.77 (1.73, 1.82) | -3.36 (-4.33, 2.39), 21.2 | 0.04 (0.04, 0.04), 0.19[7] | 1.70 (1.63, 1.77) | 4.73 (3.77, 5.92) |
| 9−NCD-Risk score[5] | 1.25 (1.21, 1.31) | 1.17 (1.13, 1.22) | 1.22 (1.16, 1.28) | 1.10 (1.09, 1.12) | 3.96 (3.82, 4.11)[6] | 0.88 (0.85, 0.90) | -6.44 (-8.41, -4.48), 20.0 | -0.06 (-0.06, -0.05), 0.19[7] | 0.74 (0.71, 0.78) | 0.45 (0.37, 0.53) |
| 23−Nova UPF score[5] | 0.98 (0.93, 1.02) | 1.11 (1.06, 1.16) | 1.23 (1.17, 1.30) | 1.14 (1.13, 1.14) | 5.37 (5.15, 5.61)[6] | 0.88 (0.86, 0.90) | -10.2 (-13.3, -7.18), 18.8 | -0.08 (-0.08, -0.08), 0.18[7] | 0.69 (0.66, 0.72) | 0.31 (0.25, 0.39) |
| **HIC, *n*** | 611 | NA | 1,011 | 611 | NA | 1,867 | 1,867 | 1,012 | 1,819 | NA |
| FGDS | 0.69 (0.46, 1.05) | NA | 1.17 (1.01, 1.36) | 0.97 (0.47, 2.00) | NA | 2.69 (2.27, 3.19) | -5.27 (-8.35, -2.19), 18.0 | 0.10 (0.09, 0.10), 0.16[7] | 2.53 (2.10, 3.05) | NA |
| GDQS+ | 0.74 (0.43, 1.25) | NA | 1.62 (1.38, 1.91) | 0.86 (0.62, 1.21) | NA | 2.44 (2.10, 2.83) | -6.05 (-9.84, -2.27), 17.8 | 0.06 (0.05, 0.08), 0.17[7] | 2.26 (1.94, 2.64) | NA |
| GDQS− | 0.55 (0.38, 0.79) | NA | 0.92 (0.81, 1.05) | 0.83 (0.53, 1.31) | NA | 0.73 (0.65, 0.83) | -5.04 (-5.87, -4.21), 0.18 | -0.05 (-0.06, -0.04), 0.17[7] | 0.56 (0.49, 0.65) | NA |
| NCD-Protect score | 0.72 (0.43, 1.19) | NA | 1.12 (0.97, 1.31) | 0.77 (0.30, 1.96) | NA | 2.47 (2.12, 2.87) | -2.72 (-5.05, -0.39), 18.4 | 0.06 (0.04, 0.07), 0.17[7] | 2.07 (1.77, 2.41) | NA |
| 9−NCD-Risk score[5] | 0.86 (0.57, 1.30) | NA | 1.01 (0.88, 1.16) | 1.00 (0.52, 1.95) | NA | 0.73 (0.65, 0.83) | -4.71 (-6.18, -3.23), 18.1 | -0.06 (-0.07, -0.05), 0.17[7] | 0.73 (0.64, 0.85) | NA |
| 23−Nova UPF score[5] | 0.84 (0.54, 1.32) | NA | 0.93 (0.80, 1.08) | 0.50 (0.18, 1.36) | NA | 0.73 (0.64, 0.82) | -9.16 (-10.5, -7.84), 16.5 | -0.07 (-0.08, -0.07), 0.17[7] | 0.73 (0.64, 0.84) | NA |

[1]Healthy diet measures were transformed to age range specific *z*-scores using the mean and SD within each World Bank income classification. Unless otherwise stated, values are the odds ratios (95% CIs) of a one-SD increment in a healthy diet measure from mixed effects logistic regression models with a random intercept for each survey. E!, energy (kcal/day); FAO, Food and Agriculture Organization of the United Nations; GDQS−, Global Quality Score Negative; GDQS+, Global Quality Score Positive; GIFT, Global Individual Food consumption data Tool; HIC, high income countries; LIC, low income countries; LMIC, lower-middle income countries; NA, not applicable; NCD, non-communicable disease; pp, percentage point; PUFA, poly-unsaturated fatty acids; TFA, trans-fatty acids; UPF, ultra-processed food; UMIC, upper-middle income countries; WHO, World Health Organization.
[2]Values are coefficients (95% CIs) and SD of the residuals from mixed effects linear regression models with a random intercept for each survey and a random slope for the healthy diet measure.
[3]Vitamin A, thiamine, riboflavin, niacin, vitamin B6, vitamin B12, vitamin C, folate, calcium, iron, and zinc.
[4]Among individuals aged 16 years and older.
[5]Likewise to GDQS−, higher scores of the complement of NCD-Risk and Nova UPF scores reflect lower consumption of food groups associated with increased risk for diet-related NCDs.
[6]Logistic regression due to failure of model convergence with a random intercept for each survey.
[7]Mixed effects linear regression model with a random intercept for each survey only due to failure of model convergence with a random slope for measures of healthy diets.

#### Table S7. Accuracy of healthy diet indicators for 10-35% E! from protein among non-pregnant females aged 18-49 years, by World Bank income classification using surveys available open access on FAO/WHO GIFT[1]

| **Characteristic** | **Macronutrient balance** | | | | |
|---|---|---|---|---|---|
| **Outcome** | **10-35% E! from protein** | | | | |
| **LIC, *n*=2,033** | **Sensitivity** | **Specificity** | **PCC** | **LR+** | **LR−** |
| FGDS ≥3 | 80.4 | 14.6 | 59.4 | 0.942 | 1.34 |
| FGDS ≥4 | 49.4 | 45.5 | 48.2 | 0.906 | 1.11 |
| FGDS ≥5 | 27.6 | 69.7 | 41.0 | 0.909 | 1.04 |
| GDQS+ ≥6 | 64.0 | 30.5 | 53.3 | 0.921 | 1.18 |
| GDQS+ ≥7 | 48.3 | 49.6 | 48.7 | 0.958 | 1.04 |
| GDQS+ ≥8 | 44.4 | 56.5 | 48.3 | 1.02 | 0.984 |
| GDQS+ ≥9 | 29.1 | 75.0 | 43.7 | 1.16 | 0.945 |
| **LMIC, *n*=30,295** | **Sensitivity** | **Specificity** | **PCC** | **LR+** | **LR−** |
| FGDS ≥3 | 85.6 | 25.2 | 70.8 | 1.14 | 0.572 |
| FGDS ≥4 | 59.0 | 60.1 | 59.3 | 1.48 | 0.681 |
| FGDS ≥5 | 31.7 | 85.6 | 44.92 | 2.19 | 0.798 |
| GDQS+ ≥6 | 59.2 | 64.7 | 60.6 | 1.68 | 0.631 |
| GDQS+ ≥7 | 43.4 | 81.0 | 52.6 | 2.29 | 0.699 |
| GDQS+ ≥8 | 34.9 | 86.4 | 47.5 | 2.56 | 0.754 |
| GDQS+ ≥9 | 21.3 | 93.8 | 39.1 | 3.46 | 0.839 |
| **UMIC, *n*=15,655** | **Sensitivity** | **Specificity** | **PCC** | **LR+** | **LR−** |
| FGDS ≥3 | 92.3 | 27.6 | 85.5 | 1.27 | 0.28 |
| FGDS ≥4 | 68.6 | 56.1 | 67.3 | 1.56 | 0.56 |
| FGDS ≥5 | 38.0 | 79.5 | 42.4 | 1.85 | 0.78 |
| GDQS+ ≥6 | 48.7 | 70.1 | 50.9 | 1.63 | 0.732 |
| GDQS+ ≥7 | 25.7 | 85.0 | 31.9 | 1.71 | 0.875 |
| **HIC, *n*=1,716** | **Sensitivity** | **Specificity** | **PCC** | **LR+** | **LR−** |
| FGDS ≥3 | 95.8 | 17.9 | 89.4 | 1.17 | 0.238 |
| FGDS ≥4 | 86.4 | 37.1 | 82.4 | 1.37 | 0.366 |
| FGDS ≥5 | 69.1 | 57.1 | 68.1 | 1.61 | 0.541 |
| FGDS ≥6 | 43.2 | 78.6 | 46.0 | 2.01 | 0.724 |
| GDQS+ ≥5 | 71.6 | 50.7 | 69.9 | 1.45 | 0.559 |
| GDQS+ ≥6 | 64.0 | 59.3 | 63.6 | 1.57 | 0.61 |
| GDQS+ ≥7 | 47.7 | 75.7 | 49.9 | 1.96 | 0.691 |

[1]Values are percentages, except for AUCs. AUC, area under the curve; E!, energy (kcal/day); FAO, Food and Agriculture Organization of the United Nations; FGDS, food group diversity score; GDQS+, Global Quality Score Positive; GIFT, Global Individual Food consumption data Tool; HIC, high income countries; LIC, low income countries; LMIC, lower-middle income countries; LR+; positive likelihood ratio; LR-, negative likelihood ratio; PCC, percentage correctly classified; UMIC, upper-middle income countries; WHO, World Health Organization.

#### Table S8. Spearman's rank correlations (*ρ*) between mean healthy diet measures and mean of primary quantitative reference metrics of dietary intake using surveys available open access on FAO/WHO GIFT[1]

| **Characteristic** | **Macronutrient balance** | | | **Moderation** | | | **Nutrient adequacy** | |
|---|---|---|---|---|---|---|---|---|
| **Outcome** | **40-70% E! from CHO** | **10-35% E! from protein[2]** | **15-30% E! from lipids[3]** | **<2,000 mg/day sodium[4]** | **<10% E! from free sugars** | **≤10% E! from SAFA** | **MAR of six micronutrients[5]** | **≥400 g/day of fruits and vegetables** |
| **Total, *n*** | 44 | 43 | 40 | 26 | 5 | 22 | 23 | 44 |
| FGDS | 0.47 | 0.42 | 0.11 | -0.59 | -0.60 | -0.28 | 0.79 | 0.66 |
| GDQS+ | 0.30 | 0.02 | 0.42 | -0.49 | -0.80 | 0.26 | 0.56 | 0.64 |
| GDQS− | -0.46 | -0.54 | 0.15 | 0.29 | 0.30 | 0.38 | -0.49 | -0.15 |
| NCD-Protect score | 0.28 | 0.12 | 0.19 | -0.46 | -0.60 | 0.21 | 0.58 | 0.56 |
| 9−NCD-Risk score[6] | -0.47 | -0.66 | 0.28 | 0.19 | 0.80 | 0.63 | -0.48 | -0.01 |
| 23−Nova UPF score[6] | -0.59 | -0.67 | 0.18 | 0.09 | 1.00 | 0.46 | -0.46 | 0.00 |

[1]Sample sizes are number of surveys. CHO, carbohydrates; E!, energy (kcal/day); FAO, Food and Agriculture Organization of the United Nations; GDQS−, Global Quality Score Negative; GDQS+, Global Quality Score Positive; GDR, Global Dietary Recommendations; GIFT, Global Individual Food consumption data Tool; HIC, high income countries; LIC, low income countries; LMIC, lower-middle income countries; MAR, mean adequacy ratio; NA, not applicable; NCD, non-communicable disease; SAFA, saturated fatty acids; UPF, ultra-processed food; UMIC, upper-middle income countries; WHO, World Health Organization.
[2]Surveys including non-pregnant females aged 18-49 years.
[3]Surveys including non-pregnant females aged 20-49 years.
[4]Surveys including non-pregnant females aged 16-49 years.
[5]Vitamin A, vitamin C, folate, calcium, iron, and zinc. The selection of micronutrients was informed by the prevalence of nutrient deficiencies globally and the availability of dietary intake data from surveys on FAO/WHO GIFT (i.e., leading to the exclusion of vitamin D, iodine, and vitamin K in the composite measure).
[6]Likewise to GDQS−, higher scores of the complement of NCD-Risk and Nova UPF scores reflect lower consumption of food groups associated with increased risk for diet-related NCDs.

#### Table S9. Accuracy of healthy diet indicators for <10% dietary energy intake from free sugars among non-pregnant females aged 15-49 years using surveys in upper-middle income countries available open access on FAO/WHO GIFT[1]

| **Characteristic** | **Moderation** | | | | |
|---|---|---|---|---|---|
| **Outcome** | **<10% E! from free sugars** | | | | |
| **UMIC, *n*=34,546** | **Sensitivity** | **Specificity** | **PCC** | **LR+** | **LR−** |
| GDQS− ≥8 | 89.3 | 38.2 | 58.6 | 1.45 | 0.279 |
| GDQS− ≥9 | 79.6 | 56.9 | 65.9 | 1.85 | 0.359 |
| GDQS− ≥10 | 69.1 | 71.0 | 70.2 | 2.38 | 0.435 |
| GDQS− ≥11 | 50.0 | 86.2 | 71.7 | 3.61 | 0.581 |
| 9−NCD-Risk score ≥6[2] | 81.9 | 50.9 | 63.3 | 1.67 | 0.356 |
| 9−NCD-Risk score ≥7[2] | 62.3 | 75.0 | 69.9 | 2.49 | 0.503 |
| 9−NCD-Risk score ≥8[2] | 36.4 | 92.7 | 70.3 | 5.00 | 0.686 |
| 23−Nova UPF score ≥20[2] | 79.4 | 60.5 | 68.0 | 2.01 | 0.341 |
| 23−Nova UPF score ≥21[2] | 58.0 | 81.2 | 72.0 | 3.09 | 0.517 |
| 23−Nova UPF score ≥22[2] | 29.7 | 94.9 | 68.9 | 5.81 | 0.741 |
| No sweet foods | 69.4 | 77.3 | 74.1 | 3.05 | 0.397 |
| No sweet beverages | 52.6 | 83.8 | 71.3 | 3.24 | 0.566 |
| Either sweet foods or sweet beverages | 84.5 | 62.9 | 71.4 | 2.27 | 0.247 |
| No sweet foods or sweet beverages | 37.5 | 98.3 | 74.0 | 21.8 | 0.636 |

[1]Values are percentages, except for AUCs. AUC, area under the curve; E!, energy (kcal/day); FAO, Food and Agriculture Organization of the United Nations; GDQS−, Global Quality Score Negative; GIFT, Global Individual Food consumption data Tool; LR+; positive likelihood ratio; LR-, negative likelihood ratio; NCD, non-communicable disease; PCC, percentage correctly classified; UPF, ultra-processed foods; UMIC, upper-middle income countries; WHO, World Health Organization.

[2]Likewise to GDQS−, higher scores of the complement of NCD-Risk and Nova UPF scores reflect lower consumption of food groups associated with increased risk for diet-related NCDs.

#### Table S10. Spearman's rank correlations (ρ) between mean healthy diet measures and mean of secondary quantitative reference metrics of dietary intake using surveys available open access on FAO/WHO GIFT[1]

| Characteristic | Macronutrient balance | | | Moderation | | | | Nutrient adequacy | | |
|---|---|---|---|---|---|---|---|---|---|---|
| **Outcome** | **5-10% E! from *n*-6 PUFA** | **0.6-1.2% E! from *n*-3 PUFA** | **≥10% E! from PUFA** | **≤1% E! from TFA** | **<5% E! from free sugars** | **≥25 g/day of dietary fibre** | **% E! from UPF** | **MAR of 11 micronutrients[2]** | **≥3,510 mg/day of potassium[3]** | **≥250 mg/day of *n*-3 PUFA** |
| **Total, *n*** | 5 | 1 | 22 | 8 | 5 | 37 | 44 | 20 | 26 | 1 |
| FGDS | -0.90 | NA | 0.45 | 0.74 | -0.60 | 0.29 | 0.26 | 0.80 | -0.11 | NA |
| GDQS+ | -0.90 | NA | 0.11 | 0.67 | -0.80 | 0.63 | 0.17 | 0.61 | 0.27 | NA |
| GDQS− | 0.90 | NA | -0.36 | -0.07 | 0.30 | 0.26 | -0.52 | -0.43 | 0.39 | NA |
| NCD-Protect score | -0.90 | NA | 0.28 | 0.69 | -0.60 | 0.47 | 0.08 | 0.60 | 0.00 | NA |
| 9−NCD-Risk score[4] | 0.20 | NA | -0.31 | 0.21 | 0.80 | 0.51 | -0.53 | -0.34 | 0.36 | NA |
| 23−Nova UPF score[4] | -0.40 | NA | -0.31 | 0.43 | 1.00 | 0.51 | -0.81 | -0.36 | 0.50 | NA |

[1]Sample sizes are number of surveys. E!, energy (kcal/day); FAO, Food and Agriculture Organization of the United Nations; GDQS−, Global Quality Score Negative; GDQS+, Global Quality Score Positive; GIFT, Global Individual Food consumption data Tool; HIC, high income countries; LIC, low income countries; LMIC, lower-middle income countries; NA, not applicable; NCD, non-communicable disease; PUFA, poly-unsaturated fatty acids; TFA, trans-fatty acids; UPF, ultra-processed food; UMIC, upper-middle income countries; WHO, World Health Organization.
[2]Vitamin A, thiamine, riboflavin, niacin, vitamin B6, vitamin B12, vitamin C, folate, calcium, iron, and zinc.
[3]Among individuals aged 16 years and older.
[4]Likewise to GDQS−, higher scores of the complement of NCD-Risk and Nova UPF scores reflect lower consumption of food groups associated with increased risk for diet-related NCDs.

#### Table S11. Associations between unhealthy food group indicators and primary quantitative reference metrics of dietary intake for the characteristic moderation among non-pregnant females aged 15-49 years, by World Bank income classification using surveys available open access on FAO/WHO GIFT[1]

| Characteristic | Moderation | | |
|---|---|---|---|
| **Outcome** | **<2,000 mg/day sodium[2]** | **<10% E! from free sugars** | **≤10% E! from SAFA** |
| **LIC, *n*** | 448 | NA | 456 |
| No sweet foods | 1.66 (0.91, 3.04) | NA | 0.73 (0.48, 1.11)[6] |
| No sweet beverages[3] | 3.11 (1.65, 5.87) | NA | 0.72 (0.44, 1.17) |
| Either sweet foods or sweet beverages[4] | 2.31 (1.01, 5.25) | NA | 0.78 (0.39, 1.54) |
| No sweet foods and sweet beverages[4] | 4.99 (2.00, 12.4) | NA | 0.56 (0.28, 1.10) |
| No fried and salty foods[5] | 2.15 (0.52, 8.96) | NA | 0.44 (0.05, 3.71) |
| **LMIC, *n*** | 20,317 | NA | 7,850 |
| No sweet foods | 1.44 (1.34, 1.55) | NA | 2.85 (2.47, 3.30)[6] |
| No sweet beverages[3] | 0.84 (0.62, 1.14) | NA | 1.22 (1.02, 1.47) |
| Either sweets foods or sweet beverages[4] | 0.63 (0.44, 0.91) | NA | 1.24 (1.01, 1.52) |
| No sweet foods and sweet beverages[4] | 0.92 (0.64, 1.33) | NA | 2.00 (1.56, 2.56) |
| No fried and salty foods[5] | 0.85 (0.63, 1.14) | NA | 1.16 (0.57, 2.35) |
| **UMIC, *n*** | 30,333 | 34,546 | 38,436 |
| No sweet foods | 1.38 (1.31, 1.47) | 7.70 (7.33, 8.08)[6] | 1.64 (1.57, 1.71)[6] |
| No sweet beverages[3] | 1.61 (1.51, 1.71) | 5.72 (5.45, 6.02)[6] | 0.89 (0.86, 0.94) |
| Either sweets foods or sweet beverages[4] | 1.36 (1.28, 1.49) | 5.35 (5.06, 5.67)[6] | 1.22 (1.17, 1.28) |
| No sweet foods and sweet beverages[4] | 2.15 (1.96, 2.35) | 88.3 (78.6, 99.3)[6] | 1.44 (1.35, 1.53) |
| No fried and salty foods[5] | 2.01 (1.89, 2.13) | 3.06 (2.97, 3.14)[6] | 1.28 (1.22, 1.34) |
| **HIC, *n*** | 980 | NA | 1,866 |
| No sweet foods | 1.90 (1.43, 2.52) | NA | 1.56 (1.29, 1.89)[6] |
| No sweet beverages[3] | 1.25 (0.94, 1.67) | NA | 0.76 (0.61, 0.91) |
| Either sweets foods or sweet beverages[4] | 1.30 (0.93, 1.80) | NA | 1.14 (0.92, 1.41) |
| No sweet foods and sweet beverages[4] | 2.49 (1.66, 3.73) | NA | 0.93 (0.69, 1.24) |
| No fried and salty foods[5] | 1.23 (0.89, 1.70) | NA | 0.77 (0.62, 0.97) |

[1]Values are the odds ratios (95% CIs) from mixed effects logistic regression models with a random intercept for each survey. E!, energy (kcal/day); FAO, Food and Agriculture Organization of the United Nations; GIFT, Global Individual Food consumption data Tool; HIC, high income countries; LIC, low income countries; LMIC, lower-middle income countries; NA, not applicable; SAFA, saturated fatty acids; UMIC, upper-middle income countries; WHO, World Health Organization.
[2]Among non-pregnant females aged 16-49 years.
[3]Includes i) sugar-sweetened beverages and ii) sweetened infusions.
[4]Reference group is females consuming both sweet foods and beverages.
[5]Includes i) packaged salty snacks; ii) deep fried snacks; iii) instant noodles; and iv) fast food restaurant foods.
[6]Logistic regression due to failure of model convergence with a random intercept for each survey.

#### Table S12. Associations between mean-standardized healthy diet measures and nutrient adequacy ratios (NARs) of six micronutrients among non-pregnant females aged 15-49 years, by World Bank income classification using surveys available open access on FAO/WHO GIFT[1]

| **Characteristic** | **Nutrient adequacy** | | | | | |
|---|---|---|---|---|---|---|
| **Outcome** | **NAR of vitamin A** | **NAR of vitamin C** | **NAR of folate** | **NAR of calcium** | **NAR of iron** | **NAR of zinc** |
| **LIC, *n*** | 1,567 | 2,408 | 2,233 | 2,408 | 2,408 | 2,408 |
| FGDS | 0.13 (0.10, 0.15), 0.34[3] | 0.13 (0.08, 0.18), 0.27 | 0.15 (0.11, 0.19) | 0.13 (0.10, 0.16), 0.25 | 0.09 (0.08, 0.10), 0.20 | 0.11 (0.10, 0.13), 0.21 |
| GDQS+ | 0.07 (0.05, 0.09), 0.35[3] | 0.08 (0.05, 0.12), 0.28 | 0.13 (0.07, 0.19), 0.25 | 0.09 (0.08, 0.10), 0.26 | 0.08 (0.06, 0.11), 0.19 | 0.10 (0.08, 0.12), 0.20 |
| GDQS− | 0.02 (0.00, 0.04), 0.35[3] | -0.01 (-0.05, 0.03), 0.28 | -0.03 (-0.06, -0.01), 0.28 | 0.01 (-0.01, 0.05), 0.27 | -0.01 (-0.03, 0.01), 0.21 | -0.01 (-0.05, 0.01), 0.22 |
| NCD-Protect score | 0.12 (0.10, 0.14), 0.34[3] | 0.08 (0.04, 0.12), 0.27 | 0.14 (0.10, 0.18), 0.26 | 0.08 (0.06, 0.11), 0.26 | 0.08 (0.07, 0.09), 0.20 | 0.08 (0.07, 0.09), 0.21 |
| 9−NCD-Risk score[2] | 0.00 (-0.02, 0.02), 0.35[3] | 0.00 (-0.01, 0.01), 0.29 | -0.01 (-0.02, 0.00), 0.28 | 0.01 (0.00, 0.03), 0.27 | 0.00 (-0.02, 0.02), 0.21 | -0.03 (-0.05, 0.00), 0.22 |
| 23−Nova UPF score[2] | 0.00 (-0.02, 0.01), 0.35[3] | 0.02 (-0.03, 0.08), 0.28 | -0.03 (-0.04, -0.02), 0..28 | -0.04 (-0.10, 0.01), 0.27 | -0.02 (-0.04, 0.01), 0.21 | -0.05 (-0.10, 0.01), 0.22 |
| **LMIC, *n*** | 21,619 | 32,766 | 21,912 | 32,766 | 32,766 | 21,912 |
| FGDS | 0.14 (0.10, 0.17), 0.26 | 0.14 (0.12, 0.17), 0.29 | 0.11 (0.09, 0.14), 0.22 | 0.12 (0.10, 0.13), 0.23 | 0.07 (0.05, 0.09), 0.19 | 0.10 (0.07, 0.12), 0.21 |
| GDQS+ | 0.11 (0.08, 0.14), 0.26 | 0.12 (0.10, 0.14), 0.30 | 0.13 (0.09, 0.16), 0.20 | 0.10 (0.08, 0.12), 0.23 | 0.08 (0.06, 0.10), 0.17 | 0.09 (0.07, 0.11), 0.19 |
| GDQS− | -0.01 (-0.04, 0.02), 0.28 | -0.05 (-0.07, -0.03), 0.31 | -0.02 (-0.04, 0.00), 0.23 | 0.00 (-0.03, 0.02), 0.24 | -0.03 (-0.05, -0.01), 0.19 | -0.02 (-0.04, 0.01), 0.21 |
| NCD-Protect score | 0.11 (0.07, 0.15), 0.27 | 0.15 (0.12, 0.18), 0.29 | 0.13 (0.09, 0.17), 0.22 | 0.07 (0.06, 0.08), 0.24 | 0.06 (0.04, 0.08), 0.19 | 0.08 (0.05, 0.10), 0.21 |
| 9−NCD-Risk score[2] | -0.04 (-0.08, 0.00), 0.28 | -0.02 (-0.03, 0.00), 0.31 | -0.03 (-0.06, 0.00), 0.24 | -0.04 (-0.07, -0.02), 0.24 | -0.04 (-0.06, -0.02), 0.19 | -0.06 (-0.09, -0.03), 0.21 |
| 23−Nova UPF score[2] | -0.04 (-0.08, 0.01), 0.28 | -0.02 (-0.05, 0.01), 0.31 | -0.02 (-0.06, 0.02), 0.24 | -0.05 (-0.09, -0.02), 0.24 | -0.02 (-0.05, 0.02), 0.19 | -0.01 (-0.04, 0.02), 0.22 |
| **UMIC, *n*** | 26,496 | 39,787 | 38,436 | 40,077 | 39,787 | 39,787 |
| FGDS | 0.14 (0.11, 0.17), 0.29 | 0.12 (0.10, 0.14), 0.40 | 0.09 (0.06, 0.12), 0.25 | 0.11 (0.09, 0.13), 0.26 | 0.06 (0.04, 0.08), 0.19 | 0.09 (0.07, 0.11), 0.23 |
| GDQS+ | 0.10 (0.08, 0.13), 0.30 | 0.11 (0.08, 0.13), 0.40 | 0.09 (0.05, 0.12), 0.25 | 0.08 (0.07, 0.10), 0.27 | 0.05 (0.03, 0.07), 0.19 | 0.07 (0.05, 0.08), 0.24 |
| GDQS− | -0.02 (-0.03, -0.01), 0.31 | -0.09 (-0.13, -0.06), 0.39 | -0.05 (-0.06, -0.03), 0.26 | -0.02 (-0.03, 0.00), 0.27 | -0.05 (-0.07, -0.03), 0.19 | -0.05 (-0.07, -0.03), 0.24 |
| NCD-Protect score | 0.10 (0.07, 0.13), 0.30 | 0.12 (0.09, 0.16), 0.39 | 0.06 (0.03, 0.09), 0.26 | 0.06 (0.05, 0.08), 0.27 | 0.04 (0.02, 0.05), 0.19 | 0.04 (0.03, 0.06), 0.24 |
| 9−NCD-Risk score[2] | -0.05 (-0.08, -0.02), 0.31 | -0.03 (-0.06, -0.01), 0.41 | -0.04 (-0.06, -0.03), 0.26 | -0.05 (-0.07, -0.04), 0.27 | -0.05 (-0.07, -0.03), 0.19 | -0.07 (-0.09, -0.05), 0.24 |
| 23−Nova UPF score[2] | -0.08 (-0.12, -0.03), 0.30 | -0.05 (-0.08, -0.02), 0.40 | -0.06 (-0.08, -0.05), 0.26 | -0.09 (-0.11, 0.06), 0.26 | -0.05 (-0.07, -0.03), 0.19 | -0.06 (-0.08, -0.04), 0.24 |
| **HIC, *n*** | 1,867 | 1,867 | 1,867 | 1,867 | 1,867 | 1,012 |
| FGDS | 0.13 (0.12, 0.14), 0.23[3] | 0.12 (0.10, 0.13), 0.29[3] | 0.09 (0.08, 0.10), 0.20 | 0.12 (0.10, 0.13), 0.24[3] | 0.07 (0.05, 0.08), 0.14 | 0.12 (0.10, 0.14), 0.21 |
| GDQS+ | 0.07 (0.05, 0.08), 0.25[3] | 0.11 (0.10, 0.12), 0.28[3] | 0.07 (0.05, 0.08), 0.21 | 0.06 (0.05, 0.08), 0.26[3] | 0.04 (0.03, 0.06), 0.15 | 0.09 (0.07, 0.12), 0.22 |
| GDQS− | -0.02 (-0.03, 0.01), 0.26[3] | -0.07 (-0.08, -0.05), 0.30[3] | -0.05 (-0.06, -0.04), 0.21 | -0.01 (-0.02, 0.00), 0.26[3] | -0.04 (-0.06, -0.03), 0.15 | -0.06 (-0.08, -0.04), 0.23 |
| NCD-Protect score | 0.09 (0.07, 0.10), 0.24 | 0.13 (0.11, 0.14), 0.28[3] | 0.06 (0.04, 0.07), 0.21 | 0.06 (0.05, 0.07), 0.26[3] | 0.04 (0.03, 0.05), 0.15 | 0.07 (0.04, 0.10), 0.23 |
| 9−NCD-Risk score[2] | -0.03 (-0.04, -0.02), 0.25[3] | -0.01 (-0.03, 0.00), 0.30[3] | -0.05 (-0.07, -0.02), 0.21 | -0.05 (-0.06, -0.04), 0.26[3] | -0.05 (-0.07, -0.02), 0.15 | -0.09 (-0.13, -0.04), 0.22 |
| 23−Nova UPF score[2] | -0.04 (-0.05, -0.03), 0.25[3] | -0.04 (-0.06, -0.03), 0.30[3] | -0.08 (-0.11, -0.05), 0.20 | -0.07 (-0.09, -0.06), 0.25[3] | -0.07 (-0.10, -0.03), 0.14 | -0.08 (-0.11, -0.06), 0.22 |

[1]Healthy diet measures were transformed to age range specific *z*-scores using the mean and SD within each World Bank income classification. Values are coefficients (95% CIs) and SD of the residuals from mixed effects linear regression models with a random intercept for each survey and a random slope for the healthy diet measure. FAO, Food and Agriculture Organization of the United Nations; GDQS−, Global Quality Score Negative; GDQS+, Global Quality Score Positive; GIFT, Global Individual Food consumption data Tool; HIC, high income countries; LIC, low income countries; LMIC, lower-middle income countries; NCD, non-communicable disease; UPF, ultra-processed food; UMIC, upper-middle income countries; WHO, World Health Organization.
[2]Likewise to GDQS−, higher scores of the complement of NCD-Risk and Nova UPF scores reflect lower consumption of food groups associated with increased risk for diet-related NCDs.
[3]Mixed effects linear regression model with a random intercept for each survey only.

### Table S13. Accuracy of healthy diet indicators for mean adequacy ratio (MAR) of six micronutrients (vitamin A, vitamin C, folate, calcium, iron, and zinc) ≥0.70 and ≥400 grams per day of fruits and vegetables among non-pregnant females aged 15-49 years, by World Bank income classification using surveys available open access on FAO/WHO GIFT[1]

| Characteristic | Nutrient adequacy | | | | | | | | | | |
|---|---|---|---|---|---|---|---|---|---|---|---|
| **Outcome** | **MAR of six micronutrients ≥0.70** | | | | | **Outcome** | **≥400 g/day of fruits and vegetables** | | | | |
| **LIC, *n*=1,392** | **Sensitivity** | **Specificity** | **PCC** | **LR+** | **LR−** | **LIC, *n*=2,408** | **Sensitivity** | **Specificity** | **PCC** | **LR+** | **LR−** |
| FGDS ≥4 | 91.6 | 44.4 | 65.7 | 1.65 | 0.190 | FGDS ≥4 | 83.7 | 59.6 | 66.3 | 2.07 | 0.27 |
| FGDS ≥5 | 67.6 | 74.4 | 71.3 | 2.64 | 0.436 | FGDS ≥5 | 57.6 | 83.4 | 76.3 | 3.47 | 0.509 |
| FGDS ≥6 | 37.4 | 92.7 | 67.7 | 5.09 | 0.676 | FGDS ≥6 | 29.3 | 94.0 | 76.0 | 4.90 | 0.752 |
| GDQS+ ≥6 | 85.7 | 41.3 | 61.4 | 1.46 | 0.347 | GDQS+ ≥6 | 79.4 | 42.9 | 53.0 | 1.39 | 0.481 |
| GDQS+ ≥7 | 74.9 | 59.5 | 66.5 | 1.85 | 0.422 | GDQS+ ≥7 | 62.5 | 58.5 | 59.6 | 1.51 | 0.641 |
| GDQS+ ≥8 | 66.1 | 65.8 | 66.0 | 1.93 | 0.515 | GDQS+ ≥8 | 53.8 | 63.1 | 60.5 | 1.46 | 0.73 |
| GDQS+ ≥9 | 54.4 | 84.0 | 70.6 | 3.40 | 0.543 | GDQS+ ≥9 | 42.0 | 79.2 | 68.9 | 2.02 | 0.732 |
| NCD-Protect ≥3 | 87.7 | 51.5 | 67.9 | 1.81 | 0.238 | NCD-Protect ≥3 | 82.2 | 55.0 | 62.6 | 1.83 | 0.323 |
| NCD-Protect ≥4 | 64.7 | 76.7 | 71.3 | 2.77 | 0.460 | NCD-Protect ≥4 | 60.5 | 81.6 | 75.8 | 3.29 | 0.484 |
| NCD-Protect ≥5 | 34.3 | 92.3 | 66.1 | 4.44 | 0.712 | NCD-Protect ≥5 | 30.3 | 94.6 | 76.7 | 5.61 | 0.736 |
| **LMIC, *n*=20,101** | **Sensitivity** | **Specificity** | **PCC** | **LR+** | **LR−** | **LMIC, *n*=32,766** | **Sensitivity** | **Specificity** | **PCC** | **LR+** | **LR−** |
| FGDS ≥4 | 86.7 | 52.5 | 64.8 | 1.82 | 0.254 | FGDS ≥4 | 90.1 | 48.8 | 51.9 | 1.76 | 0.203 |
| FGDS ≥5 | 66.3 | 86.7 | 79.3 | 4.97 | 0.389 | FGDS ≥5 | 76.3 | 76.6 | 76.6 | 3.26 | 0.310 |
| FGDS ≥6 | 39.3 | 97.2 | 76.3 | 14.3 | 0.624 | FGDS ≥6 | 51.6 | 91.1 | 88.2 | 5.82 | 0.531 |
| GDQS+ ≥6 | 87.2 | 48.6 | 62.6 | 1.70 | 0.263 | GDQS+ ≥6 | 87.4 | 49.6 | 52.4 | 1.74 | 0.253 |
| GDQS+ ≥7 | 76.2 | 69.8 | 72.1 | 2.52 | 0.341 | GDQS+ ≥7 | 76.2 | 66.0 | 66.7 | 2.24 | 0.360 |
| GDQS+ ≥8 | 67.9 | 77.4 | 73.9 | 3.00 | 0.415 | GDQS+ ≥8 | 68.4 | 73.6 | 73.2 | 2.59 | 0.429 |
| GDQS+ ≥9 | 51.1 | 92.2 | 77.3 | 6.55 | 0.531 | GDQS+ ≥9 | 55.5 | 85.6 | 83.3 | 3.84 | 0.520 |
| NCD-Protect ≥3 | 81.6 | 38.4 | 54.0 | 1.32 | 0.480 | NCD-Protect ≥3 | 83.4 | 48.2 | 50.9 | 1.61 | 0.344 |
| NCD-Protect ≥4 | 51.6 | 73.1 | 65.3 | 1.92 | 0.66 | NCD-Protect ≥4 | 57.7 | 75.8 | 74.5 | 2.38 | 0.559 |
| NCD-Protect ≥5 | 20.4 | 94.5 | 67.7 | 3.70 | 0.843 | NCD-Protect ≥5 | 27.4 | 93.8 | 88.9 | 4.45 | 0.774 |
| **UMIC, *n*=25,145** | **Sensitivity** | **Specificity** | **PCC** | **LR+** | **LR−** | **UMIC, *n*=40,077** | **Sensitivity** | **Specificity** | **PCC** | **LR+** | **LR−** |
| FGDS ≥4 | 91.5 | 27.0 | 52.4 | 1.25 | 0.316 | FGDS ≥4 | 93.0 | 27.9 | 33.6 | 1.29 | 0.251 |
| FGDS ≥5 | 69.7 | 57.9 | 62.5 | 1.65 | 0.523 | FGDS ≥5 | 75.9 | 57.2 | 58.9 | 1.77 | 0.42 |
| FGDS ≥6 | 36.5 | 85.0 | 65.9 | 2.43 | 0.747 | FGDS ≥6 | 49.8 | 83.5 | 80.5 | 3.01 | 0.602 |
| GDQS+ ≥5 | 77.9 | 42.0 | 56.1 | 1.34 | 0.53 | GDQS+ ≥6 | 75.9 | 49.9 | 52.2 | 1.51 | 0.483 |
| GDQS+ ≥6 | 69.2 | 53.3 | 59.6 | 1.48 | 0.578 | GDQS+ ≥7 | 62.8 | 68.2 | 67.8 | 1.98 | 0.546 |
| GDQS+ ≥7 | 54.6 | 67.6 | 62.5 | 1.69 | 0.67 | GDQS+ ≥8 | 54.7 | 76.3 | 74.4 | 2.30 | 0.594 |
| GDQS+ ≥8 | 43.1 | 77.3 | 63.8 | 1.90 | 0.74 | GDQS+ ≥9 | 38.5 | 88.1 | 83.8 | 3.24 | 0.698 |
| NCD-Protect ≥2 | 81.6 | 35.7 | 53.8 | 1.27 | 0.52 | NCD-Protect ≥3 | 73.7 | 72.1 | 72.2 | 2.64 | 0.365 |
| NCD-Protect ≥3 | 49.6 | 71.0 | 62.6 | 1.71 | 0.71 | NCD-Protect ≥4 | 42.0 | 90.5 | 86.3 | 4.42 | 0.641 |
| NCD-Protect ≥4 | 22.4 | 89.8 | 63.3 | 2.21 | 0.86 | NCD-Protect ≥5 | 17.1 | 97.3 | 90.2 | 6.20 | 0.853 |
| **HIC, *n*=1,012** | **Sensitivity** | **Specificity** | **PCC** | **LR+** | **LR−** | **HIC, *n*=1,867** | **Sensitivity** | **Specificity** | **PCC** | **LR+** | **LR−** |
| FGDS ≥4 | 88.0 | 50.6 | 78.3 | 1.78 | 0.238 | FGDS ≥5 | 89.5 | 45.3 | 61.8 | 1.64 | 0.231 |
| FGDS ≥5 | 75.4 | 78.0 | 76.1 | 3.42 | 0.315 | FGDS ≥6 | 65.0 | 71.9 | 69.3 | 2.31 | 0.487 |
| FGDS ≥6 | 53.0 | 92.8 | 63.3 | 7.34 | 0.507 | FGDS ≥7 | 30.7 | 90.3 | 68.0 | 3.15 | 0.767 |
| GDQS+ ≥4 | 83.3 | 43.7 | 73.0 | 1.48 | 0.382 | GDQS+ ≥6 | 83.8 | 51.3 | 63.4 | 1.72 | 0.316 |
| GDQS+ ≥5 | 69.6 | 61.6 | 67.5 | 1.81 | 0.494 | GDQS+ ≥7 | 67.7 | 67.9 | 67.8 | 2.11 | 0.476 |
| GDQS+ ≥6 | 60.8 | 70.0 | 63.1 | 2.02 | 0.56 | GDQS+ ≥8 | 55.7 | 75.6 | 68.1 | 2.28 | 0.587 |
| GDQS+ ≥7 | 43.1 | 85.2 | 54.1 | 2.91 | 0.668 | GDQS+ ≥9 | 41.0 | 85.9 | 69.2 | 2.91 | 0.687 |
| NCD-Protect ≥3 | 63.2 | 73.8 | 65.9 | 2.41 | 0.500 | NCD-Protect ≥3 | 81.6 | 59.8 | 68.0 | 2.03 | 0.307 |
| NCD-Protect ≥4 | 29.9 | 94.7 | 46.7 | 5.62 | 0.740 | NCD-Protect ≥4 | 44.6 | 84.9 | 69.8 | 2.95 | 0.653 |

[1]Values are percentages, except for AUCs. AUC, area under the curve; E!, energy (kcal/day); FAO, Food and Agriculture Organization of the United Nations; FGDQ, food group diversity score; GDQS+, Global Quality Score Positive; GIFT, Global Individual Food consumption data Tool; HIC, high income countries; LIC, low income countries; LMIC, lower-middle income countries; LR+; positive likelihood ratio; LR-, negative likelihood ratio; NCD, non-communicable disease; PCC, percentage correctly classified; UMIC, upper-middle income countries; WHO, World Health Organization.

#### Table S14. Associations between the six-point Fruits and Vegetables-Global Dietary Recommendation (FV-GDR) score and primary quantitative reference metrics of dietary intake for the characteristic nutrient adequacy among non-pregnant females aged 15-49 years, by World Bank income classification using surveys available open access on FAO/WHO GIFT[1]

| **Characteristic** | | |
|---|---|---|
| **Outcome** | **MAR of six micronutrients**[2] | **≥400 g/day of fruits and vegetables**[3] |
| **LIC, *n*** | 1,392 | 2,408 |
| FV-GDR | 0.07 (0.02, 0.13), 0.16 | 5.41 (4.48, 6.54) |
| **LMIC, *n*** | 20,101 | 32,766 |
| FV-GDR | 0.08 (0.05, 0.11), 0.16 | 3.14 (2.95, 3.34) |
| **UMIC, *n*** | 25,145 | 40,077 |
| FV-GDR | 0.06 (0.05, 0.07), 0.18 | 3.74 (3.59, 3.89) |
| **HIC, *n*** | 1,012 | 1,867 |
| FV-GDR | 0.08 (0.07, 0.10), 0.18 | 3.48 (2.99, 4.06) |

[1]Healthy diet measures were transformed to age range specific *z*-scores using the mean and SD within each World Bank income classification. Unless otherwise stated, values are coefficients (95% CIs) and SD of the residuals from mixed effects linear regression models with a random intercept for each survey and a random slope for the healthy diet measure. FAO, Food and Agriculture Organization of the United Nations; GIFT, Global Individual Food consumption data Tool; HIC, high income countries; LIC, low income countries; LMIC, lower-middle income countries; MAR, mean adequacy ratio UMIC, upper-middle income countries; WHO, World Health Organization.
[2]Vitamin A, vitamin C, folate, calcium, iron, and zinc. The selection of micronutrients was informed by the prevalence of nutrient deficiencies globally and the availability of dietary intake data from surveys on FAO/WHO GIFT (i.e., leading to the exclusion of vitamin D, iodine, and vitamin K in the composite measure).
[3]Values are the odds ratios (95% CIs) from mixed effects logistic regression models with a random intercept for each survey.
